# The Return on Investment of Scaling Tuberculosis Screening and Preventive Treatment: A Modelling Study in Brazil, Georgia, Kenya, and South Africa

**DOI:** 10.1101/2024.03.12.24303930

**Authors:** Juan F. Vesga, Mona Salaheldin Mohamed, Monica Shandal, Elias Jabbour, Nino Lomtadze, Mmamapudi Kubjane, Anete Trajman, Gesine Meyer-Rath, Zaza Avaliani, Wesley Rotich, Daniel Mwai, Julio Croda, Hlengani T. Mathema, Immaculate Kathure, Rhoda Pola, Fernanda Dockhorn Costa, Norbert O. Ndjeka, Maka Danelia, Maiko L. Tonini, Nelly Solomonia, Daniele M. Pelissari, Dennis Falzon, Cecily Miller, Ines Garcia Baena, Nimalan Arinaminpathy, Kevin Schwartzman, Saskia Den Boon, Jonathon R. Campbell

**Author notes:** Correspondence to: Jonathon R. Campbell, McGill University, Office 3D.55, 5252 Boulevard de Maisonneuve O, Montréal, Québec, H4A 3S5.

## Abstract

**Background:** Closing the tuberculosis diagnostic gap and scaling-up tuberculosis preventive treatment (TPT) are two major global priorities to end the tuberculosis epidemic. To help support these efforts, we modeled the impact and return-on-investment (ROI) of a comprehensive intervention to improve tuberculosis screening and prevention in Brazil, Georgia, Kenya, and South Africa—four distinct epidemiological settings.

**Methods:** We worked with national tuberculosis programmes (NTP) in each country to define a set of interventions (“the intervention package”) related to tuberculosis screening and TPT in three priority populations: people with HIV, household contacts, and a country-defined high-risk population. We developed transmission models calibrated to tuberculosis epidemiology for each country, and collated cost data related to tuberculosis-related activities and patient costs in 2023 $USD. We compared the intervention package without and with TPT scaled-up to reach priority populations to a status quo scenario based on projected tuberculosis epidemiology over a 27-year time horizon (2024-2050). Outcomes were health system and societal costs, number of tuberculosis episodes, tuberculosis deaths, and disability adjusted life years (DALYs). We performed 1000 simulations and calculated the mean and 95% uncertainty range (95%UR) difference in outcomes between the intervention package and the status quo. We calculated the health system cost per DALY averted and societal return on the health system investment for each country. We did not discount costs or outcomes in the base scenario.

**Findings:** Under the status quo, by 2050, tuberculosis incidence is projected to be 39 (95%UR 37-43), 34 (24-50), 204 (186-255), and 208 (124-293) per 100,000 population in Brazil, Georgia, Kenya, and South Africa, respectively. Implementing the intervention package without TPT is projected to reduce tuberculosis incidence by 9.6% (95%UR 9.3-10), 14.4% (11-19.6), 30.3% (29-33.1), and 22.7% (19.4-27.2) in Brazil, Georgia, Kenya, and South Africa, respectively, by 2050. The addition of TPT is projected to further reduce tuberculosis incidence by 9.5% (95%UR 9.3-9.8), 10.9% (9.8-12.3), 19.2% (17.6-20.1), and 13.1% (11.2-14.4%). From the health system perspective, the incremental cost per DALY averted of the intervention package is $771 in Brazil, $1402 in Georgia, $521 in Kenya, and $163 in South Africa. The societal return per $1 invested by the health system is projected to be $10.80, $3.70, $27.40, and $39.00 in Brazil, Georgia, Kenya, and South Africa, respectively.

**Interpretation:** Scaling-up interventions related to tuberculosis screening and TPT in priority populations is projected to substantially reduce tuberculosis incidence and provide large returns on investment.

**Funding:** World Health Organization.

## Introduction

Though annual tuberculosis deaths decreased in 2022 after steady rises in 2020 and 2021 due to the COVID-19 pandemic, tuberculosis remains the second-leading cause of death due to a single infectious agent globally.^1^ The World Health Organization (WHO) estimates that in 2022, 10.6 million people fell ill with tuberculosis disease and 1.3 million died from tuberculosis, including 167,000 people with HIV. Despite these staggering numbers, approximately 3.1 million (29%) people with tuberculosis disease remain undiagnosed annually, failing to receive lifesaving treatment.^1^ One of the most effective ways to prevent tuberculosis disease is to provide tuberculosis preventive treatment (TPT).^2^ Yet despite a global commitment at the United Nations High Level Meeting (UNHLM) on tuberculosis in 2018 to provide TPT to 30 million people with HIV and contacts of people with tuberculosis disease between 2018 and 2022,^3^ only 15.5 million (52%) people—the vast majority people with HIV—received TPT.^1^

These gaps in tuberculosis disease detection and TPT administration exist despite their widely recognized benefits. Systematic screening to close the tuberculosis diagnosis gap leads to early identification of individuals with tuberculosis^4^ and identification of the sizable proportion of individuals who do not have recognizable symptoms (i.e., subclinical disease), reducing transmission.^5^ Additionally, currently available TPT regimens reduce the risk of future tuberculosis disease by 60-90%,^2^ including in tuberculosis endemic settings.^6^ Despite these benefits, a key barrier to national tuberculosis programmes (NTP) scaling-up tuberculosis screening and prevention activities is uncertainty surrounding their return-on-investment (ROI). Therefore, an understanding of the costs and epidemiologic impact of these activities may motivate scale-up and could be used to advocate for their necessary funding.

The aim of this study is to project the epidemiologic impact and ROI of scaling-up tuberculosis screening and prevention among priority, high-risk groups in Brazil, Georgia, Kenya, and South Africa, four countries with varying burdens of tuberculosis and distinct tuberculosis epidemiology.

## Methods

### Study Design and Procedures

This is a modeling study and economic evaluation assessing the impact and societal ROI of scaling tuberculosis screening and prevention activities among priority high-risk groups from 2024 through 2050. It was developed with the WHO Global Tuberculosis Programme, and conducted in partnership with NTPs in Brazil, Georgia, Kenya, and South Africa, who were supported by WHO country and regional offices. NTPs provided input into populations and interventions evaluated as well as the overall study objectives, validated (and provided, when necessary) epidemiological and cost data used to parametrize the transmission model and conduct the economic evaluation, reviewed output data for consistency and accuracy (such as epidemiological calibrations and analytical outputs), and participated in the interpretation and dissemination of findings. Interim findings were formally presented to participating tuberculosis programmes at two webinars (September and November 2023), where additional suggestions were received.^7^

### Settings

In Brazil, Georgia, Kenya, and South Africa, priority populations at high-risk of tuberculosis include people with HIV and household contacts (HHC) of people with tuberculosis disease. However, tuberculosis treatment coverage and extent of TPT implementation among these two priority populations vary.^8^ Beyond these groups, each NTP identified specific high-risk groups and/or communities^9^ that would also benefit from tuberculosis active case finding and prevention activities. These included people deprived of liberty in Brazil, people accessing care for injection drug use in Georgia, people living in informal settlements within 9 high-tuberculosis prevalence counties in Kenya, and people living in the 22 highest tuberculosis prevalence subdistricts in South Africa (**Table 1**; Appendix p3).

**Table 1.** Country epidemiology in 2022.

| Country | Population (2022) | Estimated TB Incidence per 100,000 population | TB Notification Rate* | TB Case Fatality Ratio** | HIV-coinfection*** | RR-TB*** | TPT Coverage | Additional High-Risk Group |
| --- | --- | --- | --- | --- | --- | --- | --- | --- |
| **Brazil** | 203 million | 49 (42-56) | 83% (72-98) | 11% (9-13) | 18% | 2.7% | PLHIV: 30%<br>HHC: 18% | People deprived of liberty (650,000 people, TB incidence of 1.1%) |
| **Georgia** | 3.7 million | 60 (48-73) | 68% (55-84) | 4% (3-5) | 2.9% | 16.4% | PLHIV: 4%<br>HHC: 19% | People accessing treatment for injection drug use (17,500 people, TB prevalence of 0.9%) |
| **Kenya** | 54 million | 237 (149-363) | 69% (45-110) | 22% (11-35) | 22.7% | 1% | PLHIV: 32%<br>HHC: 18% | People living in informal settlements in 9 high TB prevalence counties (416,000 people, TB prevalence of 0.75%) |
| **South Africa** | 62 million | 468 (304-665) | 77% (54-120) | 20% (9-34) | 54.3% | 3.9% | PLHIV: 62%<br>HHC: 6% | People living in the 22 highest TB prevalence subdistricts (1.1 million people, TB prevalence of 1.9%) |

\*The proportion of all people estimated to develop tuberculosis, who are detected by the country; \*\*the estimated proportion of people estimated to develop tuberculosis, who die; \*\*\*calculated based on the estimated proportion of all people who developed tuberculosis who had HIV or had rifampicin-resistant tuberculosis.
Tuberculosis data derived from the WHO Global TB Programme Data Repository (https://www.who.int/teams/global-tuberculosis-programme/data)
Abbreviations: TB, tuberculosis; HIV, human immunodeficiency virus; RR-TB, rifampicin-resistant tuberculosis; TPT, tuberculosis preventive treatment; PLHIV, people living with HIV; HHC, household contacts.

### Interventions

In each country and for each priority population, we compared a package of interventions related to tuberculosis screening and prevention (referred to as “the intervention package”) to the status quo (i.e., current practice for diagnosing and treating tuberculosis disease and infection). The status quo treatment and diagnostic algorithms for each country are described in the **Appendix** (pp4-6).

The intervention package was iteratively developed with participating countries to incorporate perceived-to-be feasible levels of algorithm coverage along with implementation of currently available and recommended diagnostics and treatments (**Table 2**; **Appendix** p7).^9–11^ Diagnostic and treatment algorithms, and their coverage, were specific to each population group and stratified by age (<5 years, 5-9 years, 10-14 years, and ≥15 years). When implementing the intervention package, the status quo level of screening and treatment among groups not targeted were assumed to remain constant, while we assumed no impact of the scale-up of the intervention package on cascades of care (aside from closing the screening gap). We assumed TPT was only provided to individuals who had had tuberculosis disease ruled out according to diagnostic algorithms and was only provided once. We assumed that by 2030, all people treated for RR-TB in each country would receive short regimens.^12^

**Table 2.** Description of The Intervention Package.

| Population | Percent of Population Reached by Algorithm | Age Group (years) | Tuberculosis Disease Algorithm | Tuberculosis Infection Algorithm (Assumes Tuberculosis Disease is Already Ruled Out)* |
| --- | --- | --- | --- | --- |
| People living with HIV not on ART | 90% | 0-4 | Systematic tuberculosis symptom screening; Xpert Ultra if symptoms present. | No testing for tuberculosis infection; 3HR prescribed for those <2 years and 3HP for those 2-4 years. |
|  |  | 5-9 | Systematic tuberculosis symptom screening; Xpert Ultra if symptoms present. | No testing for tuberculosis infection; 3HP prescribed. |
|  |  | ≥10 | Systematic tuberculosis symptom screening; CRP performed among those with symptoms. Xpert Ultra if symptoms present and CRP positive. | No testing for tuberculosis infection; 3HP prescribed. |
| People living with HIV on ART | 90% | 0-4 | Annual screening for tuberculosis using symptom screening; Xpert Ultra if symptoms present. | No testing for tuberculosis infection; 3HR prescribed for those <2 years and 3HP for those 2-4 years. |
|  |  | ≥5 | Annual screening for tuberculosis using symptom screening; Xpert Ultra if symptoms present. | No testing for tuberculosis infection; 3HP prescribed. |
| Household Contacts | 90% | 0-4 | Systematic symptom screening and chest x-ray; Xpert Ultra if symptoms present or abnormal chest x-ray | Testing with tuberculin skin test; if positive, 3HR prescribed for those <2 years and 3HP for those 2-4 years. |
|  | 50% | ≥5 | Systematic symptom screening and chest x-ray; Xpert Ultra if symptoms present or abnormal chest x-ray | Testing with tuberculin skin test; if positive, 3HP prescribed. |
| High-Risk Population | 0% | 0-14 | No specific intervention. | No specific intervention. |
|  | 60% | ≥15 | Systematic symptom screening and chest x-ray with CAD. Xpert Ultra if symptoms present or abnormal chest x-ray. The intervention is implemented in this population for 3 consecutive years (2024-2027) then halted. | No specific intervention. |

\*TPT is only provided one time in the model.
Abbreviations: HIV, human immunodeficiency virus; ART, antiretroviral treatment; CAD, computer aided detection; 3HR, 3 months of daily isoniazid and rifampicin; 3HP, 3 months of once-weekly isoniazid and rifapentine.

### Modeling Approach and Parameters

We designed a deterministic, compartmental multi-strain transmission model for tuberculosis, structured in age, HIV status, and subpopulation risk groups. The latter category is defined according to local epidemiology and country-specific consultations (see study design and procedures). The main model transitions (**Appendix** p8) representing the set of ordinary differential equations are described in detail in the **Appendix** (pp9-11). The model was coded and numerically solved using MATLAB (MathWorks, MA, USA).

#### Data and calibration

For each selected country, we tuned model inputs to capture local demographics before running a calibration procedure for fitting the model to available data, such as tuberculosis incidence, mortality, and rifampicin-resistance (**Appendix** p12). We used a Bayesian framework to systematically compare model outputs to different streams of epidemiological data (**Appendix** 13-16). For each country, we introduced country-specific changes to account for changes in tuberculosis trends caused by COVID-19 disruptions.^1^

#### Model Assumptions and Parameters

Select model, epidemiological, diagnostic, treatment, and health utility parameters are presented in **Table 3** (see **Appendix** pp17-21 for the full list of assumptions and parameters). When available, epidemiological parameters were sourced from the WHO^9–12^ and the literature; otherwise, they were provided by the NTP in each country. We accounted for tuberculosis-related morbidity and mortality using disability adjusted life years (DALYs).^13^ DALY estimates for tuberculosis disease came from the Global Burden of Disease Study, while we used DALY estimates from minor ailments (e.g., headache, low-grade anxiety, mild anemia, diarrhea) to approximate DALYs associated with treatment-related adverse events during tuberculosis treatment and TPT.^13^ DALYs among people surviving tuberculosis were assumed to be lifelong, with an annual value of 0.036.^14^ Years of life lost due to tuberculosis mortality were calculated based on life expectancy in each country at time of death from tuberculosis^15^ and on the excess mortality seen among tuberculosis survivors in the five-years post-tuberculosis.^14^

**Table 3.** Select Model Parameters.

| Parameter | Brazil | Georgia | Kenya | South Africa | References |
| --- | --- | --- | --- | --- | --- |
| Average number of transmission events arising from each person with infectious DS-TB per year | 7.1 (6.5 to 12) | 4 (2.5 to 6.4) | 18 (10.5 to 22) | 13.3 (9.3 to 19.8) | Model Estimate |
| Average number of transmission events arising from each person with infectious RR-TB per year | 5.3 (4 to 6.9) | 3.8 (2.2 to 6.1) | 12 (8 to 14) | 9.1 (7.5 to 13.7) | Model Estimate |
| Rate of Progression from TB Infection to Disease Per Year |  | "Slow" Progressors: 0.000594<br>"Fast" Progressors: 0.0826 |  |  | 35 |
| Annual Transition Probability from Fast-to-Slow Progressor |  | 87% |  |  | 35 |
| Proportion of people with HIV enrolled on ART initiating TPT | 28% (15 to 30) | 5% (1 to 10) | 35% (20 to 45) | 66% (52 to 82) | Model Estimate |
| Reduced Susceptibility to TB from past infection |  | Uniform Distribution from 25% to 75% |  |  | Assumed, <sup>36</sup> |
| HHC with TB Infection |  | 0-4 years: 35.5% (30.3 to 41.1)<br>5-14 years: 53.1% (42 to 63.9)<br>15 years or older: 65.3% (35.5 to 86.5) |  |  | 37 |
| HHC per Index Patient | 2.31 | 2.42 | 2.75 | 2.36 | 38 |
| Status Quo Use of Rifamycin-Based TPT, 2021* | 13% | 88% | 8% | 12% | 8 |
| Status Quo Use of Short RR-TB Treatment, most recent year available* | 5% | 67% | 5% | 65% | 8 |
| 3HP Serious Adverse Events |  | 0-14 years: 2%<br>15 years or older: 0.2% |  |  | 39 |
| 3HP Completion |  | 81% |  |  | 39 |
| 3HP Efficacy |  | 93% |  |  | 40 |
| TB-Associated DALY (per episode) |  | TB Alone: 0.333 (0.274 to 0.549)<br>TB-HIV Coinfection: 0.408 (0.274 to 0.549) |  |  | 13 |
| Post-TB DALY (annual) |  | 0.036 (0.006 to 0.088) |  |  | 14 |
| Adverse Event DALY (per event) |  | 0.02 (0.012 to 0.03) |  |  | Assumed, see methods |
| Xpert Cost, per test | \$20.88 (4.74 to 48.88) | \$18.74 (1.77 to 55.42) | \$19.91 (8.1 to 37.02) | \$17.93 (4.49 to 40.6) | Appendix pp22-35 |
| TST Cost, per test | \$5.59 (4.56 to 6.73) | \$5.08 (4.13 to 6.13) | \$10.28 (8.37 to 12.39) | \$4.05 (3.29 to 4.89) | |
| 3HP Cost (Adults, Complete Course) | \$36.74 (10.09 to 80.52) | \$50.08 (13.75 to 109.75) | \$28.65 (7.86 to 62.8) | \$21.39 (5.87 to 46.89) | |
| DS-TB Treatment Cost (Adults, per person treated) | \$567.07 (155.63 to 1242.76) | \$325.4 (89.3 to 713.13) | \$191.24 (52.48 to 419.12) | \$133.57 (36.65 to 292.74) | |
| Short RR-TB Treatment Cost (Adults, per person treated) | \$3104.77 (850.62 to 6796.16) | \$2379.05 (651.8 to 5207.61) | \$3385.46 (927.52 to 7410.61) | \$1030.71 (282.38 to 2256.18) | |
| Case-Finding Cost (per person) | \$10.25 (5.86 to 15.82) | \$13.14 (7.51 to 20.29) | \$6.12 (3.50 to 9.44) | \$19.4 (11.10 to 29.94) | |
| Cost of Post-TB Care (per person) | \$192.66 (4.89 to 710.23) | \$283.92 (7.21 to 1046.67) | \$102.53 (2.6 to 377.99) | \$546.78 (13.88 to 2015.69) | |
| Training & Implementation Costs (annual) | \$24.6 million (20 million to 29.6 million) | \$330,000 (269,000 to 397,000) | \$734,000 (598,000 to 884,000) | \$5.8 million (4.7 million to 7.0 million) | |
| Patient Cost, per month DS-TB Treatment | \$281.36 (35.22 to 771.31) | \$414.7 (51.9 to 1136.87) | \$57.18 (7.15 to 156.76) | \$110.4 (13.82 to 302.64) | |
| Patient Cost, per month TPT | \$25.25 (3.22 to 69.56) | \$37.67 (4.79 to 103.78) | \$5.19 (0.66 to 14.28) | \$10.03 (1.28 to 27.59) | |

\*Assumed to increase to 50% in the status quo scenario by 2030, then maintained at this level until 2050; if already above 50%, the level did not change during between 2024 and 2050.
Note: All costs are in 2023 \$USD. Values represent mean and (95% UR).
Abbreviations: TB, tuberculosis; DALY, disability adjusted life year; TST, tuberculin skin test; 3HP, 3-months of once-weekly isoniazid and rifapentine; DS-TB, drug-susceptible tuberculosis; RR-TB, rifampicin-resistant tuberculosis

### Costing Approach

We collated detailed cost information for tuberculosis-related activities in the following categories: (i) testing, (ii) treatment, (iii) post-tuberculosis care, (iv) implementation and scale-up, and (v) patient and societal costs. We did not consider patient or health system costs associated with care for other conditions (e.g., HIV, diabetes, chronic kidney disease). All costs are in 2023 USD. We used GDP deflator estimates to inflate costs from other years to 2023^16^ and if estimates were not in USD, we used country-specific conversion rates to convert costs to USD. Select cost estimates for each country are in **Table 3**, while all costs including detailed cost descriptions are in the **Appendix** (pp22-35). In general, we derived costs from data recently collected through the Value-TB project for Georgia and Kenya,^17,18^ from a micro-costing exercise component of South Africa’s strategic planning around tuberculosis, and through primary cost collection in Brazil. Where cost estimates were missing, we used the most recent, robust estimates from the literature.

Testing and treatment costs were inclusive of consumables, overhead, staff, and capital expenditures. For all people treated for tuberculosis disease, we included costs associated with the cascade of additional tests performed upon tuberculosis diagnosis (**Appendix** p36), and for those surviving, additional costs in the five years post-treatment.^19^ We considered specific costs associated with implementation, such as oversight and capital infrastructure (e.g., computer aided detection (CAD) for tuberculosis on chest x-ray). We used country-specific patient cost surveys or a recent meta-regression to estimate the monthly costs borne by patients associated tuberculosis disease;^20–22^ for people receiving TPT, we assumed per-month costs borne by the patient were 9% that of those associated with drug-susceptible tuberculosis.^23^ For every year of life lost (YLL) due to premature tuberculosis-related mortality, we assumed societal costs reflected lost productivity equivalent to the per-capita GDP in 2023. Additional cost methods and explanations are in the **Appendix** (p37)

### Outcomes and Data Analysis

The primary analysis was conducted from both a health system and societal perspective, with a 27-year time horizon (2024-2050), and 0% discount rate; this discount rate was chosen for the primary analysis to avoid discounting of health outcomes and paradoxical outcomes when varying discount rates for costs and outcomes are used.^24^ We defined societal costs as the sum of tuberculosis-related health system costs, tuberculosis-related costs to patients and families, and costs of lost productivity due to premature tuberculosis-related mortality. All model and economic parameters were fit to distributions and 1000 Latin hypercube samples drawn from each across 1000 model simulations. Epidemiological outputs from each model simulation were used to estimate year-by-year and cumulative epidemiological outcomes, health system costs, and societal costs. Based on the 1000 simulations, we estimated mean outcomes and 95% uncertainty ranges (UR) using the 2.5^th^ and 97.5^th^ percentile for the lower and upper bound, respectively. Analysis of epidemiological and economic outcomes was done in R version 4.1.0 (R Foundation).

In our primary analysis, we assumed interventions associated with tuberculosis screening and prevention among people with HIV and HHC were scaled up linearly to target coverage levels from 2024 to 2030, and then maintained until 2050, while among country-specific high-risk populations/communities, interventions were implemented at full coverage for the 3 years they were used. We report epidemiological and cost outcomes associated with implementing the intervention package with tuberculosis disease screening only, or in combination with TPT, as well as among the different priority populations.

We tabulated epidemiological and economic outcomes for each country and for the status quo and the intervention package without and with TPT from 2024 to 2050. These outcomes included (i) the number of people developing tuberculosis disease, (ii) the number of people dying from tuberculosis disease, (iii) years of life lost due to premature tuberculosis-related mortality, (iv) disability-adjusted life years (DALYs) associated with tuberculosis, (v) costs of tuberculosis to the health system, (vi) costs of tuberculosis to patients and families, and (vii) the societal cost of tuberculosis.

We compared outcomes between simulated strategies to estimate incremental differences between the intervention package without TPT vs. the status quo, as well as the intervention package with TPT vs. without TPT. For each of these comparisons, we also determined changes in the number of people screened for tuberculosis disease and initiating TPT.

We leveraged incremental differences in economic and epidemiological outcomes to calculate the incremental cost per (i) DALY averted, (ii) tuberculosis episode averted, and (iii) tuberculosis death averted from the health system and societal perspectives. We used country-specific willingness-to-pay thresholds per DALY averted to determine cost-effectiveness in 2023 USD. These values reflected health opportunity costs and were $13,644 for Brazil, $1603 for Georgia, $1002 for Kenya, and $4834 for South Africa.^25^

We calculated the societal ROI of implementing the intervention package with and without TPT. We calculated the societal ROI as the ratio of the incremental difference in societal costs to the incremental difference in health system costs between comparisons. We evaluated how the ROI evolved over the simulated time frame (from 2024 to 2050) and estimated the per capita incremental investment from the health system perspective for each of these years.

We conducted several scenario analyses: (i) we varied the discount rate to a universal rate of 3%, as well as income-specific discount rates of 4% for upper-middle income countries (Brazil, Georgia, South Africa) and 5% for lower-middle income countries (Kenya);^26^ (ii) we evaluated both rapid (2024-2026) and slow (2024-2050) scale-up and implementation of strategies; (iii) we evaluated outcomes on a shorter time horizon to 2035, in line with the END TB Strategy; (iv) we evaluated implementing strategies with no tuberculosis infection testing prior to TPT in priority groups; and (v) we evaluated an enhanced intervention package that was more ambitious, reaching nearly all individuals belonging to the priority groups and utilizing technologies that could foreseeably be broadly recommended in the near future (**Appendix** p38).

## Results

### Projections under the status quo, 2024 to 2050

Anticipated tuberculosis incidence trajectories **(Figure 1**) and other epidemiologic and economic outcomes (**Table 4**) under the status quo from 2024 through 2050 varied in the four countries evaluated. In Brazil, tuberculosis incidence is projected to be 39 (95%UR 37 to 43) per 100,000 by 2050, while total health system costs associated with tuberculosis from 2024 to 2050 are estimated to be $2.2 billion ($1.2 to $3.9 billion), with substantial societal costs of $81.2 billion ($75.4 to $90.2 billion). In Georgia, a country with a much smaller population than Brazil, tuberculosis incidence is projected to be 34 (24 to 50) per 100,000 by 2050, with health system costs of $49 million ($28 to $156 million) and societal costs $830 million ($630 million to $1.2 billion) from 2024 to 2050. Small changes in tuberculosis epidemiology are projected in Kenya, with a projected incidence of 204 (186 to 255) per 100,000 in 2050, accompanied by societal costs associated with tuberculosis of $41.0 billion ($36.4 to $51.9 billion), of which $3.2 billion ($1.5 to $6.6 billion) are health system costs from 2024 to 2050. Finally, in South Africa, tuberculosis incidence is projected to fall substantially by 2050 to 208 (124 to 293) per 100,000, however the economic costs associated with tuberculosis remain large, with $2.9 billion ($0.8 to $8.5 billion) in health system costs and $167.1 billion ($83.9 to $251.1 billion) in societal costs from 2024 to 2050.

**Figure 1.**
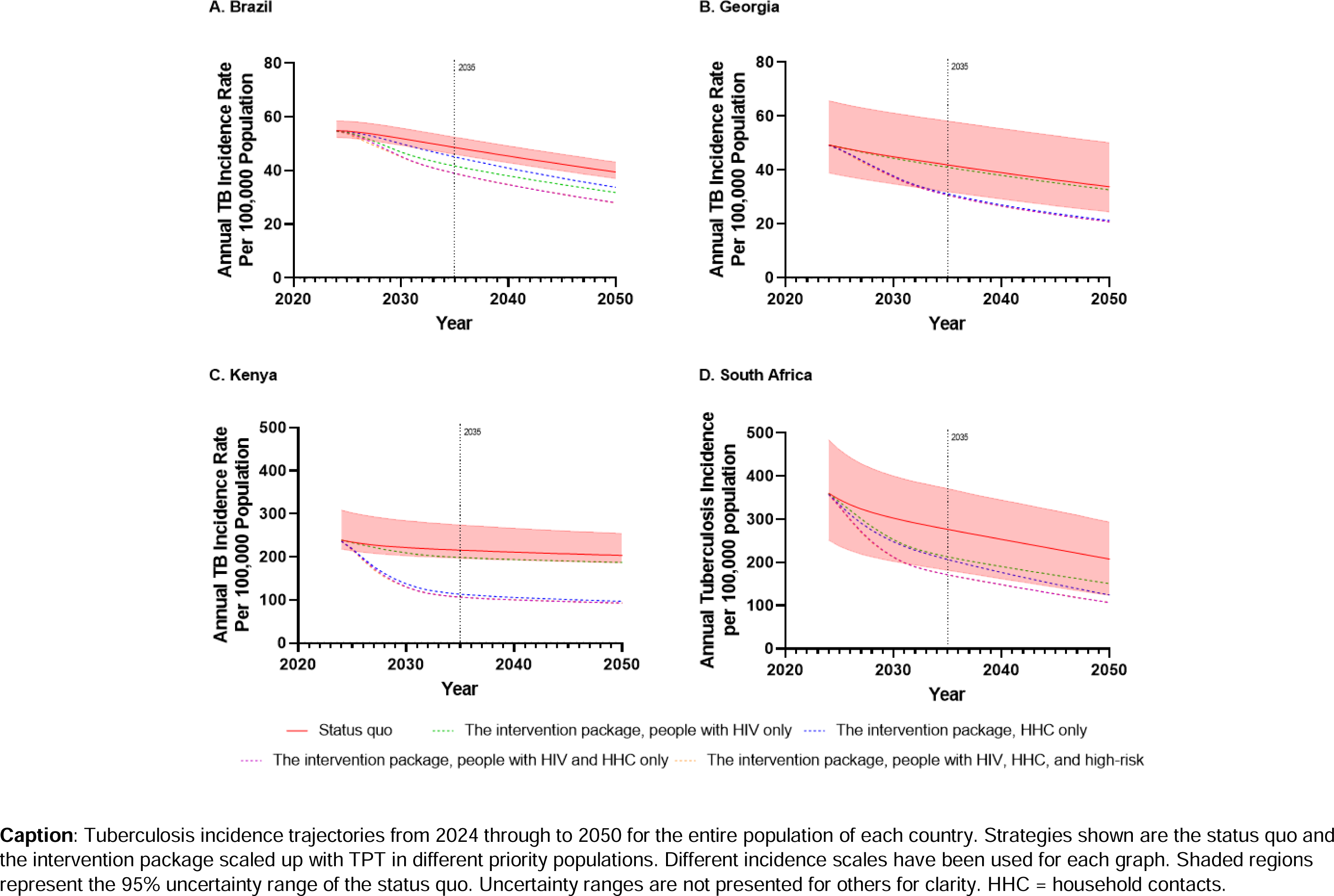
Incidence trajectories under the status quo and different scenarios.

**Table 4.** Outcomes under the status quo from 2024 to 2050. Values represent mean and (95% uncertainty range)

|  | Brazil | Georgia | Kenya | South Africa |
| --- | --- | --- | --- | --- |
| Outcome | 2024-2050 | 2024-2050 | 2024-2050 | 2024-2050 |
| Total People Developing TB | 2.7 million<br>(2.5 million to 2.9 million) | 42,000<br>(32,000 to 59,000) | 3.1 million<br>(2.9 million to 4.0 million) | 4.3 million<br>(2.8 million to 5.8 million) |
| Total People Dying from TB | 224,000<br>(211,000 to 244,000) | 3000<br>(2300 to 4000) | 454,000<br>(419,000 to 565,000) | 689,000<br>(328,000 to 1.1 million) |
| Total Years of Life Lost due to TB | 8.7 million<br>(8.1 million to 9.5 million) | 109,000<br>(84,000 to 151,000) | 17.8 million<br>(16.3 million to 22.5 million) | 24.0 million<br>(11.9 million to 36.3 million) |
| Total DALYs | 11.2 million<br>(10.6 million to 12.2 million) | 184,000<br>(148,000 to 243,000) | 20.4 million<br>(18.7 million to 25.5 million) | 27.8 million<br>(14.7 million to 40.8 million) |
| Total Health System Cost | \$2.2 billion<br>(\$1.2 billion to \$3.9 billion) | \$49 million<br>(\$28 million to \$85 million) | \$3.2 billion<br>(\$1.5 billion to \$6.6 billion) | \$2.9 billion<br>(\$0.8 billion to \$8.5 billion) |
| Total Costs to Patients and Families | \$1.7 billion<br>(\$326 million to \$4.6 billion) | \$64 million<br>(\$15 million to \$156 million) | \$427 million<br>(\$85 million to \$1.1 billion) | \$1.4 billion<br>(\$327 million to \$3.8 billion) |
| Total Societal Cost | \$81.2 billion<br>(\$75.4 billion to \$90.2 billion) | \$0.8 billion<br>(\$0.6 billion to \$1.2 billion) | \$41.0 billion<br>(\$36.4 billion to \$51.9 billion) | \$167.1 billion<br>(\$83.9 billion to \$251.1 billion) |

All costs are in \$USD and undiscounted over the time horizon. Abbreviations: TB, tuberculosis; DALY, disability adjusted life years

### Projections when implementing the intervention package, 2024 to 2050

Implementing systematic tuberculosis screening among the priority populations resulted in large increases in the number of people screened between 2024 and 2050, ranging from 9-fold in Kenya to more than 182-fold in Georgia (**Table 5, Appendix** pp39-40). The additional implementation of TPT resulted in even larger increases in the number of people initiating TPT, which varied depending on status quo level of TPT implementation. For example, in Georgia, systematic offering of TPT would result in an additional 760,000 (95%UR 639,000 to 954,000) courses of TPT being offered from 2024 to 2050.

**Table 5.**
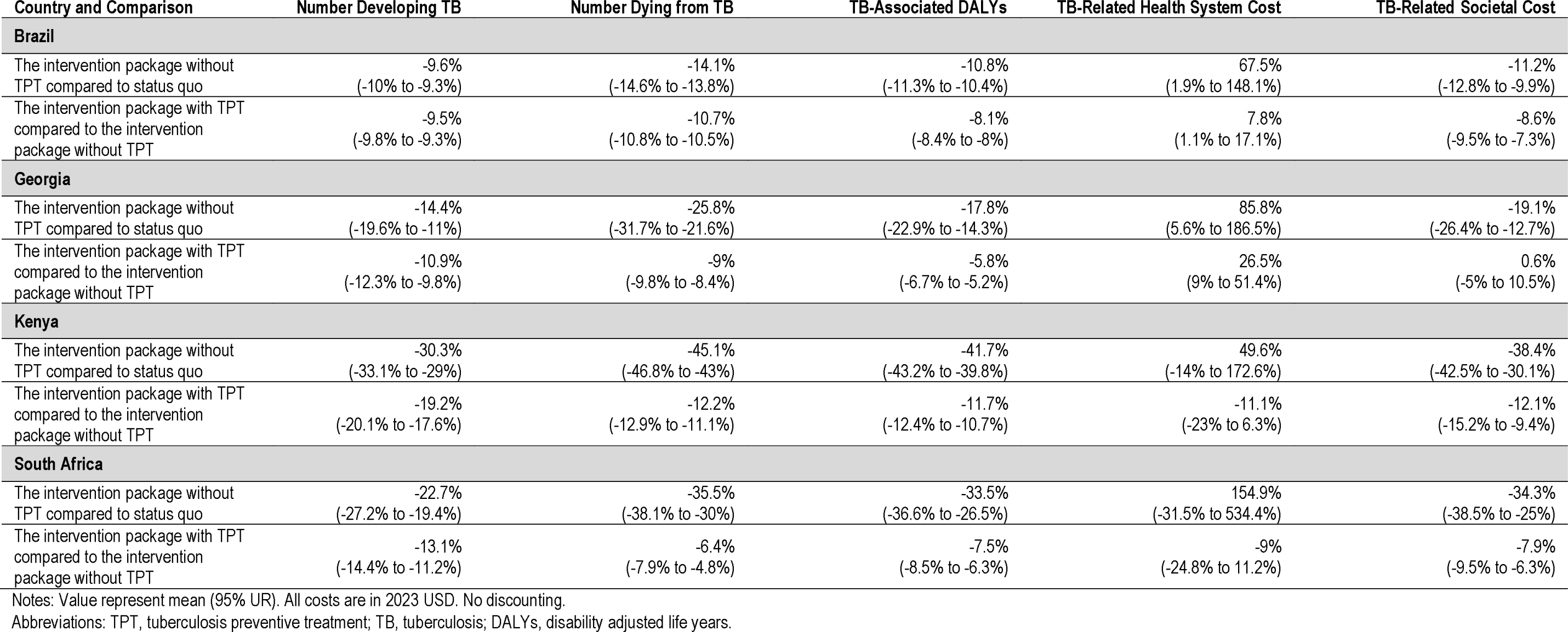
Relative Incremental Epidemiologic and Economic Outcomes by Country from 2024-2050. Values represent mean and (95% uncertainty range)

| Country and Comparison | Number Developing TB | Number Dying from TB | TB-Associated DALYs | TB-Related Health System Cost | TB-Related Societal Cost |
| --- | --- | --- | --- | --- | --- |
| **Brazil** |  |  |  |  |  |
| The intervention package without TPT compared to status quo | -9.6%<br>(-10% to -9.3%) | -14.1%<br>(-14.6% to -13.8%) | -10.8%<br>(-11.3% to -10.4%) | 67.5%<br>(1.9% to 148.1%) | -11.2%<br>(-12.8% to -9.9%) |
| The intervention package with TPT compared to the intervention package without TPT | -9.5%<br>(-9.8% to -9.3%) | -10.7%<br>(-10.8% to -10.5%) | -8.1%<br>(-8.4% to -8%) | 7.8%<br>(1.1% to 17.1%) | -8.6%<br>(-9.5% to -7.3%) |
| **Georgia** |  |  |  |  |  |
| The intervention package without TPT compared to status quo | -14.4%<br>(-19.6% to -11%) | -25.8%<br>(-31.7% to -21.6%) | -17.8%<br>(-22.9% to -14.3%) | 85.8%<br>(5.6% to 186.5%) | -19.1%<br>(-26.4% to -12.7%) |
| The intervention package with TPT compared to the intervention package without TPT | -10.9%<br>(-12.3% to -9.8%) | -9%<br>(-9.8% to -8.4%) | -5.8%<br>(-6.7% to -5.2%) | 26.5%<br>(9% to 51.4%) | 0.6%<br>(-5% to 10.5%) |
| **Kenya** |  |  |  |  |  |
| The intervention package without TPT compared to status quo | -30.3%<br>(-33.1% to -29%) | -45.1%<br>(-46.8% to -43%) | -41.7%<br>(-43.2% to -39.8%) | 49.6%<br>(-14% to 172.6%) | -38.4%<br>(-42.5% to -30.1%) |
| The intervention package with TPT compared to the intervention package without TPT | -19.2%<br>(-20.1% to -17.6%) | -12.2%<br>(-12.9% to -11.1%) | -11.7%<br>(-12.4% to -10.7%) | -11.1%<br>(-23% to 6.3%) | -12.1%<br>(-15.2% to -9.4%) |
| **South Africa** |  |  |  |  |  |
| The intervention package without TPT compared to status quo | -22.7%<br>(-27.2% to -19.4%) | -35.5%<br>(-38.1% to -30%) | -33.5%<br>(-36.6% to -26.5%) | 154.9%<br>(-31.5% to 534.4%) | -34.3%<br>(-38.5% to -25%) |
| The intervention package with TPT compared to the intervention package without TPT | -13.1%<br>(-14.4% to -11.2%) | -6.4%<br>(-7.9% to -4.8%) | -7.5%<br>(-8.5% to -6.3%) | -9%<br>(-24.8% to 11.2%) | -7.9%<br>(-9.5% to -6.3%) |

Notes: Value represent mean (95% UR). All costs are in 2023 USD. No discounting.
Abbreviations: TPT, tuberculosis preventive treatment; TB, tuberculosis; DALYs, disability adjusted life years.

The intervention package is projected to substantially reduce tuberculosis incidence and mortality in all countries (**Figure 2, Appendix** p41). The incremental per capita expenditure from the health system perspective required to achieve these impacts varied over time and between countries, typically increasing during the scale-up period from 2024 to 2030 before decreasing in each country to below $0.50 per capita by 2050 (**Figure 3, Panel A**). These increases in health system spending, however, were more than offset by the societal return (**Figure 3, Panel B**). Societal returns equivalent to the amount invested by the health system are projected to be achieved in all countries by 2027, and by 2050, the societal ROI is projected to be $10.80, $3.70, $27.40, and $39.00 for Brazil, Georgia, Kenya, and South Africa, respectively (**Table 6**).

**Figure 2.**
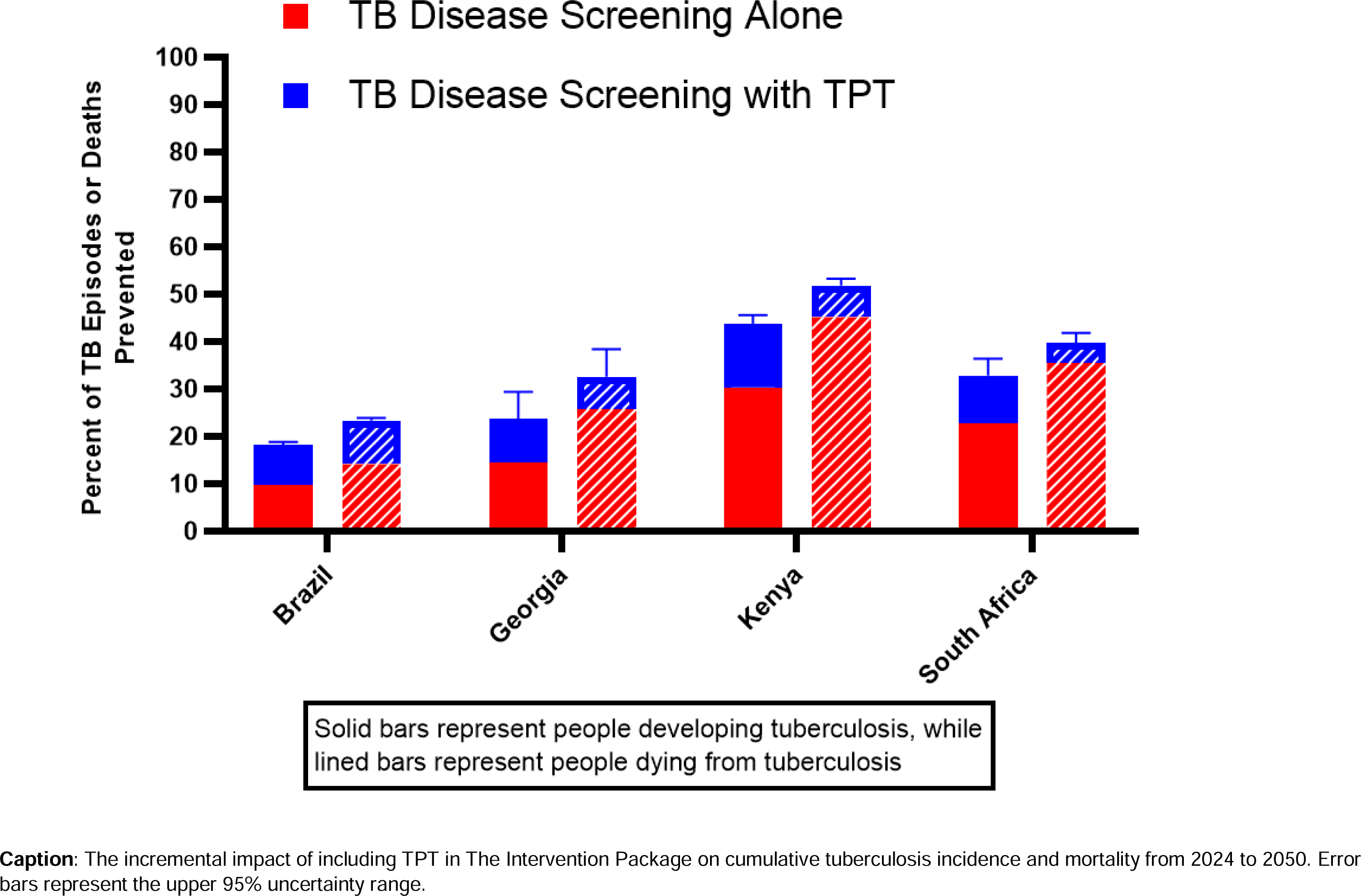
Impact of The Intervention Package without and with tuberculosis preventive treatment on cases and deaths by 2050.

**Figure 3.**
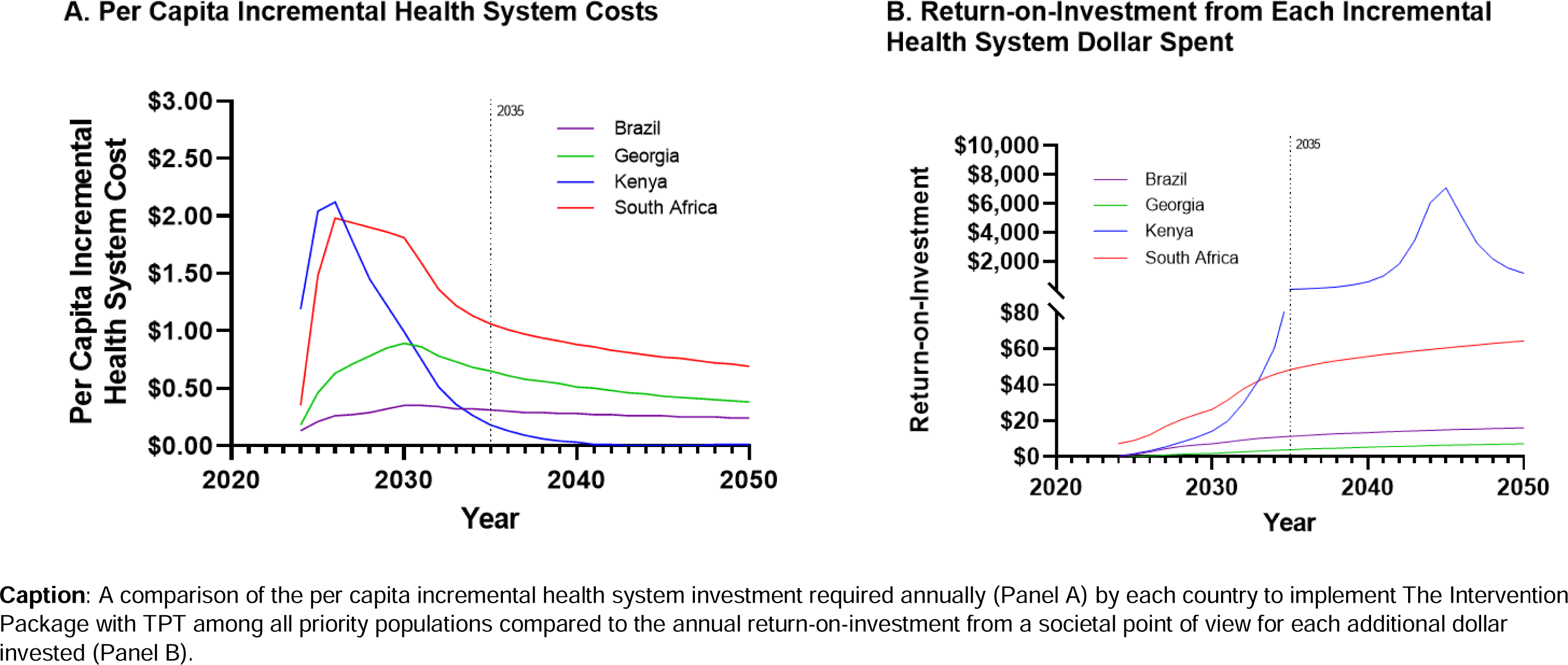
Incremental Per Capita Health System Investment vs. Return-on-Investment for Each Dollar Spent.

**Table 6.** Cost-Effectiveness and Return on Investment by 2050.

| Outcome | Brazil | Georgia | Kenya | South Africa |
| --- | --- | --- | --- | --- |
| **Intervention package without TPT vs. status quo** |  |  |  |  |
| Societal Return on Investment | \$8.00 | \$5.40 | \$13.20 | \$25.80 |
| Incremental Health System Cost per DALY averted | \$1078 | \$1118 | \$152 | \$249 |
| Incremental Health System Cost per TB case averted | \$5110 | \$5932 | \$1365 | \$2394 |
| Incremental Health System Cost per TB death averted | \$41,100 | \$48,080 | \$6316 | \$9550 |
| Incremental Societal Cost per DALY averted | Cost Saving | Cost Saving | Cost Saving | Cost Saving |
| Incremental Societal Cost per TB case averted | Cost Saving | Cost Saving | Cost Saving | Cost Saving |
| Incremental Societal Cost per TB death averted | Cost Saving | Cost Saving | Cost Saving | Cost Saving |
| **Intervention package with TPT vs. intervention package without TPT** |  |  |  |  |
| Societal Return on Investment | \$25.20 | \$0.80 | Both Health System and Societal Savings | Both Health System and Societal Savings |
| Incremental Health System Cost per DALY averted | \$315 | \$2475 | Cost Saving | Cost Saving |
| Incremental Health System Cost per TB case averted | \$1118 | \$5494 | Cost Saving | Cost Saving |
| Incremental Health System Cost per TB death averted | \$12,481 | \$109,873 | Cost Saving | Cost Saving |
| Incremental Societal Cost per DALY averted | Cost Saving | \$417 | Cost Saving | Cost Saving |
| Incremental Societal Cost per TB case averted | Cost Saving | \$926 | Cost Saving | Cost Saving |
| Incremental Societal Cost per TB death averted | Cost Saving | \$18,522 | Cost Saving | Cost Saving |
| **Intervention package with TPT vs. status quo** |  |  |  |  |
| Societal Return on Investment | \$10.80 | \$3.70 | \$27.40 | \$39.00 |
| Incremental Health System Cost per DALY averted | \$771 | \$1402 | \$72 | \$163 |
| Incremental Health System Cost per TB case averted | \$3219 | \$5762 | \$521 | \$1240 |
| Incremental Health System Cost per TB death averted | \$29,838 | \$60,693 | \$3025 | \$6415 |
| Incremental Societal Cost per DALY averted | Cost Saving | Cost Saving | Cost Saving | Cost Saving |
| Incremental Societal Cost per TB case averted | Cost Saving | Cost Saving | Cost Saving | Cost Saving |
| Incremental Societal Cost per TB death averted | Cost Saving | Cost Saving | Cost Saving | Cost Saving |

Trends among priority groups and how they vary are in the **Appendix** (pp42-48). The incremental decrease in tuberculosis incidence seen by 2050 by implementing active case finding from 2024-2026 among high-risk communities was <1% in all countries (**Appendix** p45 and p48). Most of the reduction in tuberculosis incidence in these communities is instead driven by screening and prevention efforts among people with HIV and household contacts.

### Projections when implementing the intervention package without TPT

The implementation of systematic tuberculosis disease screening without TPT in priority populations is projected to reduce tuberculosis incidence by 9.6% (95%UR 9.3% to 10%) in Brazil, 14.4% (11% to 19.6%) in Georgia, 30.3% (29% to 33.1%) in Kenya, and 22.7% (19.4% to 27.2%) in South Africa by 2050, with even greater reductions in mortality (**Figure 2**, **Table 5**). The implementation of such activities is associated with increased health system costs in all countries, but comparatively larger savings from a societal perspective, which range from $160 million (95%UR $80 to $300 million) in Georgia to $58.8 billion (95% UR $21.8 to $93.7 billion) in South Africa by 2050 (**Appendix** p39). These large differences in health system expenditures and societal savings correspond to societal ROIs of $8.00, $5.40, $13.20, and $25.80 per dollar invested in tuberculosis disease screening in Brazil, Georgia, Kenya, and South Africa, respectively (**Table 6**).

Overall, tuberculosis disease screening alone is projected to avert 1.2 million (1.1 to 1.4 million) DALYs in Brazil, 33,000 (21,000 to 55,000) DALYs in Georgia, 8.5 million (7.6 to 11.1 million) DALYs in Kenya, and 9.5 million (4.1 to 14.8 million) DALYs in South Africa. These are equivalent to health system costs per DALY averted of $1078, $1118, $152, and $249, in Brazil, Georgia, Kenya, and South Africa, respectively—all well below country-specific willingness-to-pay thresholds. The intervention is cost-saving from a societal perspective in all countries (**Table 6**).

### Projections of the incremental cost and impact of including TPT

When compared to tuberculosis disease screening alone, implementing TPT resulted in large incremental increases in the proportion of tuberculosis prevented. TPT is projected to prevent an additional 9.5% (95% UR 9.3% to 9.8%) of tuberculosis in Brazil, 10.9% (9.8% to 12.3%) in Georgia, 19.2% (17.6% to 20.1%) in Kenya, and 13.1% (11.2% to 14.4%) in South Africa (**Figure 2**, **Table 5**). Paradoxically, the systematic implementation of TPT is also projected to increase costs to patients and families due to the comparatively large number of people receiving TPT per additional case of tuberculosis prevented (**Appendix** p39).

In both Brazil and Georgia, the additional implementation of TPT is associated with modest incremental increases in health system costs, while it is projected to potentially be cost-saving from health system perspective in both Kenya (savings of $0.6 billion, 95% UR savings of $1.9 billion to additional costs of $0.2 billion) and South Africa (savings of $0.6 billion, 95% UR savings of $2.2 billion to additional costs of $0.3 billion). In Brazil, Kenya, and South Africa, TPT is projected to result in significant societal savings, while in Georgia, it is projected to be essentially cost neutral (societal cost of $3.7 million, 95% UR savings of $40 million to additional costs of $70 million). The societal ROI of adding TPT is projected to be $25.20 in Brazil and $0.80 in Georgia **(Table 6).** In Kenya and South Africa, the addition of TPT is projected to become cost-saving from a health system perspective by the year 2031.

The use of TPT is projected to significantly reduce the number of DALYs in all countries. When compared to tuberculosis disease screening alone, TPT is projected to avert an additional 815,000 (95%UR 756,000 to 903,000) DALYs in Brazil, 8800 (6500 to 10,400) DALYs in Georgia, 1.4 million (1.3 million to 1.6 million) DALYs in Kenya, and 1.4 million (815,000 to 1.7 million) DALYs in South Africa (**Appendix** p39). TPT is cost-saving from a health system perspective in Kenya and South Africa and cost-effective in Brazil ($315 per DALY averted), however did not reach the willingness-to-pay threshold per DALY averted in Georgia ($2475 per DALY averted). From a societal perspective, the cost per DALY averted in Georgia is $417 (cost-effective); in all other countries TPT was cost-saving from a societal perspective (**Table 6**).

### Scenario Analysis

Overall conclusions in our primary analysis did not change when considering differing discount rates (**Appendix** pp49-52). When considering rates specific to country-level income (the largest rates), the societal ROI of the intervention package was $9.70 in Brazil, $3.10 in Georgia, $17.20 in Kenya, and $34.20 in South Africa. Findings across all other scenario analyses were also consistent with our conclusions surrounding the cost-effectiveness and societal ROI of implementing the intervention package with TPT (**Appendix** pp49-68), with the one exception being when evaluating outcomes from 2024 to 2035, the intervention package was no longer cost-effective from a health system perspective in Georgia when implemented with TPT (**Appendix** p58).

## Discussion

In this evaluation, we found scaling-up a package of interventions to improve tuberculosis screening and TPT in four countries with distinct epidemiological profiles could prevent many people from developing tuberculosis, while being cost-saving from a societal perspective in all countries. The societal ROI per incremental health system dollar spent ranged from $4 in Georgia to $39 in South Africa.

The mechanisms and populations by which reductions in tuberculosis incidence were obtained varied by country epidemiology and current levels of screening and TPT coverage. In Brazil, Kenya, and South Africa, a large proportion of people developing tuberculosis have HIV, and in these countries, scaling-up screening and TPT among people with HIV is associated with large reductions in tuberculosis incidence. However, in Georgia, where HIV co-infection comparatively uncommon, focusing efforts on this group is projected to have little epidemiological impact; much more impact is projected instead when focusing screening and TPT on HHC. Moreover, in Georgia, rifampicin-resistance is common and while TPT is projected to still avert many episodes of tuberculosis, it is expected to increase societal costs.

Active case finding in high-tuberculosis prevalence communities (>0.5% tuberculosis prevalence) has been recommended by WHO,^9^ driven by evidence of reductions in prevalence and transmission seen in a recent randomized controlled trial.^27^ However, over the long-term, we found short-term intensive case-finding activities did not result in sustained reductions in tuberculosis incidence. Similar findings were seen among people deprived of liberty in Brazil.^28^ This could be due to the duration of case-finding activities in our model (3 years) and insufficient reductions in tuberculosis prevalence to limit epidemic recrudescence. Indeed, while there were short term reductions, they quickly dissipated in each country among high-risk communities once case-finding activities stopped. In stark contrast, we found the implementation of TPT—even in very high tuberculosis incidence countries—was associated with large incremental reductions in tuberculosis disease incidence over the long-term. A major criticism of scaling up TPT in high tuberculosis incidence settings is that ongoing transmission will offset gains from TPT. However, we found when TPT is coupled with systematic tuberculosis disease screening it can be highly impactful and cost-saving from both health system and societal perspectives. This is true even though we allowed TPT to be provided only once and considered DALY decrements associated with TPT-related adverse events and patient costs of TPT—two important consequences of TPT infrequently considered.

The societal ROI estimated in this study for the scale-up of tuberculosis screening and TPT are large and in line with other ROI estimates for tuberculosis when considered in context. The Copenhagen Consensus estimated $46 USD would be returned for each $1 invested in tuberculosis when six major activities were conducted.^29^ Similarly, a recent WHO-commissioned investment case suggested a return of $7 USD for each $1 invested in developing and implementing a new tuberculosis vaccine over a 25-year horizon.^30^ We have shown that by using tools available now, significant returns can be achieved in countries of varying tuberculosis epidemiology, absent other (no less important) advances, emphasizing the importance of investing in improving tuberculosis prevention and care now.

Though we project substantial reductions in country-level tuberculosis incidence and mortality with the intervention package among people with HIV, HHC, and high-risk communities, this is insufficient to eliminate tuberculosis (annual incidence <1 per million) by 2050. To eliminate tuberculosis, efforts need to go beyond these groups, as recognized by several low-incidence countries and the WHO.^31^ For the countries modeled, elimination will require annual tuberculosis incidence reductions of 22-27% from 2024 to 2050.^32^ However, with the intervention package, we project much more modest annual reductions ranging from 2.5% in Brazil to 4.5% in South Africa. Yet, these advances towards elimination are still impactful, saving millions of years of life and averting substantial morbidity and should not be discounted.^33^ Our results provide strong grounds to support the scale-up of both tuberculosis case finding and associated TPT among people with HIV and HHC, with further research needed to understand the optimal screening and prevention strategies in high-risk communities.

Our study has several strengths. We worked closely with national tuberculosis programmes in each country to develop the intervention package, with a view to developing a feasible-to-implement intervention, and worked together to parameterize model and cost data and interpret results. We developed robust, country-specific models, specifically calibrated to tuberculosis epidemiology in each country. We developed common cost models for each country while incorporating country-specific cost data considering elements of implementation and scale-up. Our model considered age-specific consequences associated with serious adverse events during tuberculosis disease treatment and TPT, as well as increased mortality and long-term morbidity among tuberculosis survivors, two factors often overlooked.

This study should be interpreted in the context of its limitations. We did not consider DALYs or costs associated with future morbidity and care that would now be incurred due to averted tuberculosis mortality. While important, our aim was to evaluate the direct impacts of our intervention on tuberculosis disease. We did not consider patient or family costs among children, as these are uncertain, particularly the indirect costs. Our model was age-structured in four strata for efficiency, which may miss important differences in transmission dynamics, particularly for the large age group of ≥15y. Similarly, we modeled three populations in people with HIV, high-risk communities, and the general population (household contacts could emerge from any of these three). This will miss disparities in risk that exist within the general population (e.g., among those with immunocompromising conditions) and across all strata (e.g., socioeconomic status). We estimated lost productivity due to premature mortality to be equivalent to country-level per-capita GDP. This is a conservative estimate when compared to other methods (e.g., value of a statistical life), and so may have underestimated societal costs and cost savings.^34^ Finally, it is not possible to predict all costs associated with implementation, so we may have underestimated health system costs. However, the large societal gains seen in all countries suggest we would have had to under-estimate health system expenditures on the order of several billion USD to render interventions not cost-saving from a societal perspective.

In summary, we found a comprehensive package of feasible-to-implement interventions related to tuberculosis screening and TPT targeted to people with HIV, HHC, and high-risk communities was associated with significant reductions in tuberculosis incidence and mortality, and societal ROI ranging from $4 to $39 USD, in Brazil, Georgia, Kenya, and South Africa—four epidemiologically distinct countries. These data can be used to advocate for and motivate scale-up of tuberculosis screening and prevention activities globally, and demonstrate the significant societal value of improving tuberculosis prevention and care, even with currently available technologies.

## Acknowledgments

We are grateful for Lindiwe Mvusi, Yolisa Tsibolane, Jason Andrews, José Nildo de Barros Silva, Luiz Henrique Arroyo, Farley Liliana Romera Vega, Danielle Gomes Dell’Orti, Monica Rondon Cotacio, Irma Khonelidze, Aiban Ronoh, Wesley Tomno, Silan Kamuren, Simion Ndemo, S.K. Macharia, Auma Perez, Nkateko Mkhondo, Eunice Omesa, Kleydson Alves, Pedro Avedillo, Nestor Vera Nieto, Monica Rondon Cotacio, Andrei Dadu, Nino Mamulahvili, and Placide Nsengiyumva who facilitated this work and provided additional information key to developing the transmission and economic models.

## Contributions

Initial Conceptualization: JRC, SDB, JFV, KS, NA, DF

Study Development: All Authors

Data Curation: All Authors

Formal Analysis: JRC, JFV, MS

Funding Acquisition: JRC

Methodology: JRC, JFV, KS, NA, CM, IGB, SDB, DF

Project Administration and Supervision: JRC

Visualization: JFV, JRC

Writing (original draft): JRC

Writing (reviewing and editing): All authors.

## Data Sharing

All data is available from the authors upon request. All code in support of TB transmission models can be found in the following online repository https://github.com/juanvesga/return-on-investment-of-TB-screening-and-prevention

