## Appendix for "The Return on Investment of Scaling Tuberculosis Screening and Preventive Treatment: A Modelling Study in Brazil, Georgia, Kenya, and South Africa"

### Additional Methods

#### Defining high-risk populations in each country

**Brazil**: We used data provided by the national tuberculosis programme in Brazil that reported the estimated population size in prisons in 2022 (~650,000) and the reported number of people developing TB in prisons that same year (7290) to initially parameterize TB epidemiology in prisons (i.e., incidence of 1.1%). Based on data provided by the NTP, length of stay in prison was estimated to be an average of 2 years. We used these data to calibrate tuberculosis epidemiology among people deprived of liberty in Brazil (assumed all were at least 15 years of age).

**Georgia**: We used data provided by the national tuberculosis programme in Georgia collected through active case finding activities in priority populations in 2021 and 2022, where people who are accessing care for injection drug use had a tuberculosis prevalence of 0.9% based on 21 cases being detected among 2192 screened. Data provided by the national tuberculosis programme estimated the size of the overall population accessing care to be 17,500 in Georgia (assumed all were at least 15 years of age).

**Kenya**: We worked with the national tuberculosis programme in Kenya to analyse notification data by slums in Mombasa and Nairobi as well as refugee camps in Turakana and Garisa. We used the estimated prevalence:notification ratio of 1.75 to adjust for under-notification of tuberculosis in the populations in these settings. As people living with HIV in these populations are targeted by other interventions in our model and we only target those at least 15 years of age, we further adjusted the population size to only include those living without HIV and those at least 15 years of age based on reported HIV prevalence and TB/HIV incidence in each area, number of individuals under 15 years of age (38.4%), and proportion of tuberculosis occurring in those under 15 years of age (9%). The overall population size was 416,000 and the estimated prevalence was 744 per 100,000.

**South Africa**: We followed similar methods to Kenya. We worked with the national tuberculosis programme in South Africa to analyse tuberculosis notification data in each subdistrict, stratified by HIV. Within each subdistrict, we estimated overall prevalence of tuberculosis based on an estimated prevalence:notification ratio of 1.75. Using data on HIV prevalence in each province, and assuming homogeneity by subdistrict, plus data on the proportion of all tuberculosis occurring among people living with HIV, we estimated the prevalence of tuberculosis among people living without HIV (as they are the focus of other interventions). Finally, as the intervention only targets those at least 15 years of age, we adjusted population size based on the proportion <15 years of age (28.7%) and the proportion of tuberculosis occurring among those under 15 years of age (7%). We then arrived at a population size of 1.1 million and tuberculosis prevalence of 1900 per 100,000.

#### Country status quo algorithms

<table>
<tr><th>Population</th><th>Age Group (years)</th><th>Brazil</th><th>Georgia</th><th>Kenya</th><th>South Africa</th></tr>
<tr><td rowspan="4">People living with HIV</td><td>0-4</td><td rowspan="4">Symptom screening as a triage for chest x-ray and Xpert testing. Repeat Xpert testing if chest x-ray abnormalities suggest tuberculosis. Testing for tuberculosis infection with IGRA; 3HP and 6H used for TPT.</td><td rowspan="2">Symptoms as a triage for Xpert testing. 3HP and 6H used for TPT.</td><td rowspan="4">Symptoms as a triage for Xpert testing; if both negative, then chest x-ray performed to definitively rule out tuberculosis. 3HR and 6H used for TPT.</td><td rowspan="4">Systematic symptoms and Xpert testing. No tuberculosis infection testing; 3HP and 6H used for TPT.</td></tr>
<tr><td>5-9</td></tr>
<tr><td>10-14</td><td rowspan="2">Symptoms, CRP, and chest x-ray done in parallel; if any positive, Xpert testing. 3HP and 6H used for TPT.</td></tr>
<tr><td>≥15</td></tr>
<tr><td rowspan="4">Household Contacts</td><td>0-4</td><td rowspan="4">Symptom screening and tuberculin skin test performed in parallel; if either positive, a chest x-ray is performed. If chest x-ray abnormalities, Xpert testing done; if normal chest x-ray, TPT prescribed (3HP or 6H).</td><td rowspan="4">Symptom screening and chest x-ray performed in parallel; if symptoms present or chest x-ray abnormalities, Xpert testing. If testing for tuberculosis infection is done, typically with tuberculin skin test. 3HP and 6H used for TPT.</td><td rowspan="4">Symptom screening as a triage for chest x-ray. If chest x-ray abnormalities, then Xpert testing. Tuberculosis infection done when tests are available; 3HR and 6H used for TPT.</td><td rowspan="4">Systematic symptoms and Xpert testing. Tuberculin skin test used, when available for select age groups; 3HP and 6H used for TPT when indicated.</td></tr>
<tr><td>5-9</td></tr>
<tr><td>10-14</td></tr>
<tr><td>≥15</td></tr>
<tr><td rowspan="4">All Other Populations</td><td>0-4</td><td rowspan="4">Symptom screening as a triage for sputum smear microscopy. Typically, no TPT.</td><td rowspan="3">Symptom screening as a triage for Xpert. Typically, no TPT.</td><td rowspan="4">Symptom screening as a triage for sputum smear microscopy. Typically, no TPT.</td><td rowspan="4">Symptoms as a triage for Xpert. Typically, no TPT.</td></tr>
<tr><td>5-9</td></tr>
<tr><td>10-14</td></tr>
<tr><td>≥15</td><td>Symptom screening as a triage for sputum smear microscopy. Typically, no TPT.</td></tr>
</table>

Abbreviations: IGRA, interferon-gamma release assay; TPT, tuberculosis preventive treatment; CRP, C-reactive protein; HIV, human immunodeficiency virus; 3HP, three-months of once-weekly isoniazid and rifapentine; 6H, six-months of daily isoniazid; 3HR, three-months of daily rifampicin and isoniazid

#### Diagnostic Parameters Used to Estimate Diagnostic Performance

| Test | Sensitivity (95% CI) | Specificity (95% CI) | Reference |
|---|---|---|---|
| Symptom screen (child) | 89 (52-98) | 69 (51-83) | 1 |
| Symptom Screen (non-HIV) | 71 (62-79) | 64 (52-74) | 2 |
| Symptom screen (HIV, no ART) | 84 (75-90) | 37 (25-50) | 2 |
| Symptom screen (HIV, ART) | 53 (36-69) | 70 (50-85) | 2 |
| Xpert (child) | 73 (65-80) | 97 (96-98) | 4 |
| Xpert (adult) | 90 (84-94) | 96 (93-97) | 4 |
| Xpert (HIV) | 88 (75-94) | 93 (82-97) | 5 |
| Xpert (child, screening) | 50 (35-62) | 99 (97-99) | * |
| Xpert (adult, screening) | 69 (48-86) | 99 (97-99) | 2 |
| Xpert (HIV, screening) | 69 (60-76) | 98 (97-99) | 2 |
| CXR (child) | 84 (70-92) | 91 (90-92) | 2 |
| CXR (no HIV) | 85 (77-90) | 96 (93-97) | 2 |
| CXR (HIV) | 63 (56-70) | 78 (66-86) | 3 |
| CRP (HIV, no ART) | 89 (85-92) | 54 (45-62) | 2 |
| CRP (HIV, on ART) | 40 (10-80) | 80 (75-84) | 2 |
| Smear (child) | 26 (14-39) | 100 (99-100) | 6 |
| Smear (no HIV) | 80 (60-90) | 99 (98-100) | 7, 8, 9 |
| Smear (HIV) | 64 (56-72) | 99 (94-100) | 10 |
| TST (no BCG) | 77 (72-81) | 97 (95-99) | 11 |
| TST (BCG) | 77 (72-81) | 59 (46-73) | 11 |
| TST (HIV) | 60 (34-82) | 97 (95-99) | 12 |
| IGRA (no HIV) | 78 (73-81) | 98 (96-99) | 11 |
| IGRA (HIV) | 69 (50-84) | 98 (96-99) | 13 |
| TBST (no HIV) | 78 (73-81) | 98 (96-99) | 14 |
| TBST (HIV) | 69 (50-84) | 98 (96-99) | 14 |

Abbreviations: HIV, human immunodeficiency virus; CXR, chest x-ray; CRP, C-reactive protein; ART, antiretroviral treatment; BCG, Bacille Calmette-Guerin; TST, tuberculin skin test; IGRA, interferon-gamma release assay; TBST, tuberculosis-based skin test (antigen based).
*Inferred from difference in sensitivity between child and adults in a diagnostic setting; specificity assumed the same. Confidence intervals generated based on proportional difference in LCI and UCI from point estimate in adults

#### Country-Specific Diagnostic Algorithms and Their Performance Characteristics

| Country | Group | Age | Algorithm | Sensitivity | Specificity |
|---|---|---|---|---|---|
| Kenya | PLHIV | <5y<br>5-9y<br>10-14y<br>≥15y | Symptoms-->Xpert-->CXR-->Xpert* | ART = 49.85 (33.63 to 66.4)<br>No ART = 79.18 (68.71 to 87.17) | ART = 97.45 (92.82 to 99.68)<br>No ART = 94.67 (85.96 to 99.2) |
| | HHC | <5y<br>5-9y<br>10-14y | Symptoms-->CXR-->Xpert | 54.61 (31.3 to 69.13) | 99.92 (99.86 to 99.96) |
| | | ≥15y | Symptoms-->CXR-->Xpert | 54.46 (45.58 to 62.2) | 99.94 (99.86 to 99.98) |
| | Other Groups | <15y | Symptoms-->Smear | 22.94 (10.37 to 37.51) | 99.95 (99.54 to 100) |
| | | ≥15y | Symptoms-->Smear | 56.93 (42.35 to 70.26) | 99.64 (99.22 to 99.88) |
| South Africa | PLHIV | <5y<br>5-9y<br>10-14y<br>≥15y | Symptoms + Xpert | 69 (60.03 to 76.96) | 98.00 (97.02 to 98.85) |
| | HHC | <5y<br>5-9y<br>10-14y | Symptoms + Xpert | 49.57 (34.67 to 64.22) | 99.01 (96.92 to 99.92) |
| | | ≥15y | Symptoms + Xpert | 69.91 (50.28 to 86.6) | 98.96 (96.77 to 99.92) |
| | Other Groups | <15y | Symptoms-->Xpert | 64.96 (38.65 to 79) | 99.07 (98.46 to 99.56) |
| | | ≥15y | Symptoms-->Xpert | 63.96 (54.88 to 72.05) | 98.56 (97.28 to 99.42) |
| Georgia | PLHIV | <5y<br>5-9y | Symptoms-->Xpert | ART = 46.45 (31.11 to 62.62)<br>No ART = 73.78 (60.87 to 84.08) | ART = 97.87 (93.9 to 99.74)<br>No ART = 95.55 (87.87 to 99.33) |
| | | 10-14y<br>≥15y | CRP+Symptoms+CXR-->Xpert | ART = 78.82 (64.16 to 89.3)<br>No ART = 87.41 (73.30 to 96.07) | ART = 96.04 (89.63 to 99.41)<br>No ART = 94.06 (84.27 to 99.13) |
| | HHC | <5y<br>5-9y<br>10-14y | Symptoms+CXR-->Xpert | 71.81 (63.06 to 80.06) | 98.88 (98.27 to 99.37) |
| | | ≥15y | Symptoms+CXR-->Xpert | 86.13 (79.9 to 90.94) | 98.46 (97.16 to 99.37) |
| | Other Groups | <15y | Symptoms-->Xpert | 64.96 (38.65 to 79) | 99.07 (98.46 to 99.56) |
| | | ≥15y | Symptoms-->Smear | 56.93 (42.35 to 70.26) | 99.64 (99.22 to 99.88) |
| Brazil | PLHIV | <5y<br>5-9y<br>10-14y<br>≥15y | Symptoms-->CXR+Xpert-->Xpert** | ART = 49.85 (33.63 to 66.4)<br>No ART = 79.18 (68.71 to 87.17) | ART = 97.45 (92.82 to 99.68)<br>No ART = 94.67 (85.96 to 99.20) |
| | HHC | <5y<br>5-9y<br>10-14y | Symptoms+TST-->CXR-->Xpert | 59.89 (47.69 to 70.59) | 99.84 (99.73 to 99.89) |
| | | 15+y | Symptoms+TST-->CXR-->Xpert | 71.53 (63.67 to 78.31) | 99.90 (99.78 to 99.97) |
| | Other Groups | <15y | Symptoms-->Smear | 22.94 (10.37 to 37.51) | 99.95 (99.54 to 100) |
| | | ≥15y | Symptoms-->Smear | 56.93 (42.35 to 70.26) | 99.64 (99.22 to 99.88) |

Notes: Tests are done in parallel when connected with a "+" and done sequentially when denoted with "-->". Sequential tests are only performed if the previous test (or at least one of the previous tests if done in parallel) are positive, except where noted with an asterisk.
*The CXR is only performed after Xpert if the initial Xpert is NEGATIVE. If the CXR suggests abnormalities, the Xpert is repeated.
**The Xpert is only repeated if CXR suggests abnormalities but the initial Xpert is NEGATIVE.
Abbreviations: PLHIV, people living with HIV; HHC, household contacts; CXR, chest x-ray; TST, tuberculin skin test; CRP, C-reactive protein; y, years.

#### Performance of The Intervention Package and the Enhanced Package's Diagnostic Algorithms

| | | | Sensitivity (95% UR) | Specificity (95% UR) | TB Infection Test Used? | Algorithm Details |
|---|---|---|---|---|---|---|
| PLHIV | <5y | The Intervention Package | No ART = 73.78 (60.87 to 84.04)<br>ART = 46.45 (31.11 to 62.62) | No ART = 95.55 (87.87 to 99.33)<br>ART = 97.87 (93.9 to 99.74) | No | No ART = W4SS as triage for Xpert<br>ART = W4SS as triage for Xpert |
| | | Enhanced Package | No ART = 73.78 (60.87 to 84.04)<br>ART = 46.45 (31.11 to 62.62) | No ART = 95.55 (87.87 to 99.33)<br>ART = 97.87 (93.9 to 99.74) | No | No ART = W4SS as triage for Xpert<br>ART = W4SS as triage for Xpert |
| | 5-9y | The Intervention Package | No ART = 73.78 (60.87 to 84.04)<br>ART = 46.45 (31.11 to 62.62) | No ART = 95.55 (87.87 to 99.33)<br>ART = 97.87 (93.9 to 99.74) | No | No ART = W4SS as triage for Xpert<br>ART = W4SS as triage for Xpert |
| | | Enhanced Package | No ART = 82.74 (69.21 to 91.48)<br>ART = 46.45 (31.11 to 62.62) | No ART = 94.99 (86.72 to 99.24)<br>ART = 97.87 (93.9 to 99.74) | No | No ART = W4SS + CXR (parallel) as triage for Xpert<br>ART = W4SS as triage for Xpert |
| | 10-14y | The Intervention Package | No ART = 65.65 (53.69 to 75.57)<br>ART = 46.45 (31.11 to 62.62) | No ART = 97.94 (94.42 to 99.71)<br>ART = 97.87 (93.9 to 99.74) | No | No ART = W4SS as triage for CRP as triage for Xpert<br>ART = W4SS as triage for Xpert |
| | | Enhanced Package | No ART = 65.65 (53.69 to 75.57)<br>ART = 72.65 (60.12 to 82.65) | No ART = 97.94 (94.42 to 99.71)<br>ART = 96.8 (91.49 to 99.56) | No | No ART = W4SS as triage for CRP as triage for Xpert<br>ART = W4SS + CXR (parallel) as triage for Xpert |
| | ≥15y | The Intervention Package | No ART = 65.65 (53.69 to 75.57)<br>ART = 46.45 (31.11 to 62.62) | No ART = 97.94 (94.42 to 99.71)<br>ART = 97.87 (93.9 to 99.74) | No | No ART = W4SS as triage for CRP as triage for Xpert<br>ART = W4SS as triage for Xpert |
| | | Enhanced Package | No ART = 65.65 (53.69 to 75.57)<br>ART = 72.65 (60.12 to 82.65) | No ART = 97.94 (94.42 to 99.71)<br>ART = 96.8 (91.49 to 99.56) | No | No ART = W4SS as triage for CRP as triage for Xpert<br>ART = W4SS + CXR (parallel) as triage for Xpert |
| HHC | <5y | The Intervention Package | 71.81 (63.06 to 80.16) | 98.88 (98.27 to 99.37) | No | W4SS + CXR (parallel) as triage for Xpert |
| | | Enhanced Package | 71.81 (63.06 to 80.16) | 98.88 (98.27 to 99.37) | No | W4SS + CXR (parallel) as triage for Xpert |
| | 5-9y | The Intervention Package | 71.81 (63.06 to 80.16) | 98.88 (98.27 to 99.37) | TST | W4SS + CXR (parallel) as triage for Xpert |
| | | Enhanced Package | 71.81 (63.06 to 80.16) | 98.88 (98.27 to 99.37) | TBST | W4SS + CXR (parallel) as triage for Xpert |
| | 10-14y | The Intervention Package | 71.81 (63.06 to 80.16) | 98.88 (98.27 to 99.37) | TST | W4SS + CXR (parallel) as triage for Xpert |
| | | Enhanced Package | 71.81 (63.06 to 80.16) | 98.88 (98.27 to 99.37) | TBST | W4SS + CXR (parallel) as triage for Xpert |
| | ≥15y | The Intervention Package | 86.13 (79.9 to 90.94) | 98.46 (97.16 to 99.37) | TST | W4SS + CXR (parallel) as triage for Xpert |
| | | Enhanced Package | 86.13 (79.9 to 90.94) | 98.46 (97.16 to 99.37) | TBST | W4SS + CXR (parallel) as triage for Xpert |
| High Risk | ≥15y | The Intervention Package | 86.13 (79.9 to 90.94) | 98.46 (97.16 to 99.37) | No | W4SS + CXR (parallel) as triage for Xpert |
| | | Enhanced Package | 86.13 (79.9 to 90.94) | 98.46 (97.16 to 99.37) | TBST | W4SS + CXR (parallel) as triage for Xpert |

Abbreviations: PLHIV, people living with HIV; HHC, household contacts; CXR, chest x-ray; TST, tuberculin skin test; TBST, tuberculosis skin test (antigen-based); CRP, C-reactive protein; y, years; W4SS, WHO recommended four symptom screening.

#### Model structures and demographics used to simulate tuberculosis epidemiology in each country

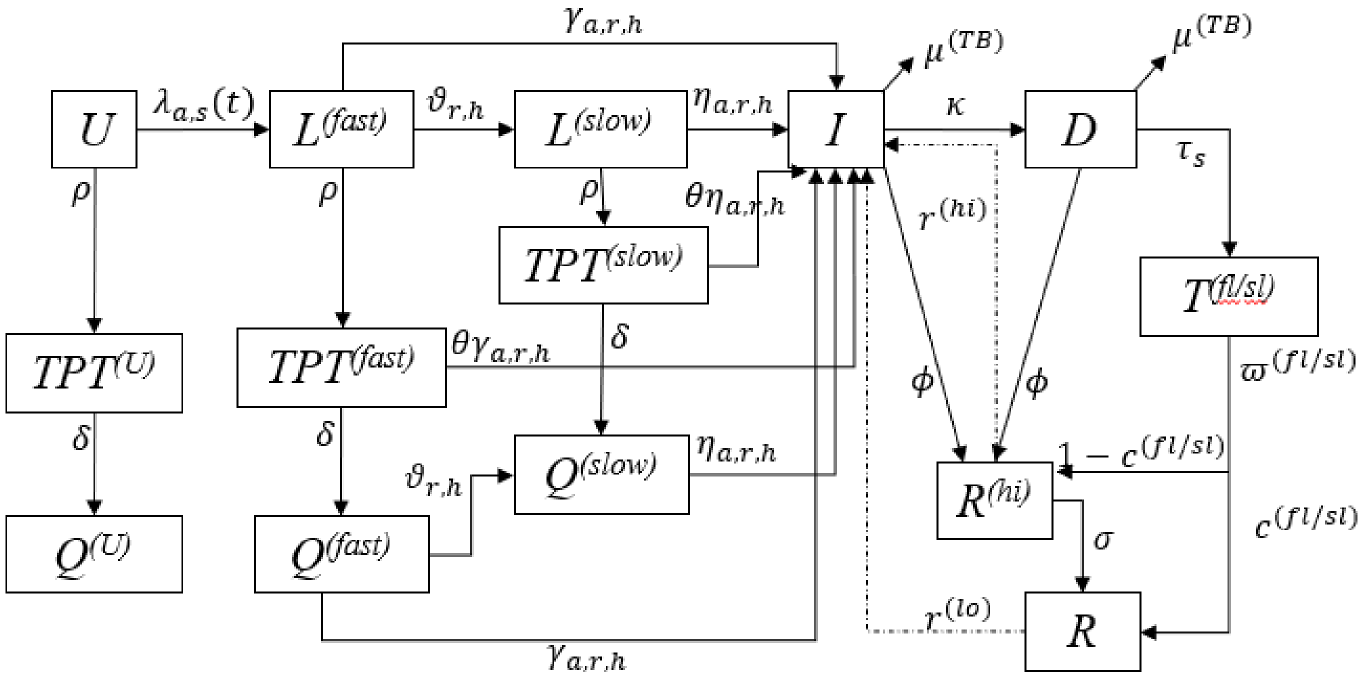

***Fig S1:*** *This schematic describes the natural history and TB –care-cascade designed for this study. Subscripts indicate age (a), strain (s), subpopulation (r), and HIV status (h) dependencies. TB infection occurs with a strain and age-specific force of infection (lambda), leading to TB infection (L(fast)) with a transient elevated risk of progression to active disease (I), or transition to a slow progression TB infected state (L(slow)). Recruitment into TB preventive treatment (TPT) can occur from the uninfected state (U) or the TB infected compartments. Removal from the TPT compartment reflect specific regimen durations, and this has been adjusted to match local guidelines and further interventions. TPT regimen is followed by a post regimen state (Q). Transition into active disease from the TPT compartments occurs at rate reduced according to the specific TPT efficacy (θ). Active TB compartment (I) reflects full symptomatic and infectious TB disease, leading to TB mortality, or self-cure (high relapse recovered - R(hi)) if untreated, or to the "Awaiting diagnosis" (D) compartment at a care-seeking rate (κ). Transition from D to the treatment compartment (T) occur at a rate τ(fl/sl) that combines the probability of testing positive given the testing algorithm and infecting strain, and a treatment coverage parameter. Treated individuals can complete and cure disease (move to low relapse compartment R) or default and move to high relapse R(hi). Treatment duration and completion is specified according to line of treatment (e.g., first or second).*

#### Model equations

Uninfected

$$\frac{dU_{a,h}}{dt} = \begin{cases} b - U_{a,h}(t)[\lambda_{a,s}(t) + \rho_a(t) + \mu_a] + \sum_{i=1}^{4} Z_{i,a}U_{i,h}(t) + W_h^{(1)}(t) & , for\ a = 1 \\ \sum_{i=1}^{4} Z_{i,a}U_{i,h}(t) - U_{a,h}(t)[\lambda_{a,s}(t) + \rho_a(t) + \mu_a] + W_h^{(1)}(t) & , for\ a > 1 \end{cases}$$ (Equation 1)

TPT among uninfected

$$\frac{dTPT_{a,h}^{(U)}}{dt} = U_{a,h}(t)\rho_a(t) - TPT_{a,h}^{(U)}(t)[\delta + \mu_a] + \sum_{i=1}^{4} Z_{i,a}TPT_{i,h}^{(U)}(t) + W_h^{(2)}(t)$$ (Equation 2)

Post-TPT uninfected

$$\frac{dQ_{a,h}^{(U)}}{dt} = TPT_{a,h}^{(U)}(t)\delta - Q_{a,h}^{(U)}[\lambda_{a,s}(t) + \mu_a] + \sum_{i=1}^{4} Z_{i,a}Q_{i,h}^{(U)}(t) + W_h^{(3)}(t)$$ (Equation 3)

TB infection fast progression

$$\frac{dL_{a,s,h}^{(f)}}{dt} = \lambda_{a,s}(t)\left[U_{a,h}(t) + L_{a,s,h}^{(f)}(t) + L_{a,s,h}^{(s)} + R_{a,h}^{(hi)}(t) + R_{a,h}(t)\right] - L_{a,s,h}^{(f)}(t)[\gamma_{a,r,h} + \vartheta_{r,h} + \rho_a(t) + \lambda_{a,s}(t) + \mu_a] + \sum_{i=1}^{4} Z_{i,a}L_{i,s,h}^{(f)}(t) + W_h^{(4)}(t)$$ (Equation 4)

TPT among TB infection fast progressors

$$\frac{dTPT_{a,s,h}^{(f)}}{dt} = L_{a,s,h}^{(f)}(t)\rho_a(t) - TPT_{a,s,h}^{(f)}(t)[\delta + \theta\gamma_{a,r,h} + \mu_a] + \sum_{i=1}^{4} Z_{i,a}TPT_{i,s,h}^{(f)}(t) + W_h^{(5)}(t)$$ (Equation 5)

Post-TPT among latent fast progressors

$$\frac{dQ_{a,s,h}^{(f)}}{dt} = \lambda_{a,s}(t)\left[Q_{a,h}^{(u)}(t) + Q_{a,s,h}^{(f)}(t) + Q_{a,s,h}^{(s)}(t)\right] + TPT_{a,s,h}^{(f)}(t)\delta - Q_{a,s,h}^{(f)}[\lambda_{a,s}(t) + \gamma_{a,r,h} + \vartheta_{r,h} + \mu_a] + \sum_{i=1}^{4} Z_{i,a}Q_{i,s,h}^{(f)}(t) + W_h^{(6)}(t)$$ (Equation 6)

TB infection slow progression

$$\frac{dL_{a,s,h}^{(s)}}{dt} = L_{a,s,h}^{(f)}(t)\vartheta_{r,h} - L_{a,s,h}^{(s)}(t)[\eta_{a,r,h} + \rho_a(t) + \lambda_{a,s}(t) + \mu_a] + \sum_{i=1}^{4} Z_{i,a}L_{i,s,h}^{(s)}(t) + W_h^{(7)}(t)$$ (Equation 7)

TPT among TB infection slow progressors

$$\frac{dTPT_{a,s,h}^{(s)}}{dt} = L_{a,s,h}^{(s)}(t)\rho_a(t) - TPT_{a,s,h}^{(s)}(t)[\delta + \theta\eta_{a,r,h} + \mu_a] + \sum_{i=1}^{4} Z_{i,a}TPT_{i,s,h}^{(s)}(t) + W_h^{(8)}(t)$$ (Equation 8)

Post-TPT among TB infection slow progressors

$$\frac{dQ_{a,s,h}^{(s)}}{dt} = Q_{a,s,h}^{(f)}(t)\vartheta_{r,h} + TPT_{a,s,h}^{(s)}(t)\delta - Q_{a,s,h}^{(s)}[\lambda_{a,s}(t) + \eta_{a,r,h} + \mu_a] + \sum_{i=1}^{4} Z_{i,a}Q_{i,s,h}^{(s)}(t) + W_h^{(9)}(t)$$

(Equation 9)

TB Disease

$$\frac{dI_{a,s}}{dt} = \gamma_{a,r,h}\left[L^{(f)}_{a,s,h}(t) + Q^{(f)}_{a,s,h}(t)\right] + \theta\gamma_{a,r,h}TPT^{(f)}_{a,s,h}(t) + \eta_{a,r,h}\left[L^{(s)}_{a,s,h}(t) + Q^{(s)}_{a,s,h}(t)\right] + \theta\eta_{a,r,h}TPT^{(s)}_{a,s,h}(t) + r^{(lo)}R_{a,h}(t) + r^{(hi)}R^{(hi)}_{a,h} - I_{a,s,h}(t)\left[\kappa_{a,s} + \mu_a + \phi + \mu^{(TB)}\right] + \sum_{i=1}^{4} Z_{i,a}I_{i,s,h}(t) + W^{(10)}_h(t)$$ *(Equation 10)*

TB disease awaiting diagnosis

$$\frac{dD_{a,s,h}}{dt} = I_{a,s,h}(t)\kappa_{a,s} + R^{(hi)}_{a,h}r^{(hi)} + R_{a,h}r^{(lo)} - D_{a,s,h}(t)\left[\tau_s + \phi + \mu_a + \mu^{(TB)}\right] + \sum_{i=1}^{4} Z_{i,a}D_{i,s,h}(t) + W^{(11)}_h(t)$$

*(Equation 11)*

TB disease diagnosed and on first line DS-TB treatment

$$\frac{dT^{(fl)}_{a,s,h}}{dt} = D_{a,s,h}(t)\tau_{ds} - T^{(fl)}_{a,s,h}(t)\left[\varpi^{(fl)} + \mu_a\right] + \sum_{i=1}^{4} Z_{i,a}T^{(fl)}_{i,s,h}(t) + W^{(12)}_h(t)$$ *(Equation 12)*

TB Disease diagnosed and on RR-TB treatment

$$\frac{dT^{(sl)}_{a,s,h}}{dt} = D_{a,s,h}(t)\tau_{dr} - T^{(sl)}_{a,s,h}(t)\left[\varpi^{(sl)} + \mu_a\right] + \sum_{i=1}^{4} Z_{i,a}T^{(sl)}_{i,s,h}(t) + W^{(13)}_h(t)$$ *(Equation 13)*

Recovered with transient high-relapse rates

$$\frac{dR^{(hi)}_{a,h}}{dt} = I_{a,s,h}(t)\phi + D_{a,s,h}(t)\phi + T^{(fl)}_{a,s,h}\varpi^{(fl)}\left(1 - c^{(fl)}\right) + T^{(sl)}_{a,s,h}\varpi^{(sl)}\left(1 - c^{(sl)}\right) - R^{(hi)}_{a,h}(t)\left[\sigma + r^{(hi)} + \lambda_{a,s}(t) + \mu_a\right] + \sum_{i=1}^{4} Z_{i,a}R^{(hi)}_{i,h}(t) + W^{(14)}_h(t)$$ *(Equation 14)*

Recovered with stable relapse rates

$$\frac{dR_{a,h}}{dt} = R^{(hi)}_{a,h}(t)\sigma + T^{(fl)}_{a,s,h}\varpi^{(fl)}c^{(fl)} + T^{(sl)}_{a,s,h}\varpi^{(sl)}c^{(sl)} - R_{a,h}(t)\left[r^{(lo)} + \lambda_{a,s}(t) + \mu_a\right] + \sum_{i=1}^{4} Z_{i,a}R_{ih}(t) + W^{(15)}_h(t)$$ *(Equation 15)*

In equations 1 to 15 the terms in $W^{l}_h(t)$ represent transitions between HIV stages which, for clarity, are listed separately below.

Force of Infection

$$\lambda_{a,s}(t) = \begin{cases} \frac{\beta_{ds}\{\sum_{i=1}^{4} C_{a,i}\sum_{h=1}^{2}[I_{i,s,h}(t)+D_{i,s,h}(t)]\}}{N(t)}, & for\ s = 0 \\ \frac{\beta_{dr}\{\sum_{i=1}^{4} C_{a,i}\sum_{h=1}^{2}[I_{i,s,h}(t)+D_{i,s,h}(t)]\}}{N(t)}, & for\ s = 1 \end{cases}$$ *(Equation 16)*

where *N(t)* represents the total population at time *t*. Sub index *s* denotes the strain of TB (with *s=0* for DS and *s=1* for DR). $C_{a,i}$ reflects the per-capita contact matrix for people of age *a* with those of age *i.*

Transitions through HIV stages

We write $X_h^l(t)$ to represent any one of the compartments in FigS1, with HIV transition dimension *h*, and TB dimension *l*. The following equations represent the dynamics of HIV and ART

*HIV transitions*

$$W_h^l = \begin{cases} -X_h^l(t)h(t) & for\ h = 0 \\ X_0^l(t)h(t) - X_h^l(t)e(t)\Omega_l\Upsilon + \sum_{h=2}^{4}\left(X_h^l(t)\right)\alpha, & for\ h = 1 \\ X_h^l(t)e(t)\Omega_l\Upsilon(1-f) - X_h^l(t)\Lambda(t) - X_h^l(t)\alpha, & for\ h = 2 \end{cases} \quad \textit{(Equation 17)}$$

We write super index $l$ to indicate model stages in the TB transition sequence described in the equations 1 to 23 (e.g. *i* = 1 for TB uninfected stage).

#### Country specific data for calibration

| Data stream | Brazil | Georgia | Kenya | South Africa |
|---|---|---|---|---|
| Total TB incidence per 100,000 (2022)* | 49 (42 - 56) | 60 (42 - 73) | 237 (149 - 363) | 468 (304 - 665) |
| RR-TB incidence per 100,000 (2022) * | 1.4 (0.2 - 2.64) | 12 (9.8 - 14.5) | 2.9 (1.5 - 4.3) | 39 (24 - 53) |
| TB/HIV incidence per 100,000 (2022) * | 8.9 (7.2 - 11) | 1.7 (1.1 - 2.4) | 54 (32 - 81) | 255 (166 - 362) |
| TB mortality among HIV (-) per 100,000 (2022) * | 3.4 (3.2 - 3.5) | 1.8 (1.4 - 2.3) | 32 (17 - 52) | 39 (37 - 41) |
| TB mortality among PLHIV per 100,000 (2022) * | 1.8 (1.2 - 2.6) | 0.53 (0.34 - 0.77) | 16 (10 - 25) | 52 (17 - 107) |
| TB treatment coverage % (2022) ** | 83 (73 - 97) | 66 (54 – 83) | 69 (45 - 100) | 76 (53 - 100) |
| TB prevalence per 100,000, latest year available*** | No prevalence survey | No prevalence survey | 560 (460 - 660) | 852 (679 - 1026) |
| TB prevalence/incidence among high-risk subpopulations, latest year available**** | Incidence:<br>1123 (898 – 1347) | Prevalence:<br>900 (720 - 1086) | Prevalence:<br>744 per 100,000 | Prevalence:<br>1900 per 100,000 |
| Percent of PLHIV on ART (%) – 2019***** | 71 | 69 | 80 | 71 |
| Percent PLHIV starting ART receiving TPT (%), 2019***** | 30 | 4 | 32 | 63 |
| Number of household contacts receiving TPT, 2019***** | 16,238 | 288 | 7253 | 15,392 |
| Under-five household contacts receiving TPT, 2019***** | 1614 | 30 | 5619 | 15,392 |

Abbreviations: TB, tuberculosis; RR-TB, rifampicin-resistant tuberculosis; PLHIV, people living with HIV; ART, antiretroviral therapy; TPT, tuberculosis preventive treatment
*Data points for 2022 presented here for simplicity, but calibration included the time series 2012-2022 based on WHO estimates.
**Estimated as the total reported number on TB treatment divided by the estimated TB incidence in 2022.
***Prevalence estimates from Kenya are from the 2016 prevalence survey (Enos M, et al. PLoS One, 2018); estimates from South Africa are from the 2018 prevalence survey (https://www.nicd.ac.za/wp-content/uploads/2021/02/TB-Prevalence-survey-report_A4_SA_TPS-Short_Feb-2021.pdf).
*****Data for incidence/prevalence among high-risk subpopulations per 100,000 are from the years 2022 for Brazil, 2021-2022 for Georgia, 2022 for Kenya, and 2022 for South Africa.
*****Data from 2019 is the last year available at time of calibration.

#### Model Calibration

We denote by $\theta$ the vector of input parameters, for all model inputs subject to uncertainty. For a given country, and a given parameter set $\theta$, we determine model projections for calibration targets described in **Country Specific Data for Calibration.**

To compare these model projections with data *D*, we defined the *posterior density* $\pi(\theta)$ as:

$$\pi(\theta) \propto L(D|\theta).P(\theta), \qquad \text{(Equation 18)}$$

Where $L$ is the likelihood of the data *D* given $\theta$ and *P* is the joint prior distribution for $\theta$. For *P*, we took independent uniform distributions over the ranges shown in **Model Parameters Table** The likelihood $L$ was constructed as follows. For a given country, we fit a beta distribution for proportions, and a log-normal distribution to all other calibration parameters in **Country Specific Data for Calibration**. In particular, we determined the mean and variance of these distributions in order for the 2.5th, 50th and 97.5th percentiles to match respectively the lower, mid and upper ranges of estimates. For a given parameter set $\theta$, we then constructed the overall likelihood $\pi(\theta)$ as a product of these distributions over all calibration targets listed in **Country Specific Data for Calibration**. In practice we computed the logarithm of $\pi(\theta)$, thus taking the sum of the logarithms of each of the probability densities involved.

With $\pi(\theta)$ thus defined, we sampled the posterior density using a Markov Chain Monte Carlo approach. In brief, this approach implements a random walk through the space of parameter values $\theta$ to obtain an unbiased sample of the posterior density. We implemented the 'adaptive' MCMC algorithm first introduced by *Haario et a*l which incorporates a dynamic covariance matrix to adjust endogenously the scale of 'jumps' in proposals for each of the parameter values. For the set of parameter values thus obtained, we took every tenth element to reduce autocorrelation, thus yielding an 'ensemble' of parameters $\theta_1, \theta_2, \ldots$ ; This ensemble captures simultaneously the uncertainty in the parameter inputs, as well as in the calibration data. Then, to estimate uncertainty in a given simulated output $\Gamma$ (e.g. in the reduction of incidence with a given coverage of intervention), we simulated this output $\Gamma_i$ for every $\theta_i$. We finally estimated uncertainty in $\Gamma_i$ by determining its 2.5th, 50th and 97.5th percentiles.

Below, selected model targets and model projections for each country.

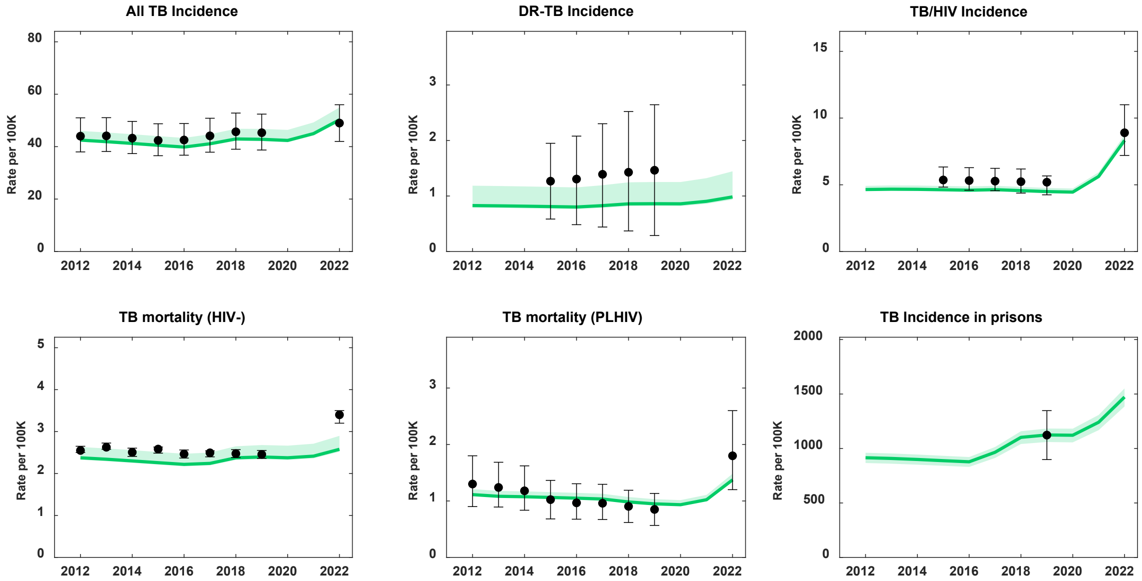

***Fig S2**: Model fit to data for Brazil. Model projections in green show the 50th percentile (solid line) of the posterior sample, and shaded area reflecting 95% Credible interval. Data points (black) from WHO country profiles or country specific sources, as detailed in **Country Specific Data for Calibration***

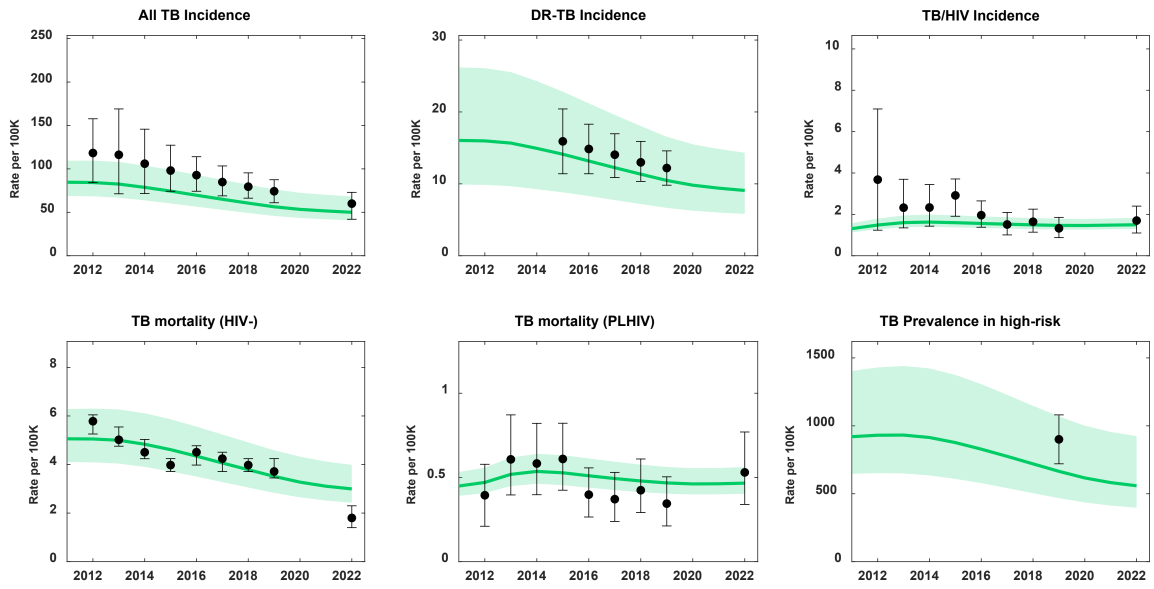

***Fig S3:*** *Model fit to data for Georgia. Model projections in green show the 50th percentile (solid line) of the posterior sample, and shaded area reflecting 95% Credible interval. Data points (black) from WHO country profiles or country specific sources, as detailed in* ***Country Specific Data for Calibration***

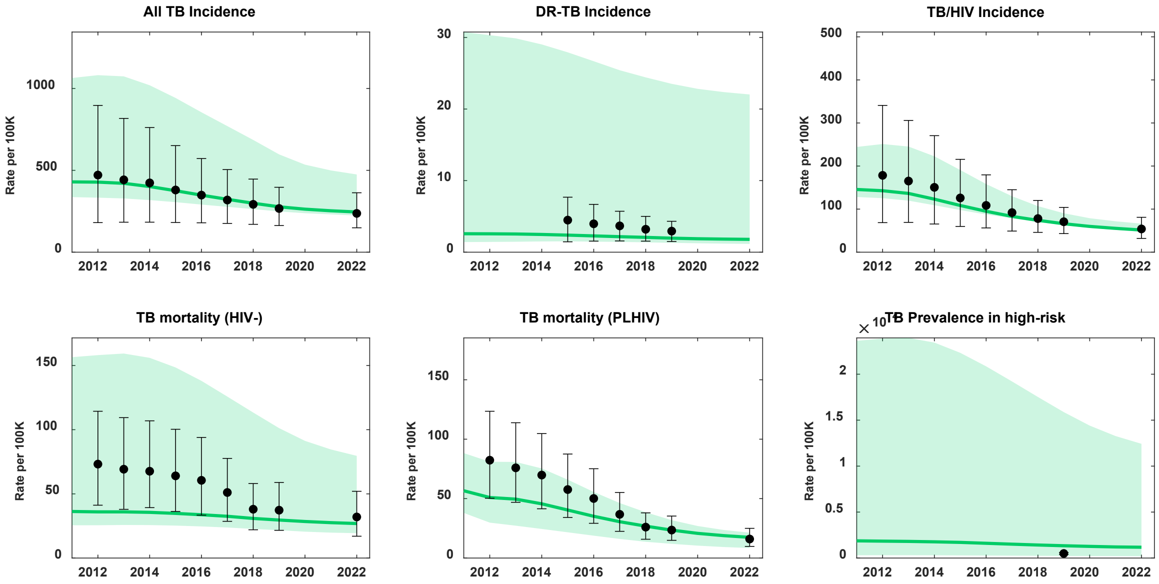

***Fig S4****: Model fit to data for Kenya. Model projections in green show the 50th percentile (solid line) of the posterior sample, and shaded area reflecting 95% Credible interval. Data points (black) from WHO country profiles or country specific sources, as detailed in* ***Country Specific Data for Calibration***

.

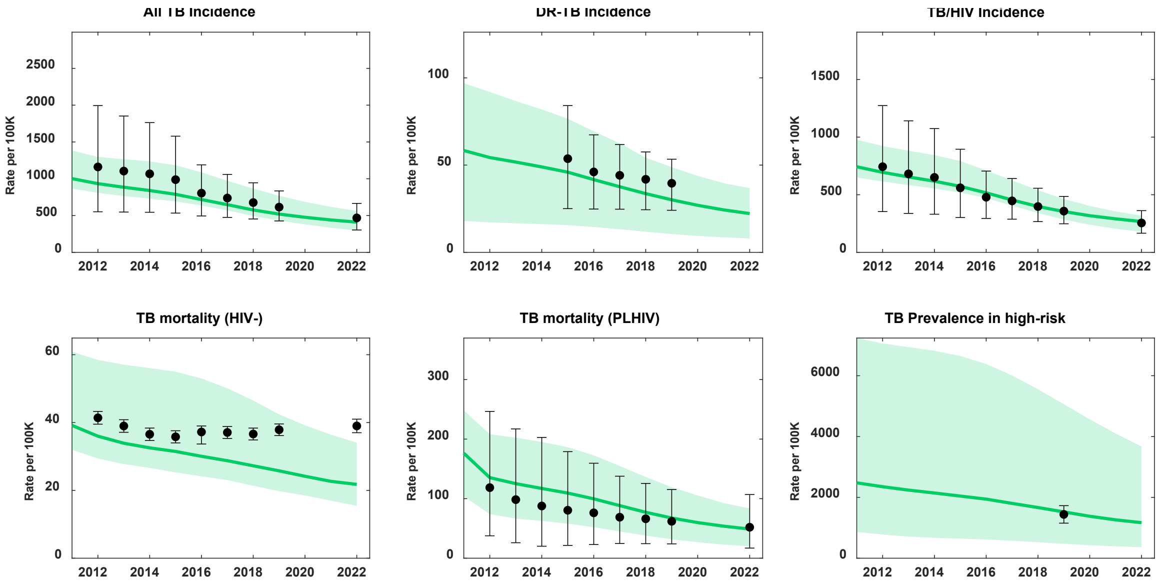

***Fig S5:*** *Model fit to data for South Africa. Model projections in green show the 50th percentile (solid line) of the posterior sample, and shaded area reflecting 95% Credible interval. Data points (black) from WHO country profiles or country specific sources, as detailed in* ***Country Specific Data for Calibration***

References this section: Haario H, Saksman E, Tamminen J. An adaptive Metropolis algorithm.Bernoulli. 2001;7:223–42

#### MCMC Diagnostics

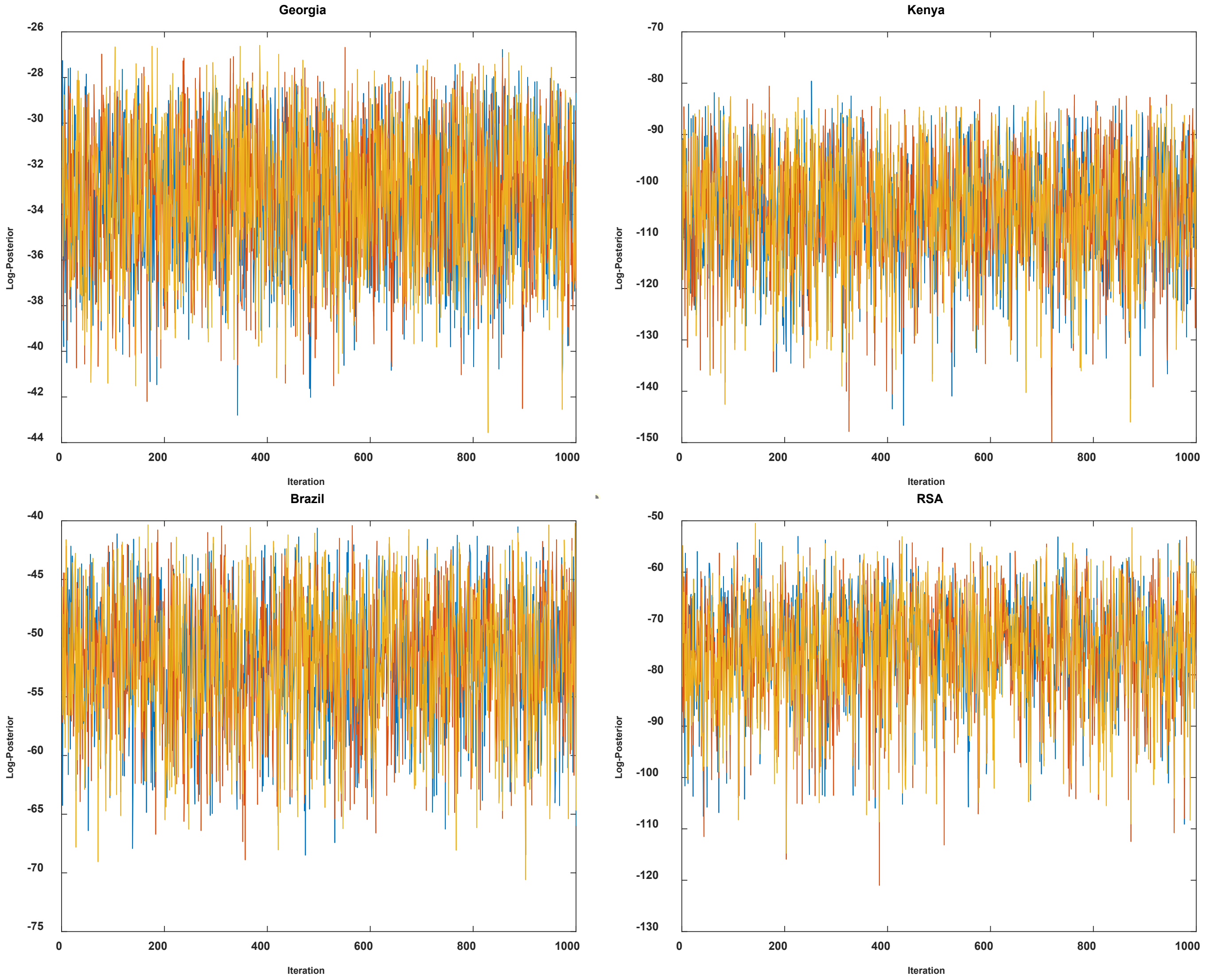

**Fig S6** Trace plots of Log-posterior for the four calibrated countries. Each panel showing convergence of three MCMC chains. Each chain originally of >150,000 iterations, and in display we show 1000 samples from the posterior distribution after burn-in period removed and chains thinned at a ratio 1:10. Convergence was examined visually by Gelman-Rubin test (Gelman et al).

References this section: Gelman A, Rubin DB. Inference from iterative simulation using multiple sequences. Stat Sci. 1992;7:457–72.

#### Additional Model Assumptions

While we allow the set of free parameters to calibrate against the data targets, we introduced country specific changes to account for the changes in tuberculosis epidemic trends caused by COVID-19 disruptions. We examined incidence and mortality trends individually, and we found it was necessary to introduce disruptions for the case studies of Brazil and South Africa to reproduce the observed trends between 2019 and 2022. Hence, we allow an extra free parameter for these two countries. This parameter acts specifically by reducing testing capacity and treatment coverage during the pandemic period. We assume that any disruptions during COVID-19 are resolved by 2023.

To account for mortality that is attributable to tuberculosis even after treatment, we assume that only deaths arising from the high relapse (i.e., $R^{(hi)}$) compartment are accounted for. This is captured by assuming an increased risk of death in the high relapse compartment of ~10% relative to general population. The average time spent in this compartment is ~6 months.

Our approach to modelling the effect of TPT assumes that observed trial efficacy arises from a single mechanism by which the progression from tuberculosis infection to tuberculosis disease is reduced by a factor equal to the remainder of the relative risk (i.e, 1-RR). The implication of this is that we do not consider infection clearance as a background mechanism of efficacy. We consider this a conservative approach given the lack of information existing for accurately attributing specific values of effect to clearance and progression reduction mechanisms respectively.

#### Model Parameters Table

| Parameter | Symbol | Value | | | | Source/Notes |
|---|---|---|---|---|---|---|
| | | South Africa | Kenya | Georgia | Brazil | |
| **TB natural history** | | | | | | |
| Mean rate of transmission per person with DS-TB | $\beta_{ds}$ | 13.3 (95% CrI 9.3 – 19.8) | 18 (95% CrI 10.5 – 22) | 4 (95% CrI 2.5 – 6.4) | 7.1 (95% CrI 6.5 – 12) | Model estimate |
| Mean rate of transmission per person with RR-TB | $\beta_{dr}$ | 9.1 (95% CrI 7.5 – 13.7) | 12 (95% CrI 8 – 14) | 3.8 (95% CrI 2.2 – 6.1) | 5.27 (95% CrI 4 – 6.9) | Model estimate |
| Breakdown to active disease in slow progressors | $\gamma^{slow}$ | 0.000594 | | | | Ref 1 |
| Breakdown to active disease in fast progressors | $\gamma^{fast}$ | 0.0826 | | | | Ref 1 |
| Rate of transition to the slow latent compartment | $\delta$ | 0.87 | | | | Ref 1 |
| Increased progression to TB in $\boldsymbol{HIV^{+}}$ relative to $\boldsymbol{HIV^{-}}$ | $\varepsilon$ | 29 (95% CrI 27.5 – 32) | 35 (95% CrI 22-38) | 24.5 (95%CrI 22-28) | 19.5 (95%CrI 14 - 24) | Model estimate |
| Relapse, per-capita hazard rates | r(hi) | relapse following treatment default | 0.14 | | | Ref 2, 3, 4 |
| | r(lo) | relapse >2 years after treatment | 0.0015 | | | |
| 'Stabilisation' of relapse risk following treatment | $\sigma$ | 0.5 | | | | Based on Thomas et al (Ref 3): most relapse occurs in first 2 yr after treatment. |
| TB mortality rate | $\mu^{(tb)}$ | 0.19 (95% CrI 0.12 – 0.24) | 0.18 (95% CrI 0.16 – 0.21) | 0.07 (95% CrI 0.02 – 0.19) | 0.18 (95% CrI 0.12 – 0.22) | Specified together to yield ~50% cure, ~50% mortality in average of 3 years. Tiemersma et al (Ref 5) |
| Spontaneous cure | $\theta$ | 0.15 (95% CrI 0.14 –0.18) | | | | |
| Relative Risk of TB mortality in HIV+ | $RR^{(h+)}$ | 3 (95% CrI 1.45-6.8) | 4.5 (95% CrI 3.2-6.8) | 5.8 (95% CrI 3-8.4) | 8.1 (95% CrI 7-12) | Model estimate |
| Relative risk of TB-associated mortality in the 5 years post-TB treatment | | 1.14 (1.02 to 1.34) | | | | Ref 6 |
| Reduced susceptibility from past infection | $\iota$ | [0.25 –0.75] | | | | Assumed range (uniform distribution) |

| Parameter | Symbol | Value | | | | Source/Notes |
|---|---|---|---|---|---|---|
| | | South Africa | Kenya | Georgia | Brazil | |
| **Health system** | | | | | | |
| Per-capita rate of initial presentation to care | κ | 1.8 (95% CrI 1 – 4.9) | 2.3 (95% CrI 1.7 – 4.2) | 3 (95% CrI 1.5 – 5.2) | 3.5 (95% CrI 1.9 – 6) | Model estimate: corresponds to mean initial patient delay of 4.6 months (95% CrI 3.6 – 6) |
| Treatment initiation delay | ξ | 52 | | | | Sreeramareddy et al (Ref 7); corresponds a mean treatment delay of 1 week |
| Treatment initiation probability | ϱ | 0.87 (95% CrI 0.75 – 0.96) | 0.95 (95% CrI 0.88 – 0.99) | 0.92 (95% CrI 0.81 – 0.95) | 0.9 (95% CrI 0.69 – 0.97) | Model estimate |
| Rate of recruitment into ART | Π | 1.7 (95% CrI 1.3 – 2.8) | 2.5 (95% CrI 1.5 – 6.8) | 5.5 (95% CrI 3.3 – 9.5) | 4.9 (95% CrI 3.5 – 9.1) | Model estimate |
| Fraction of new ART starters enrolled in TPT | A | 0.66 (95% CrI 0.52 – 0.82) | 0.35 (95% CrI 0.2 – 0.45) | 0.05 (95% CrI 0.01 – 0.1) | 0.28 (95% CrI 0.15 – 0.3) | Model estimate |
| **TB Preventive treatment (parameters for a baseline 6 months course of isoniazid - 6H)** | | | | | | |
| TPT regimen duration (months) | Γ | 6 | | | | Regimen characteristic |
| Suppression of reactivation effect | e | 70% | | | | Whalen et al (Ref 8) |
| Serious Adverse Event Rate | | Adults = 2%; Children = 0.2% | | | | Ref 9, 10 |
| **TB Preventive treatment (parameters for a regimen of three-months daily isoniazid and rifampicin – 3HR)** | | | | | | |
| TPT regimen duration (months) | Γ | 3 | | | | Regimen characteristic |
| Suppression of reactivation effect | e | 70% | | | | Equivalent to 6H, Ref 11 |
| Serious Adverse Event Rate | | Adults = 2%; Children = 0.2% | | | | Assumed equivalent to 6H, Ref 11 |
| **TB Preventive treatment (parameters for a regimen of three-months once-weekly isoniazid and rifapentine – 3HP)** | | | | | | |
| TPT regimen duration (months) | Γ | 3 | | | | Regimen characteristic |
| Suppression of reactivation effect | e | 93% | | | | Equivalent to 9 months isoniazid (Ref 12, 13) |
| Serious Adverse Event Rate | | Adults = 2%; Children = 0.2% | | | | Ref 9 |
| **Serious Adverse Events with TB disease treatment (treatment-related, grade 3 or higher)** | | | | | | |

| Parameter | Symbol | Value | | | | Source/Notes |
|---|---|---|---|---|---|---|
| | | South Africa | Kenya | Georgia | Brazil | |
| SAE standard DS-TB (adults) | | 9.8% | | | | Ref 14 |
| SAE standard DS-TB (children) | | 2.2% | | | | Ref 15 |
| SAE short DS-TB (children) | | 1% | | | | Ref 15 |
| SAE long RR-TB | | 58% | | | | Ref 16 |
| SAE short RR-TB | | 18% | | | | Ref 16 |
| **Demographics** | | | | | | |
| Birth rate | B | 0.013 | 0.023 | 0.0001 | 0.008 | Ref 17– adjusted to yield annual population growth from 1970 |
| Background mortality rate | $\mu$ | 0.016 | 0.015 | 0.013 | 0.013 | Ref 18 |
| Number of PLHIV, 2022 | | 990,000 | 8500 | 1,400,000 | 7,600,000 | Ref 19 |
| HHC Per Index Patient | | 2.31 | 2.42 | 2.75 | 2.36 | Ref 20 |
| **TB Epidemiology** | | | | | | |
| Proportion HHC TB infection, < 5 years (%) | | 35.5 (95% UR 30.3-41.1) | | | | Ref 21 |
| Proportion HHC TB infection, 5-14 years (%) | | 53.1 (95% UR 42-63.9) | | | | Ref 21 |
| Proportion HHC TB infection, ≥ 15 years (%) | | 65.3 (95% UR 35.5-86.5) | | | | Ref 21 |
| Proportion HHC TB disease | | 3% | | | | Ref 21 |
| Proportion PLHIV TB infection (2024) | | 65% (95%CrI 55 - 70) | 25% (95%CrI 18 - 32) | 2% (95%CrI 1 - 3) | 17% (95%CrI 16 - 19) | Model estimate |
| Proportion High-Risk Community TB infection (2024) | | 9% (95%CrI 3 - 22) | 12% (95%CrI 4 - 35) | 6% (95%CrI 5 - 8) | 6% (95%CrI 5 - 8) | Model estimate |
| **DALY Information** | | | | | | |
| TB Disease DALY (event-based) | | 0.333 (95% UR 0.274-0.549) | | | | Ref 22 |

| Parameter | Symbol | Value | | | | Source/Notes |
|---|---|---|---|---|---|---|
| | | South Africa | Kenya | Georgia | Brazil | |
| TB Disease with HIV DALY (event-based) | | 0.408 (95% UR 0.274-0.549) | | | | Ref 22 |
| Post-TB Lung Disease (annual) | | 0.036 (95% UR 0.006-0.088) | | | | Ref 6 |
| TB-mortality DALY | | Based on life expectancy at time of death for each country | | | | Ref 18 |
| SAE DALY (event-based) | | 0.02 (0.01 to 0.03) | | | | Ref 22 |

Abbreviations: 95% UR, 95% uncertainty range; 95% CrI, 95% credible interval; TB, tuberculosis; HHC, household contacts; PLHIV, people living with HIV; DALY, disability adjusted life year; SAE, serious adverse event; HIV, human immunodeficiency virus; ART, antiretroviral treatment; TPT, tuberculosis preventive treatment; DS-TB, drug-susceptible tuberculosis; RR-TB, rifampicin-resistant tuberculosis.

**References this section:**

1. Menzies NA, Wolf E, Connors D, Bellerose M, Sbarra AN, Cohen T, et al. Progression from latent infection to active disease in dynamic tuberculosis transmission models: a systematic review of the validity of modelling assumptions. Lancet Infect Dis. 2018;18:e228–38.
2. Driver CR, Munsiff SS, Li J, Kundamal N, Osahan SS. Relapse in persons treated for drug-susceptible tuberculosis in a population with high coinfection with human immunodeficiency virus in New York City. Clin Infect Dis. 2001;33:1762–9.
3. Thomas A, Gopi PG, Santha T, Chandrasekaran V, Subramani R, Selvakumar N, et al. Predictors of relapse among pulmonary tuberculosis patients treated in a DOTS programme in South India. Int J Tuberc Lung Dis.2005;9:556–61.
4. Menzies D, Benedetti A, Paydar A, Martin I, Royce S, Pai M, et al. Effect of duration and intermittency of rifampin on tuberculosis treatment outcomes: a systematic review and meta-analysis. PLoS Med. 2009;6:e1000146.
5. Tiemersma EW, van der Werf MJ, Borgdorff MW, Williams BG, Nagelkerke NJ. Natural history of tuberculosis: duration and fatality of untreated pulmonary tuberculosis in HIV negative patients: a systematic review. PLoSOne. 2011;6:e17601.
6. Menzies NA, Quaife M, Allwood BW, et al. Lifetime burden of disease due to incident tuberculosis: a global reappraisal including post-tuberculosis sequelae. Lancet Glob Health 2021; 9: e1679–87.
7. Sreeramareddy CT, Qin ZZ, Satyanarayana S, Subbaraman R, Pai M. Delays in diagnosis and treatment of pulmonary tuberculosis in India: a systematic review. Int J Tuberc Lung Dis. 2014;18:255–66.
8. Whalen CC, Johnson JL, Okwera A, Hom DL, Huebner R, Mugyenyi P, et al. A trial of three regimens to prevent tuberculosis in Ugandan adults infected with the human immunodeficiency virus. N Engl J Med.1997;337:801–8.
9. Winters N, Belknap R, Benedetti A, et al. Completion, safety, and efficacy of tuberculosis preventive treatment regimens containing rifampicin or rifapentine: an individual patient data network meta-analysis. Lancet Respir Med 2023; 11: 782–90.
10. Diallo T, Adjobimey M, Ruslami R, et al. Safety and Side Effects of Rifampin versus Isoniazid in Children. N Engl J Med 2018; 379: 454–63.
11. Zenner D, Beer N, Harris RJ, Lipman MC, Stagg HR, van der Werf MJ. Treatment of Latent Tuberculosis Infection: An Updated Network Meta-analysis. Ann Intern Med 2017; 167: 248–55.
12. International Union Against Tuberculosis Committee on Prophylaxis. Efficacy of various durations of isoniazid preventive therapy for tuberculosis: five years of follow-up in the IUAT trial. Bull World Health Organ 1982; 60: 555–64.
13. Sterling TR, Villarino ME, Borisov AS, et al. Three months of rifapentine and isoniazid for latent tuberculosis infection. N Engl J Med 2011; 365: 2155–66.
14. Dorman SE, Nahid P, Kurbatova E V., Phillips PPJ, Bryant K, Dooley KE, et al. Four-Month Rifapentine Regimens with or without Moxifloxacin for Tuberculosis. New England Journal of Medicine. 2021;384:1705–18.
15. Turkova A, Wills GH, Wobudeya E, Chabala C, Palmer M, Kinikar A, et al. Shorter Treatment for Nonsevere Tuberculosis in African and Indian Children. New England Journal of Medicine. 2022;386:911–22.
16. Nyang'wa BT, Berry C, Kazounis E, Motta I, Parpieva N, Tigay Z, et al. A 24-Week, All-Oral Regimen for Rifampin-Resistant Tuberculosis. New England Journal of Medicine. 2022;387:2331–43.
17. World Bank. Total Fertility Rate. 2023. https://data.worldbank.org/indicator/SP.DYN.TFRT.IN (accessed March 11, 2024)
18. Global Health Observatory. Life tables by country (GHE: Life tables). 2023. https://www.who.int/data/gho/data/indicators/indicator-details/GHO/gho-ghe-life-tables-by-country (accessed March 11, 2024).
19. UNAIDS. Country Data Tables. 2023. https://www.unaids.org/en/regionscountries/countries (accessed March 11, 2024)
20. United Nations Department of Economic and Social Affairs. Household: Size and Composition, 2022. 2022. https://population.un.org/Household/#/countries/840 (accessed March 11, 2024).
21. Fox GJ, Barry SE, Britton WJ, Marks GB. Contact investigation for tuberculosis: a systematic review and meta-analysis. Eur Respir J 2013; 41: 140–56.
22. Global Burden of Disease Collaborative Network. Global Burden of Disease Study 2019 (GBD 2019) Disability Weights. 2020.

#### Cost Estimates and 95% UR for TB-related activities by country (All Costs 2023 USD)

| Cost Parameter | Brazil Costs (95% UR) | Georgia Costs (95% UR) | Kenya Costs (95% UR) | South Africa Costs (95% UR) |
|---|---|---|---|---|
| Cost to perform symptom screening per person | 2.94 (0.58 to 7.17) | 3.76 (1.10 to 8.00) | 2.98 (0.37 to 8.22) | 0.74 (0.14 to 1.81) |
| Cost of household contact investigation per index patient | 2.29 (0.36 to 5.98) | 15.69 (13.07 to 18.57) | 17.69 (0.11 to 76.91) | 8.53 (1.35 to 22.25) |
| Cost of TST, inclusive of materials and personnel time | 5.59 (4.56 to 6.73) | 5.08 (4.13 to 6.13) | 10.28 (8.37 to 12.39) | 4.05 (3.29 to 4.89) |
| Cost of IGRA, inclusive of materials | 24.65 (20.08 to 29.69) | 21.02 (17.11 to 25.33) | 30.06 (24.48 to 36.21) | 78.5 (63.95 to 94.54) |
| Cost of antigen-based skin test, inclusive of materials and personnel time | 6.11 (4.98 to 7.35) | 5.14 (4.17 to 6.21) | 10.34 (8.4 to 12.47) | 4.72 (3.84 to 5.68) |
| Cost of Xpert per person | 20.88 (4.74 to 48.88) | 18.74 (1.77 to 55.42) | 19.91 (8.1 to 37.02) | 17.93 (4.49 to 40.6) |
| Cost of HIV test per person | 6.93 (5.64 to 8.35) | 4.02 (3.28 to 4.84) | 7.64 (6.23 to 9.19) | 0.52 (0.41 to 0.64) |
| Cost of CRP per person | 6.4 (0.19 to 23.11) | 4.02 (0.12 to 14.52) | 7.64 (0.22 to 27.6) | 6.54 (0.19 to 23.63) |
| Cost of CXR per person | 6.58 (0.65 to 19.13) | 3.01 (1.03 to 6.01) | 30.72 (0.47 to 119.99) | 18.72 (1.85 to 54.4) |
| Annual cost of CAD license (annuitized); considered in analysis as a new investment and part of implementation in 5% of all healthcare facilities | 1057.98 (605.64 to 1633.73) | 1057.98 (605.64 to 1633.73) | 1057.98 (605.64 to 1633.73) | 1057.98 (605.64 to 1633.73) |
| Cost of AFB smear, ZN | 2.91 (0.26 to 8.64) | 5.88 (1.66 to 12.7) | 17.41 (0.32 to 66.83) | 2.35 (0.22 to 6.94) |
| Cost of sputum collection per person | 1.78 (0.01 to 7.84) | 2.49 (0.22 to 7.43) | 8.17 (0 to 47.73) | 3.64 (0.02 to 16.05) |
| Cost of liquid culture per sample | 52.67 (34.96 to 73.84) | 22.05 (14.63 to 30.9) | 35.19 (23.35 to 49.33) | 7.35 (4.87 to 10.31) |
| Cost of solid culture per sample | 32.51 (21.6 to 45.57) | 12.14 (8.08 to 17) | 46.43 (30.84 to 65.1) | 9.69 (6.42 to 13.6) |
| Cost of first-line DST per panel | 183.64 (21.01 to 518.13) | 27.92 (3.2 to 78.76) | 52.88 (0.26 to 233.5) | 30.9 (3.53 to 87.19) |
| Cost of second-line DST per panel | 173.11 (19.93 to 485.38) | 17.73 (2.05 to 49.7) | 33.55 (0.17 to 146.87) | 23.16 (2.67 to 64.95) |
| Cost of first-line LPA per sample | 38.74 (4.44 to 109.06) | 88.91 (10.19 to 250.29) | 61.99 (39.17 to 89.88) | 14.59 (1.67 to 41.05) |
| Cost of second-line LPA per sample | 45.25 (5.23 to 127.49) | 88.85 (10.28 to 250.38) | 61.98 (39.23 to 89.89) | 14.58 (1.69 to 41.07) |
| Cost of pre-TPT evaluation, considering a treatment visit | 4.36 (0.62 to 11.58) | 2.98 (0.49 to 7.65) | 5.35 (0.67 to 14.68) | 2.58 (0.37 to 6.87) |
| Cost of 6INH if complete, adults | 16.58 (8.33 to 27.59) | 30.45 (15.3 to 50.67) | 35.22 (17.69 to 58.64) | 13.31 (6.69 to 22.15) |
| Cost of 6INH if complete, children | 19.16 (9.64 to 31.89) | 54.87 (27.62 to 91.33) | 73.85 (37.16 to 122.94) | 22.81 (11.48 to 37.96) |
| Cost of 3HR if complete, adults | 22.61 (11.4 to 37.67) | 29.66 (14.97 to 49.4) | 34.82 (17.57 to 58) | 20.17 (10.16 to 33.63) |
| Cost of 3HR if complete, children | 14.51 (7.31 to 24.11) | 33.73 (17 to 56.05) | 27.69 (13.95 to 46.02) | 45.7 (23.02 to 76) |
| Cost of 3HP if complete, adults | 36.74 (10.09 to 80.52) | 50.08 (13.75 to 109.75) | 28.65 (7.86 to 62.8) | 21.39 (5.87 to 46.89) |
| Cost of 3HP if complete, children | 35.04 (9.6 to 76.6) | 50.08 (13.73 to 109.45) | 28.65 (7.85 to 62.63) | 21.39 (5.86 to 46.76) |
| Cost of 1HP if complete, adults | 29.86 (3.02 to 86.91) | 17.91 (1.81 to 52.12) | 28.43 (2.87 to 82.74) | 23.72 (2.4 to 69.04) |
| Cost of 1HP if complete, children | 22.8 (2.3 to 66) | 26.07 (2.63 to 75.44) | 35.15 (3.54 to 101.71) | 23.72 (2.39 to 68.64) |
| Cost of 6Lfx if complete, adults | 48.78 (24.57 to 81.35) | 38.6 (19.45 to 64.36) | 42.54 (21.42 to 70.95) | 26.3 (13.24 to 43.87) |
| Cost of 6Lfx if complete, children | 28.59 (14.38 to 47.6) | 36.56 (18.38 to 60.88) | 36.52 (18.38 to 60.79) | 24.33 (12.24 to 40.51) |
| Cost of 3HR if incomplete, adults; assumes 50% the price of complete | 11.3 (4.67 to 20.85) | 15.01 (7.56 to 25.03) | 17.41 (5.83 to 35.31) | 10.09 (4.17 to 18.63) |
| Cost of 3HR if incomplete, children; assumes 50% the price of complete | 7.25 (3.65 to 12.05) | 17.06 (8.59 to 28.33) | 13.86 (6.98 to 23.01) | 22.87 (11.5 to 38.01) |
| Cost of 6INH if incomplete, adults; assumes 50% the price of complete | 8.3 (4.17 to 13.8) | 15.42 (7.73 to 25.68) | 17.62 (8.84 to 29.35) | 6.66 (3.34 to 11.1) |
| Cost of 6INH if incomplete, children; assumes 50% the price of complete | 9.57 (4.81 to 15.95) | 27.62 (13.91 to 45.99) | 36.93 (18.57 to 61.55) | 11.41 (5.74 to 19.01) |
| Cost of 3HP if incomplete, adults; assumes 50% the price of complete | 18.37 (5.02 to 40.11) | 25.22 (6.89 to 55.07) | 14.33 (3.92 to 31.28) | 10.7 (2.92 to 23.36) |
| Cost of 3HP if incomplete, children; assumes 50% the price of complete | 17.52 (4.82 to 38.18) | 25.22 (6.94 to 54.96) | 14.33 (3.95 to 31.21) | 10.7 (2.94 to 23.32) |

| Cost Parameter | Brazil Costs (95% UR) | Georgia Costs (95% UR) | Kenya Costs (95% UR) | South Africa Costs (95% UR) |
|---|---|---|---|---|
| Cost of 1HP if incomplete, adults; assumes 50% the price of complete | 14.93 (1.51 to 43.38) | 9.14 (0.93 to 26.53) | 14.22 (1.44 to 41.32) | 11.86 (1.2 to 34.47) |
| Cost of 1HP if incomplete, children; assumes 50% the price of complete | 11.4 (1.15 to 33.18) | 13.22 (1.33 to 38.47) | 17.58 (1.77 to 51.19) | 11.86 (1.19 to 34.54) |
| Cost of 6Lfx if incomplete, adults; assumes 50% the price of complete | 24.4 (12.3 to 40.63) | 19.48 (9.81 to 32.45) | 21.27 (10.72 to 35.42) | 13.15 (6.63 to 21.89) |
| Cost of 6Lfx if incomplete, children; assumes 50% the price of complete | 14.3 (7.2 to 23.79) | 18.46 (9.28 to 30.75) | 18.26 (9.19 to 30.39) | 12.17 (6.13 to 20.25) |
| Cost of drug-susceptible TB treatment, adults | 567.07 (155.63 to 1242.76) | 325.4 (89.3 to 713.13) | 191.24 (52.48 to 419.12) | 133.57 (36.65 to 292.74) |
| Cost of 4-month DS-TB treatment, children | 487.43 (134.1 to 1063.54) | 385.16 (105.96 to 840.42) | 134.11 (36.89 to 292.64) | 67.16 (18.48 to 146.54) |
| Cost of 6-month DS-TB treatment, adults | 541.38 (148.2 to 1184.22) | 611.87 (167.5 to 1338.43) | 186.61 (51.08 to 408.2) | 98.47 (26.95 to 215.41) |
| Cost of BPaLM (or BPaLL, in South Africa) RR-TB treatment, adults | 3104.77 (850.62 to 6796.16) | 2379.05 (651.8 to 5207.61) | 3385.46 (927.52 to 7410.61) | 1030.71 (282.38 to 2256.18) |
| Cost of BPaLM (or BPaLL, in South Africa) RR-TB treatment, children | 4393.91 (1206.38 to 9618.67) | 2336.98 (641.63 to 5115.86) | 3301.22 (906.37 to 7226.69) | 1009.01 (277.03 to 2208.81) |
| Cost of 18-month all oral RR-TB treatment, adults | 7497.07 (2060.08 to 16386.49) | 3367.87 (925.43 to 7361.23) | 7338.97 (2016.63 to 16040.95) | 2540.43 (698.07 to 5552.69) |
| Cost of 18-month all oral RR-TB treatment, children | 8798.78 (2397.94 to 19258.05) | 3678.68 (1002.55 to 8051.6) | 7938.57 (2163.51 to 17375.3) | 3296.63 (898.43 to 7215.39) |
| Patient costs per month of DS-TB treatment; mean episode costs were divided by average duration of treatment (6 months) | 281.36 (35.22 to 771.31) | 414.7 (51.9 to 1136.87) | 57.18 (7.15 to 156.76) | 110.4 (13.82 to 302.64) |
| Patient costs per month of RR-TB treatment; mean episode costs were divided by average duration of treatment (18 months) | 281.41 (13.22 to 942.22) | 414.77 (19.48 to 1388.75) | 106.93 (5.92 to 348.19) | 110.42 (5.19 to 369.69) |
| Cost of post-tuberculosis healthcare utilization; considers 12.2 outpatient visits and 2.2 additional inpatient bed days | 192.66 (4.89 to 710.23) | 283.92 (7.21 to 1046.67) | 102.53 (2.6 to 377.99) | 546.78 (13.88 to 2015.69) |
| Patient costs per month of TPT; assumed as a proportion of patient costs of DS-TB treatment (9.1%) | 25.25 (3.22 to 69.56) | 37.67 (4.79 to 103.78) | 5.19 (0.66 to 14.28) | 10.03 (1.28 to 27.59) |
| Annual cost of healthcare worker training in intervention scenarios; annual recurrent 1-day training cost in intervention scenarios for training of 10% of healthcare workforce, plus TST training | 24,600,712.52 (20,042,482.12 to 29,610,967.59) | 330,009.56 (268,862.57 to 397,220.3) | 734,021.26 (598,015.52 to 883,514.24) | 5,800,167.99 (4,725,463.26 to 6,981,447.64) |
| Annual cost of active case finding training in years conducted for The Intervention Package; assumes 2 days training for all years of case finding in high-risk communities, applied as an annual implementation cost | 76,998.12 (62,704.76 to 92,670.81) | 14,999.63 (12,215.21 to 18,052.76) | 7999.8 (6514.78 to 9628.14) | 86,997.87 (70,848.23 to 104,705.98) |
| Annual cost of active case finding training in years conducted, enhanced scenario; assumes 2 days training for all years of case finding in high-risk communities, applied as an annual implementation cost | 107,993.75 (87,938.35 to 130,040.84) | 17,998.96 (14,656.39 to 21,673.47) | 9999.42 (8142.44 to 12,040.82) | 103,993.98 (84,681.38 to 125,224.51) |
| Cost to screen one person during active case finding for The Intervention Package; considers costs of materials (cooler, ice pack, fuel, van), 50% of nights stayed out of town (hotel, food), personnel costs | 10.25 (5.86 to 15.82) | 13.14 (7.51 to 20.29) | 6.12 (3.50 to 9.44) | 19.4 (11.10 to 29.94) |
| Cost to screen one person during active case finding, enhanced scenario; considers costs of materials (cooler, ice pack, fuel, van), 50% of nights stayed out of town (hotel, food), personnel costs, and cost of antigen-based skin test | 14.26 (8.18 to 22.02) | 16.67 (9.56 to 25.74) | 8.12 (4.66 to 12.54) | 23.04 (13.22 to 35.57) |

Abbreviations: 95% UR, 95% uncertainty range; TST, tuberculin skin test; IGRA, interferon-gamma release assay; HIV, human immunodeficiency virus; CRP, C-reactive protein; CXR, chest x-ray; CAD, computer aided detection; AFB, acid-fast bacilli; ZN, Ziehl-Nielsen; DST, drug susceptibility test; LPA, line probe assay; TPT, tuberculosis preventive treatment; 6INH, six-months of isoniazid daily; 3HR, three-months isoniazid and rifampicin daily; 3HP, three-months isoniazid and rifapentine weekly; 1HP, one-month isoniazid and rifapentine daily; 6Lfx, six months levofloxacin daily; DS-TB, drug-susceptible tuberculosis; RR-TB, rifampicin-resistant tuberculosis; BPaLM, bedaquiline, pretomanid, linezolid, and moxifloxacin; BPaLL, bedaquiline, pretomanid, linezolid, and levofloxacin.

#### Detailed Costing Estimation Methods for Each Country

##### Brazil

Costs were estimated in Brazil through micro-costing methods largely using federal reimbursement data, adjusted for "under-reimbursement" as estimated by Pinto and colleagues. All costs were reviewed by the national tuberculosis programme, who provided some additional costs (eg, first-line drug susceptibility testing). The composition of the 18-month rifampicin-resistant tuberculosis regimen was assumed to be bedaquiline (six months), terizidone, linezolid, and levofloxacin. We assumed 13% of all individuals receiving tuberculosis preventive treatment received rifamycin-based regimens and 5% of all individuals treated for rifampicin-resistant tuberculosis received short regimens in 2024. These values increased to 50% by 2030 in status quo scenarios, but increased to 100% by 2030 with The Intervention Package.

We worked with the national tuberculosis programme and others with experience screening high-risk communities—in this case, people deprived of liberty. Based on a population size of 650,000 and a target to screen on 60% of this population, we estimated 3 teams composed of one nurse, one physician, and one corrections officer each would be deployed per state (27 states) to conduct this screening at a rate of 80 people screened per day. The screening would take 60 days to complete. For each team, we assumed they would have a mobile van fully equipped with chest x-ray, with an annual, annuitized cost of $55,100 USD, they would require $1000 USD in fuel costs, $290 in coolers and ice to transport sputum samples, 30 days in hotel (at a cost of $50 per night), and a per diem of $13 per person per day. Complete costs are described below.

| Cost Parameter | Description | Mean (USD 2023) |
|---|---|---|
| Cost to perform symptom screening per person | Cost of a diagnostic patient visit according to federal reimbursement (2023) | 2.94 |
| Cost of household contact investigation per index patient | Cost of a nurse or physician (average) performing over-the-phone contact investigation (7.5 mins). Time estimate from Alsdurf et al, time cost estimated through real salaries collected in Manaus, Porto Alegre, Sao Paolo, Recife, and Rio in 2023 | 2.29 |
| Cost of TST, inclusive of materials and personnel time | Cost of a single patient visit (assuming TST is done in conjunction with another visit) and all materials required. Costs from Rio de Janeiro 2023 | 5.59 |
| Cost of IGRA, inclusive of materials | Cost as reported by the National TB Program in Brazil based on purchase records (91.23 BRL) and estimation of other costs from human resources, consumables, and equipment (5.12 USD, inflated at 30% from 2020-2023 to $6.66 found in: https://www.nature.com/articles/s41598-020-78737-w; converted to USD using direct exchange rates (0.197:1) | 24.65 |
| Cost of antigen-based skin test, inclusive of materials and personnel time | Same non-antigen costs of the TST, except using the dia-skin test reported cost ($1.60 USD) in place of tuberculin. Dia-skin test cost has been reported to be consistent over time and no inflation factor applied. | 6.11 |
| Cost of Xpert per person | Cost of cartridge as reported by National TB Program in Brazil (45.18 BRL, converted 0.197:1), subtracting $2 price reduction in late 2023 ($6.91). Also includes non-cartridge costs as estimated in three representative labs elsewhere (https://www.ncbi.nlm.nih.gov/pmc/articles/PMC4722795/); converted to USD using study reported rate and then inflated using country specific GDP deflator used (2012) - 111% inflation | 20.88 |
| Cost of HIV test per person | Federal reimbursement for HIV testing (2023), with factor adjustment as estimated by Pinto and colleagues (3.51). Converted to USD at a rate of 0.197:1 | 6.93 |
| Cost of CRP per person | Federal reimbursement for CRP (2023), with factor adjustment as estimated by Pinto and colleagues (3.51). Converted to USD at a rate of 0.197:1 | 6.4 |
| Cost of CXR per person | Federal reimbursement for CXR (2023), with factor adjustment as estimated by Pinto and colleagues (3.51). Converted to USD at a rate of 0.197:1 | 6.58 |
| Annual cost of CAD license (annuitized); considered in analysis as a new investment and part of implementation in 5% of all healthcare facilities | InferRead CAD software (one license), with annual costs and annuitized capital costs at 3% over 10 years for software and installation/training, 5 years for laptop, and 3 years for initial maintenance agreement with the next years costing $250 per annum (GDF price catalogue) | 1058 |
| Cost of AFB smear, ZN | Federal reimbursement for ZN smear (2023), with factor adjustment as estimated by Pinto and colleagues (3.51). Converted to USD at a rate of 0.197:1 | 2.91 |
| Cost of sputum collection per person | Use nurse salaries (from the five-city study described in the household contact investigation row) and assumed 20 mins for sputum collection | 1.78 |
| Cost of liquid culture per sample | Cost provided by the National Tuberculosis Program (2023) and converted to USD at a rate of 0.197:1 | 52.68 |
| Cost of solid culture per sample | Taken from de Almeida (https://www.ncbi.nlm.nih.gov/pmc/articles/PMC5309222/), with cost of $16.50 USD in 2013, inflated using GDP deflator - 97% inflation | 32.51 |
| Cost of first-line DST per panel | Cost provided by the National Tuberculosis Program (2023) and converted to USD at a rate of 0.197:1 | 183.46 |
| Cost of second-line DST per panel | Cost provided by the National Tuberculosis Program (2023) and converted to USD at a rate of 0.197:1 | 173.08 |
| Cost of first-line LPA per sample | Cost estimated by Figueredo (https://www.scielo.br/j/rsbmt/a/L5jBYQTqpgBFgc8vsQfqCxg/?format=pdf&lang=en), activity based costing and inflated using GDP deflator from 2022 to 2023 - 8.2% | 38.74 |
| Cost of second-line LPA per sample | Cost estimated by Figueredo (https://www.scielo.br/j/rsbmt/a/L5jBYQTqpgBFgc8vsQfqCxg/?format=pdf&lang=en), activity based costing and inflated using GDP deflator from 2022 to 2023 - 8.2% | 45.28 |
| Cost of pre-TPT evaluation, considering a treatment visit | Cost of treatment visit from federal reimbursement list | 4.36 |
| Cost of 6INH if complete, adults | Costs estimated based on monthly follow-up visits; drug costs = $7.15 | 16.58 |
| Cost of 6INH if complete, children | Costs estimated based on monthly follow-up visits; drug costs = $9.73 | 19.16 |
| Cost of 3HR if complete, adults | Costs estimated based on monthly follow-up visits; drug costs = $17.89 | 22.61 |
| Cost of 3HR if complete, children | Costs estimated based on monthly follow-up visits; drug costs = $9.80 | 14.51 |
| Cost of 3HP if complete, adults | Costs estimated based on monthly follow-up visits, with prorated DOT visits based on country utilization; drug costs = $24.18 | 36.74 |

| Cost Parameter | Description | Mean (USD 2023) |
|---|---|---|
| Cost of 3HP if complete, children | Costs estimated based on monthly follow-up visits, with prorated DOT visits based on country utilization; drug costs = $22.49 | 35.04 |
| Cost of 1HP if complete, adults | Costs estimated based on monthly follow-up visits; drug costs = $28.62 | 29.86 |
| Cost of 1HP if complete, children | Costs estimated based on monthly follow-up visits; drug costs = $21.57 | 22.8 |
| Cost of 6Lfx if complete, adults | Costs estimated based on monthly follow-up visits; drug costs = $39.36 | 48.78 |
| Cost of 6Lfx if complete, children | Costs estimated based on monthly follow-up visits; drug costs = $19.18 | 28.59 |
| Cost of 3HR if incomplete, adults; assumes 50% the price of complete | Assumed half the cost of complete treatment | 11.3 |
| Cost of 3HR if incomplete, children; assumes 50% the price of complete | Assumed half the cost of complete treatment | 7.25 |
| Cost of 6INH if incomplete, adults; assumes 50% the price of complete | Assumed half the cost of complete treatment | 8.29 |
| Cost of 6INH if incomplete, children; assumes 50% the price of complete | Assumed half the cost of complete treatment | 9.57 |
| Cost of 3HP if incomplete, adults; assumes 50% the price of complete | Assumed half the cost of complete treatment | 18.37 |
| Cost of 3HP if incomplete, children; assumes 50% the price of complete | Assumed half the cost of complete treatment | 17.52 |
| Cost of 1HP if incomplete, adults; assumes 50% the price of complete | Assumed half the cost of complete treatment | 14.93 |
| Cost of 1HP if incomplete, children; assumes 50% the price of complete | Assumed half the cost of complete treatment | 11.4 |
| Cost of 6Lfx if incomplete, adults; assumes 50% the price of complete | Assumed half the cost of complete treatment | 24.4 |
| Cost of 6Lfx if incomplete, children; assumes 50% the price of complete | Assumed half the cost of complete treatment | 14.3 |
| Cost of drug-susceptible TB treatment, adults | Drug costs reflective of those procured, plus, 23.3 days hospitalized (prorated for 14% hospitalization rate; https://www.ncbi.nlm.nih.gov/pmc/articles/PMC8332839/), 1.5 DOT doses per week (prorated), 1 follow-up per month, 1 smear per month, 3 cultures, and 1 chest x-ray every 4 months. Duration of treatment is 6 months. Cost of hospitalization day inflated from 2015 using GDP deflator from 38.40 to 64.90 | 566.99 |
| Cost of 4-month DS-TB treatment, children | Drug costs reflective of those procured, plus, 23.3 days hospitalized (prorated for 14% hospitalization rate), 1.5 DOT doses per week (prorated), 1 follow-up per month, 1 smear per month, 3 cultures, and 1 chest x-ray every 4 months. Duration of treatment is 4 months | 486.98 |
| Cost of 6-month DS-TB treatment, adults | Drug costs reflective of those procured, plus, 23.3 days hospitalized (prorated for 14% hospitalization rate), 1.5 DOT doses per week (prorated), 1 follow-up per month, 1 smear per month, 3 cultures, and 1 chest x-ray every 4 months. Duration of treatment is 6 months | 540.71 |
| Cost of BPaLM RR-TB treatment, adults (6 months bedaquiline, pretomanid, linezolid, moxifloxacin) | Drug costs reflective of those procured. Considers: 26 days hospitalized (on average), 1.5 DOT doses per week (prorated), 1 follow-up per month, 1 smear per month, 3 cultures, and 1 chest x-ray every 4 months. Duration of treatment is 6 months | 3105.25 |
| Cost of BPaLM RR-TB treatment, children (6 months bedaquiline, pretomanid, linezolid, moxifloxacin) | Drug costs reflective of those procured. Considers: 26 days hospitalized (on average), 1.5 DOT doses per week (prorated), 1 follow-up per month, 1 smear per month, 3 cultures, and 1 chest x-ray every 4 months | 4394.48 |
| Cost of 18-month all oral RR-TB treatment, adults (6 months bedaquiline, 18 months terizidone, linezolid, levofloxacin) | Drug costs reflective of those procured in the country. Considers: 26 days hospitalized (on average), 1.5 DOT doses per week (prorated), 1 follow-up per month, 1 smear per month, 3 cultures, and 1 chest x-ray every 4 months | 7494.76 |
| Cost of 18-month all oral RR-TB treatment, children (6 months bedaquiline, 18 months terizidone, linezolid, levofloxacin) | Drug costs reflective of those procured in the country. Considers: 26 days hospitalized (on average), 1.5 DOT doses per week (prorated), 1 follow-up per month, 1 smear per month, 3 cultures, and 1 chest x-ray every 4 months | 8797.9 |
| Patient costs per month of DS-TB treatment; mean episode costs were divided by average duration of treatment (6 months) | From the patient cost survey conducted in Brazil on a per month basis (assumed episode was 6 months). GDP deflator from 2021 to 2023 used - 17% inflation. | 281.23 |
| Patient costs per month of RR-TB treatment; mean episode costs were divided by average duration of treatment (18 months) | From the patient cost survey conducted in Brazil on a per month basis (assumed episode was 18 months). Since lower than DS-TB, assumed equivalent to DS-TB | 281.23 |
| Cost of post-tuberculosis healthcare utilization; considers 12.2 outpatient visits and 2.2 additional inpatient bed days | From Romanowski et al, considers 12.2 outpatient visits (health centre with beds) and 2.2 additional inpatient bed days (secondary hospital). Costs come from WHO-CHOICE in USD and inflated from 2008 using GDP deflator (187% increase) | 192.82 |
| Patient costs per month of TPT; assumed as a proportion of patient costs of DS-TB treatment (9.1%) | Assumed to be 9.09% of the DS-TB patient cost based on Shepardson et al. | 25.26 |

| Cost Parameter | Description | Mean (USD 2023) |
|---|---|---|
| Annual cost of healthcare worker training in intervention scenarios; annual recurrent 1-day training cost in intervention scenarios for training of 10% of healthcare workforce, plus TST training | Inclusive of annual costs for training 81 nurses on TST for 1.5 days per year, training 45,900 physicians and 118,000 nurses on TB 1 day per year, and paying a full-time nurse manager to oversee implementation per million population; plus $10,000 cost of a portion of a zoom license to carry out virtual training | 24,600,000 |
| Annual cost of active case finding training in years conducted for The Intervention Package; assumes 2 days training for all years of case finding in high-risk communities, applied as an annual implementation cost | Inclusive of training teams conducting case finding for 2 days - 81 corrections workers, 162 nurses, and 121 physicians | 77,000 |
| Annual cost of active case finding training in years conducted, enhanced scenario; assumes 2 days training for all years of case finding in high-risk communities, applied as an annual implementation cost | Inclusive of training teams conducting case finding for 2 days - 81 corrections workers, 324 nurses, and 121 physicians | 108,000 |
| Cost to screen one person during active case finding for The Intervention Package; considers costs of materials (cooler, ice pack, fuel, van), 50% of nights stayed out of town (hotel, food), personnel costs | 60 days of screening per year, with 81 teams. Each team has a van (annuitized at 10y lifespan) equipped with CXR, coolers, ice packs, fuel (prorated at $10,000 per 240 days), per diem for hotel and food for half the screening days. Cost includes the personnel trained above (prorated on a per team basis). Overall cost is a per person screened. | 10.25 |
| Cost to screen one person during active case finding, enhanced scenario; considers costs of materials (cooler, ice pack, fuel, van), 50% of nights stayed out of town (hotel, food), personnel costs, and cost of antigen-based skin test | 90 days of screening per year, with 81 teams. Each team has a van (annuitized at 10y lifespan) equipped with CXR, coolers, ice packs, fuel (prorated at $10,000 per 240 days), per diem for hotel and food for half the screening days. Costs also include the materials for TB antigen skin testing. Cost includes the personnel trained above (prorated on a per team basis). Overall cost is a per person screened. | 14.26 |

Abbreviations: 95% UR, 95% uncertainty range; TST, tuberculin skin test; IGRA, interferon-gamma release assay; HIV, human immunodeficiency virus; CRP, C-reactive protein; CXR, chest x-ray; CAD, computer aided detection; AFB, acid-fast bacilli; ZN, Ziehl-Nielsen; DST, drug susceptibility test; LPA, line probe assay; TPT, tuberculosis preventive treatment; 6INH, six-months of isoniazid daily; 3HR, three-months isoniazid and rifampicin daily; 3HP, three-months isoniazid and rifapentine weekly; 1HP, one-month isoniazid and rifapentine daily; 6Lfx, six months levofloxacin daily; DS-TB, drug-susceptible tuberculosis; RR-TB, rifampicin-resistant tuberculosis; BPaLM, bedaquiline, pretomanid, linezolid, and moxifloxacin; BPaLL, bedaquiline, pretomanid, linezolid, and levofloxacin.

**References this section**

1. Pinto M, Entringer AP, Steffen R, Trajman A. Cost analysis of nucleic acid amplification for diagnosing pulmonary tuberculosis, within the context of the Brazilian Unified Health Care System. J Bras Pneumol. 2015;41(6):536-8.
2. Romanowski K, Law MR, Karim ME, et al. Healthcare Utilization After Respiratory Tuberculosis: A Controlled Interrupted Time Series Analysis. Clin Infect Dis 2023; 77: 883–91.
3. Shepardson D, Marks SM, Chesson H, et al. Cost-effectiveness of a 12-dose regimen for treating latent tuberculous infection in the United States. Int J Tuberc Lung Dis 2013; 17: 1531–7.
4. WHO-CHOICE. WHO. https://www.who.int/choice/cost-effectiveness/inputs/en/ (accessed Feb 11, 2019).
5. Stop TB Partnership | Global Drug Facility - Products List. https://www.stoptb.org/facilitate-access-to-tb-drugs-diagnostics/global-drug-facility-gdf (accessed March 11, 2024).
6. Maciel ELN, Negri L dos SA, Guidoni LM, et al. The economic burden of households affected by tuberculosis in Brazil: First national survey results, 2019-2021. PLOS ONE 2023; 18: e0287961.
7. World Health Organization. WHO consolidated guidelines on tuberculosis: module 3: diagnosis: tests for TB infection. Geneva, Switzerland: WHO, 2022.
8. Alsdurf H, Oxlade O, Adjobimey M, et al. Resource implications of the latent tuberculosis cascade of care: a time and motion study in five countries. BMC Health Serv Res 2020; 20: 341.
9. de Almeida IN, de Assis Figueredo LJ, Soares VM, et al. Evaluation of the Mean Cost and Activity Based Cost in the Diagnosis of Pulmonary Tuberculosis in the Laboratory Routine of a High-Complexity Hospital in Brazil. Front Microbiol 2017; 8: 249.
10. Pinto M, Entringer AP, Steffen R, Trajman A. Cost analysis of nucleic acid amplification for diagnosing pulmonary tuberculosis, within the context of the Brazilian Unified Health Care System. J Bras Pneumol 2015; 41: 536–8.
11. Steffen RE, Pinto M, Kritski A, Trajman A. Cost-effectiveness of newer technologies for the diagnosis of Mycobacterium tuberculosis infection in Brazilian people living with HIV. Sci Rep 2020; 10: 21823.
12. Figueredo LJ de A, César ALA, Ferrazoli L, et al. Cost analysis of GenoType® MTBDRplus and GenoType® MTBDRsl at the State Laboratory of São Paulo, Brazil. Rev Soc Bras Med Trop 2023; 56: e0238-2023.
13. Cortez AO, de Melo AC, Neves L de O, Resende KA, Camargos P. Tuberculosis in Brazil: one country, multiple realities. J Bras Pneumol 2021; 47: e20200119.

#### Georgia

Costs were estimated in Georgia largely through the Value-TB project (Chikovani I, et al. *Int J Tuberc Lung Dis*. 2021;25(12):1019-27). All costs were reviewed by the national tuberculosis programme, who provided some additional costs (eg, costs of tuberculin). The IHT TB Tool (https://tb.integratedhealthtool.org/) was used to supplement costs of medications. As Georgia was the only country without a patient cost survey, we used estimates from Portnoy and colleagues' meta-regression (Portnoy A, et al. *Lancet Glob Health*. 2023;11(10):E1640-7). The composition of the 18-month rifampicin-resistant tuberculosis regimen was assumed to be bedaquiline (six months), clofazimine, linezolid, and levofloxacin. We assumed 88% of all individuals receiving tuberculosis preventive treatment received rifamycin-based regimens and 67% of all individuals treated for rifampicin-resistant tuberculosis received short regimens in 2024. These values remained the same in status quo scenarios, but increased to 100% by 2030 with The Intervention Package.

We worked with the national tuberculosis programme to define the high-risk community—in this case, people accessing care for injection drug use. Based on a population size of 17,556 and a target to screen on 60% of this population, we estimated 1 team composed of one nurse, one physician, and one lab technician would be deployed per region (11 regions) to conduct this screening at a rate of 80 people screened per day. The screening would take 12 days to complete. For each team, we assumed they would have a mobile van fully equipped with chest x-ray, with an annual, annuitized cost of $55,100 USD, they would require $200 USD in fuel costs, $290 in coolers and ice to transport sputum samples, 6 days in hotel (at a cost of $57 per night), and a per diem of $12 per person per day. Complete costs are described below.

| Cost Parameter | Description | Mean (USD 2023) |
|---|---|---|
| Cost to perform symptom screening per person | Cost of an outpatient screening visit, VALUE-TB, inflated using GDP deflator 2018-2023 - 40.7% | 3.76 |
| Cost of household contact investigation per index patient | Taken from Value TB pooled dataset for contact tracing (per patient episode); geometric mean from of the contact tracing costs at two facilities reporting such data. Original data in 2018 was USD$11.15, adjusted to 2023 at 40.7% | 15.69 |
| Cost of TST, inclusive of materials and personnel time | Value TB study - value of $3.61 in 2018 - adjusted to 2023 @ 40.7% | 5.08 |
| Cost of IGRA, inclusive of materials | From National TB Program: Cost of service is 44 lari, converted at 0.37 USD per Lari, plus cost of $900 USD for 192 units in 2023. | 21.02 |
| Cost of antigen-based skin test, inclusive of materials and personnel time | National TB Program reports cost of tuberculin to be $1.54 per dose and one dose of dia skin test is $1.60 USD. Assume all other costs identical | 5.14 |
| Cost of Xpert per person | Value TB Study - value of $14.75 - adjusted to 2023 @ 40.7% and incorporating $2 reduction in cartridge cost | 18.75 |
| Cost of HIV test per person | Value TB Study - value of $2.86 - adjusted to 2023 @ 40.7% | 4.02 |
| Cost of CRP per person | Assumed same cost as HIV test | 4.02 |
| Cost of CXR per person | Digital CXR cost from Value TB study - value of $2.14 - adjusted to 2023 @40.7% | 3.01 |
| Annual cost of CAD license (annuitized); considered in analysis as a new investment and part of implementation in 5% of all healthcare facilities | InferRead CAD software (one license), with annual costs and annuitized capital costs at 3% over 10 years for software and installation/training, 5 years for laptop, and 3 years for initial maintenance agreement with the next years costing $250 per annum (GDF price catalogue) | 1058 |
| Cost of AFB smear, ZN | Value TB Study for ZN microscopy - value of $4.18 - adjusted to 2023 @ 40.7% | 5.88 |
| Cost of sputum collection per person | Value TB study - value of 1.77 - adjusted to 2023 @ 40.7% | 2.49 |
| Cost of liquid culture per sample | Value TB study - value of 15.67 - adjusted to 2023 @ 40.7% | 22.05 |
| Cost of solid culture per sample | Value TB study - value of 8.63 - adjusted to 2023 @ 40.7% | 12.14 |
| Cost of first-line DST per panel | Value TB study, first line LIQUID - value of 19.82 - adjusted to 2023 @ 40.7% | 27.89 |
| Cost of second-line DST per panel | Value TB study, second line LIQUID - value of 12.60 - adjusted to 2023 @ 40.7% | 17.73 |
| Cost of first-line LPA per sample | Value TB study, LPA - value of 63.20 - adjusted to 2023 @ 40.7% | 88.92 |
| Cost of second-line LPA per sample | Assumed same cost of LPA 1 | 88.92 |
| Cost of pre-TPT evaluation, considering a treatment visit | Value TB study, cost of a treatment visit - value of 2.12 - adjusted to 2023 @ 40.7% | 2.98 |
| Cost of 6INH if complete, adults | Monthly visits assumed (cost of $2.98 in 2023 USD) = 17.88; and medication: 10.80 in 2021, inflated to 12.21 (13.1%, GDP deflator) in 2023; meds from IHT TB tool; plus 10% the cost of liver function test (assumed equivalent cost to electrolyte test, $2.54 in 2018 and 3.57 in 2023 = $0.36 | 30.45 |
| Cost of 6INH if complete, children | Monthly visits assumed (cost of $2.98 in 2023 USD) = 17.88; and medication: 32.40 in 2021, inflated to 36.64 in 2023 (13.1% GDP deflator); meds from IHT TB tool; plus 10% the cost of liver function test (assumed equivalent cost to electrolyte test, $2.54 in 2018 and 3.57 in 2023 = $0.36 | 54.88 |
| Cost of 3HR if complete, adults | Monthly visits assumed (cost of $2.98 in 2023 USD) = 8.94; and medication: 18 in 2021, inflated to 20.36 in 2023 (13.1% GDP deflator); meds from IHT TB tool; plus 10% the cost of liver function test (assumed equivalent cost to electrolyte test, $2.54 in 2018 and 3.57 in 2023 = $0.36 | 29.66 |
| Cost of 3HR if complete, children | Monthly visits assumed (cost of $2.98 in 2023 USD) = 8.94 and medication: 21.60 in 2021, inflated to 24.43 in 2021 (13.1% GDP deflator); meds from IHT TB tool; plus 10% the cost of liver function test (assumed equivalent cost to electrolyte test, $2.54 in 2018 and 3.57 in 2023 = $0.36 | 33.73 |
| Cost of 3HP if complete, adults | Monthly visits assumed (cost of $2.98 in 2023 USD) = 8.94. VOT for 6.5 doses (prorated at 81% of the 8 remaining doses) and DOT for 1.5 doses. VOT cost is 3.08 (2.19*40.7% inflation) based on Value TB; DOT costs assumed same as treatment costs ($2.98); Therefore VOT/DOT costs are $24.49. Medication: 14.40 in 2021, inflated to 16.29 in 2023 (13.1% GDP deflator); meds from IHT TB tool; plus 10% the cost of liver function test (assumed equivalent cost to electrolyte test, $2.54 in 2018 and 3.57 in 2023 = $0.36 | 50.08 |
| Cost of 3HP if complete, children | Monthly visits assumed (cost of $2.98 in 2023 USD) = 8.94. VOT for 6.5 doses (prorated at 81% of the 8 remaining doses) and DOT for 1.5 doses. VOT cost is 3.08 | 50.08 |

| Cost Parameter | Description | Mean (USD 2023) |
|---|---|---|
| | (2.19*40.7% inflation) based on Value TB; DOT costs assumed same as treatment costs ($2.98); Therefore VOT/DOT costs are $24.49. Medication:14.40 in 2021, inflated to 16.29 in 2023 (13.1% GDP deflator); meds from IHT TB tool; plus 10% the cost of liver function test (assumed equivalent cost to electrolyte test, $2.54 in 2018 and 3.57 in 2023 = $0.36 | |
| Cost of 1HP if complete, adults | One treatment visit assumed ($2.98) and medication: $12.88 in 2021, inflated to 14.57 in 2023 (13.1% GDP deflator); meds from IHT TB tool; plus 10% the cost of liver function test (assumed equivalent cost to electrolyte test, $2.54 in 2018 and 3.57 in 2023 = $0.36 | 17.91 |
| Cost of 1HP if complete, children | One treatment visit assumed ($2.98) and medication: 20.10 in 2021, inflated to 22.73 in 2023 (13.1% GDP deflator); meds from IHT TB tool; plus 10% the cost of liver function test (assumed equivalent cost to electrolyte test, $2.54 in 2018 and 3.57 in 2023 = $0.36 | 26.07 |
| Cost of 6Lfx if complete, adults | Monthly visits assumed (cost of $2.98 in 2023 USD) = 17.88 and medication: 18, inflated to 20.36 in 2023 (13.1% GDP deflator); meds from IHT TB tool; plus 10% the cost of liver function test (assumed equivalent cost to electrolyte test, $2.54 in 2018 and 3.57 in 2023 = $0.36 | 38.6 |
| Cost of 6Lfx if complete, children | Monthly visits assumed (cost of $2.98 in 2023 USD) = 17.88 and medication: 16.20, inflated to 18.32 in 2023 (13.1% GDP); meds from IHT TB tool; plus 10% the cost of liver function test (assumed equivalent cost to electrolyte test, $2.54 in 2018 and 3.57 in 2023 = $0.36 | 36.56 |
| Cost of 3HR if incomplete, adults; assumes 50% the price of complete | Assumed half the cost of complete treatment, other than LFT costs, which remain ($0.36) | 15.01 |
| Cost of 3HR if incomplete, children; assumes 50% the price of complete | Assumed half the cost of complete treatment, other than LFT costs, which remain ($0.36) | 17.045 |
| Cost of 6INH if incomplete, adults; assumes 50% the price of complete | Assumed half the cost of complete treatment, other than LFT costs, which remain ($0.36) | 15.405 |
| Cost of 6INH if incomplete, children; assumes 50% the price of complete | Assumed half the cost of complete treatment, other than LFT costs, which remain ($0.36) | 27.62 |
| Cost of 3HP if incomplete, adults; assumes 50% the price of complete | Assumed half the cost of complete treatment, other than LFT costs, which remain ($0.36) | 25.22 |
| Cost of 3HP if incomplete, children; assumes 50% the price of complete | Assumed half the cost of complete treatment, other than LFT costs, which remain ($0.36) | 25.22 |
| Cost of 1HP if incomplete, adults; assumes 50% the price of complete | Assumed half the cost of complete treatment, other than LFT costs, which remain ($0.36) | 9.135 |
| Cost of 1HP if incomplete, children; assumes 50% the price of complete | Assumed half the cost of complete treatment, other than LFT costs, which remain ($0.36) | 13.215 |
| Cost of 6Lfx if incomplete, adults; assumes 50% the price of complete | Assumed half the cost of complete treatment, other than LFT costs, which remain ($0.36) | 19.48 |
| Cost of 6Lfx if incomplete, children; assumes 50% the price of complete | Assumed half the cost of complete treatment, other than LFT costs, which remain ($0.36) | 18.46 |
| Cost of drug-susceptible TB treatment, adults | Value TB Study - First Line TB Treatment from the Value TB Dataset for adult pulmonary TB patients; geometric mean of 231.24 - adjusted to 2023 @ 40.7%; 6 months duration | 325.35 |
| Cost of 4-month DS-TB treatment, children | Estimated from the 6 month regimen, taking difference in drug costs from the IHT-TB tool (55.20 - 38.40 = drug cost reduction of 16.80). Remaining non-drug costs are: 611.12 - 62.43 ( from GDP deflator 2021-2023 = 13.1%) = 548.69 or 91.45 per month. So costs of 4 month short child regimen is 365.80 + 16.8*1.131 | 384.81 |
| Cost of 6-month DS-TB treatment, adults | Value TB Study - First Line TB Treatment from the Value TB Dataset for adult pulmonary TB patients; geometric mean of 434.34 - adjusted to 2023 @ 40.7%; 6 months duration | 611.12 |
| Cost of BPaLM RR-TB treatment, adults (6 months bedaquiline, pretomanid, linezolid, moxifloxacin) | From IHT Integrated Health Tool. Drug costs are equivalent to 745.20 in 2021; GDP deflator of 13.1%. Other service level costs from Sweeney et al in 2019USD of 1149.29; GDP deflator of 33.7%. | 2379.42 |
| Cost of BPaLM RR-TB treatment, children (6 months bedaquiline, pretomanid, linezolid, moxifloxacin) | From IHT Integrated Health Tool. Drug costs are assumed equivalent to 95% that of adults (based on GDF prices, 2023); GDP deflator of 13.1%. Other service level costs from Sweeney et al (assume same costs in child) in 2019USD of 1149.29; GDP deflator of 33.7%. | 2337.28 |
| Cost of 18-month all oral RR-TB treatment, adults (6 months bedaquiline, 18 months clofazimine, linezolid, levofloxacin) | From IHT Integrated Health Tool for drug costs: 1506.60 plus GDP deflator of 13.1% = 1703.96. Other service level costs from Sweeney et al in 2019USD of 1243.73; GDP deflator of 33.7% | 3366.83 |
| Cost of 18-month all oral RR-TB treatment, children (6 months bedaquiline, 18 months clofazimine, linezolid, levofloxacin) | From IHT Integrated Health tool for drug costs: 1782 plus GDP deflator of 13.1%. Other service level costs from Sweeney et al (assume same costs in child) in 2019USD of 1243.73; GDP deflator of 33.7%. | 3678.31 |
| Patient costs per month of DS-TB treatment; mean episode costs were divided by average duration of treatment (6 months) | From Portnoy and colleagues meta-regression on a per month basis (assumed episode was 6 months). GDP deflator from 2021 to 2023 used - 13.1% inflation on a value of 366.50 per month | 414.51 |
| Patient costs per month of RR-TB treatment; mean episode costs were divided by average duration of treatment (18 | From Portnoy and colleagues meta-regression on a per month basis (assumed episode was 18 months). Since original estimate of 329.33 was lower than DS-TB, assumed equivalent to DS-TB | 414.51 |

| Cost Parameter | Description | Mean (USD 2023) |
|---|---|---|
| months) | | |
| Cost of post-tuberculosis healthcare utilization; considers 12.2 outpatient visits and 2.2 additional inpatient bed days | From Romanowski et al, considers 12.2 outpatient visits (health centre with beds) and 2.2 additional inpatient bed days (secondary hospital). Costs come from WHO-CHOICE in USD and inflated from 2008 using GDP deflator (123% increase) | 284.16 |
| Patient costs per month of TPT; assumed as a proportion of patient costs of DS-TB treatment (9.1%) | Assumed to be 9.09% of the DS-TB patient cost based on Shepardson et al. | 37.68 |
| Annual cost of healthcare worker training in intervention scenarios; annual recurrent 1-day training cost in intervention scenarios for training of 10% of healthcare workforce, plus TST training | Inclusive of annual costs for training 11 nurses on TST for 1.5 days per year, training 279 physicians and 200 nurses on TB 1 day per year, and paying a full time nurse manager to oversee implementation per million population; plus $10,000 cost of a portion of a zoom license to carry out virtual training | 330,000 |
| Annual cost of active case finding training in years conducted for The Intervention Package; assumes 2 days training for all years of case finding in high-risk communities, applied as an annual implementation cost | Inclusive of training teams conducting case finding for 2 days - 11 lab technicians, 11 nurses, and 11 physicians | 15,000 |
| Annual cost of active case finding training in years conducted, enhanced scenario; assumes 2 days training for all years of case finding in high-risk communities, applied as an annual implementation cost | Inclusive of training teams conducting case finding for 2 days - 11 lab technicians, 22 nurses, and 11 physicians | 18,000 |
| Cost to screen one person during active case finding for The Intervention Package; considers costs of materials (cooler, ice pack, fuel, van), 50% of nights stayed out of town (hotel, food), personnel costs | 12 days of screening per year, with 11 teams. Each team has a van (annuitized at 10y lifespan) equipped with CXR, coolers, ice packs, fuel (prorated at $10,000 per 240 days), per diem for hotel and food for half the screening days. Cost includes the personnel trained above (prorated on a per team basis). Overall cost is a per person screened. | 13.14 |
| Cost to screen one person during active case finding, enhanced scenario; considers costs of materials (cooler, ice pack, fuel, van), 50% of nights stayed out of town (hotel, food), personnel costs, and cost of antigen-based skin test | 18 days of screening per year, with 11 teams. Each team has a van (annuitized at 10y lifespan) equipped with CXR, coolers, ice packs, fuel (prorated at $10,000 per 240 days), per diem for hotel and food for half the screening days. Costs also include the materials for TB antigen skin testing. Cost includes the personnel trained above (prorated on a per team basis). Overall cost is a per person screened. | 16.67 |

Abbreviations: 95% UR, 95% uncertainty range; TST, tuberculin skin test; IGRA, interferon-gamma release assay; HIV, human immunodeficiency virus; CRP, C-reactive protein; CXR, chest x-ray; CAD, computer aided detection; AFB, acid-fast bacilli; ZN, Ziehl-Nielsen; DST, drug susceptibility test; LPA, line probe assay; TPT, tuberculosis preventive treatment; 6INH, six-months of isoniazid daily; 3HR, three-months isoniazid and rifampicin daily; 3HP, three-months isoniazid and rifapentine weekly; 1HP, one-month isoniazid and rifapentine daily; 6Lfx, six months levofloxacin daily; DS-TB, drug-susceptible tuberculosis; RR-TB, rifampicin-resistant tuberculosis; BPaLM, bedaquiline, pretomanid, linezolid, and moxifloxacin; BPaLL, bedaquiline, pretomanid, linezolid, and levofloxacin.

**References this section**

1. IHT. Integrated Health Tool: TB. 2023. https://tb.integratedhealthtool.org/ (accessed March 12, 2024).
2. Portnoy A, Yamanaka T, Nguhiu P, et al. Costs incurred by people receiving tuberculosis treatment in low-income and middle-income countries: a meta-regression analysis. The Lancet Global Health 2023; 11: e1640–7.
3. Chikovani I, Shengelia N, Marjanishvili N, et al. Cost of TB services in the public and private sectors in Georgia. Int J Tuberc Lung Dis 2021; 25: 1019–27.
4. Romanowski K, Law MR, Karim ME, et al. Healthcare Utilization After Respiratory Tuberculosis: A Controlled Interrupted Time Series Analysis. Clin Infect Dis 2023; 77: 883–91.
5. Shepardson D, Marks SM, Chesson H, et al. Cost-effectiveness of a 12-dose regimen for treating latent tuberculous infection in the United States. Int J Tuberc Lung Dis 2013; 17: 1531–7.
6. WHO-CHOICE. WHO. https://www.who.int/choice/cost-effectiveness/inputs/en/ (accessed Feb 11, 2019).
7. Stop TB Partnership | Global Drug Facility - Products List. https://www.stoptb.org/facilitate-access-to-tb-drugs-diagnostics/global-drug-facility-gdf (accessed March 11, 2024).
8. World Health Organization. WHO consolidated guidelines on tuberculosis: module 3: diagnosis: tests for TB infection. Geneva, Switzerland: WHO, 2022.
9. Sweeney S, Berry C, Kazounis E, et al. Cost-effectiveness of short, oral treatment regimens for rifampicin resistant tuberculosis. PLOS Global Public Health 2022; 2: e0001337.

#### Kenya

Costs were estimated in Kenya largely through the Value-TB project (Kairu A, et al. *Int J Tuberc Lung Dis*. 2021;25(12):1028-34). All costs were reviewed by the national tuberculosis programme and experts in TB finance, who provided some additional costs (eg, costs of medications). The IHT TB Tool (https://tb.integratedhealthtool.org/) was used to supplement some costs of medications. The composition of the 18-month rifampicin-resistant tuberculosis regimen was assumed to be bedaquiline (six months), linezolid (six months), clofazimine, cycloserine, and levofloxacin. We assumed 8% of all individuals receiving tuberculosis preventive treatment received rifamycin-based regimens and 5% of all individuals treated for rifampicin-resistant tuberculosis received short regimens in 2024. These values increased to 50% by 2030 in status quo scenarios, but increased to 100% by 2030 with The Intervention Package.

We worked with the national tuberculosis programme to define the high-risk community—in this case, people living in slums in nine high prevalence counties. Based on a population size of 416,250 and a target to screen on 60% of this population, we estimated 4 teams composed of one nurse, one physician, and one lab technician would be deployed per county (9 counties) to conduct this screening at a rate of 80 people screened per day. The screening would take 87 days to complete. For each team, we assumed they would have a mobile van fully equipped with chest x-ray, with an annual, annuitized cost of $55,100 USD, they would require $1090 USD in fuel costs, $290 in coolers and ice to transport sputum samples, 44 days in hotel (at a cost of $70 per night), and a per diem of $17 per person per day. Complete costs are described below.

| Cost Parameter | Description | Mean (USD 2023) |
|---|---|---|
| Cost to perform symptom screening per person | Cost of an outpatient screening visit, VALUE-TB, inflated using GDP deflator 2018-2023 - 30.5% | 2.98 |
| Cost of household contact investigation per index patient | Taken from VALUE-TB. Contact tracing involved phone call and home visit. Original data in 2018 was USD$13.54, adjusted to 2023 at 30.5% | 17.67 |
| Cost of TST, inclusive of materials and personnel time | Estimated based on cost of tuberculin from Georgia ($1.54) plus cost of screening and diagnostic visit in 2018 from VALUE-TB. Visits inflated using deflator at 30.5% | 10.28 |
| Cost of IGRA, inclusive of materials | Not used in Kenya; estimated based on average GDF procurement prices (17.44 in 2023) and service level costs from Georgia, adjusted using PPP @ 46.6 = 2050 KEL in 2023 and converted to USD at 0.0066 USD per KEL = 13.62 | 30.06 |
| Cost of antigen-based skin test, inclusive of materials and personnel time | Based cost of tuberculin to be $1.54 per dose and one dose of dia skin test is $1.60 USD. Assume all other costs identical | 10.34 |
| Cost of Xpert per person | Value TB Study - value of $16.79 - adjusted to 2023 @ 30.5% and incorporating $2 reduction in cartridge cost | 19.91 |
| Cost of HIV test per person | Value TB Study - value of $5.86 - adjusted to 2023 @ 30.5% | 7.64 |
| Cost of CRP per person | Assumed same cost as HIV test | 7.64 |
| Cost of CXR per person | Digital CXR cost from Value TB study - value of $23.55 - adjusted to 2023 @30.5% | 30.73 |
| Annual cost of CAD license (annuitized); considered in analysis as a new investment and part of implementation in 5% of all healthcare facilities | InferRead CAD software (one license), with annual costs and annuitized capital costs at 3% over 10 years for software and installation/training, 5 years for laptop, and 3 years for initial maintenance agreement with the next years costing $250 per annum (GDF price catalogue) | 1058 |
| Cost of AFB smear, ZN | Value TB Study for ZN microscopy - value of $13.36 - adjusted to 2023 @ 30.5% | 17.43 |
| Cost of sputum collection per person | Value TB study - value of 6.26 - adjusted to 2023 @ 30.5% | 8.17 |
| Cost of liquid culture per sample | Value TB study - value of 26.96 - adjusted to 2023 @ 30.5% | 35.19 |
| Cost of solid culture per sample | Value TB study - value of 35.58 - adjusted to 2023 @ 30.5% | 46.43 |
| Cost of first-line DST per panel | Value TB study, first line LIQUID - value of 40.42 - adjusted to 2023 @ 30.5% | 52.75 |
| Cost of second-line DST per panel | Estimated based on relative difference in first and second line DST in Georgia (63.57% of the cost) | 33.53 |
| Cost of first-line LPA per sample | Value TB study, LPA - value of 47.50 - adjusted to 2023 @ 30.5% | 61.99 |
| Cost of second-line LPA per sample | Assumed same cost of LPA first line. | 61.99 |
| Cost of pre-TPT evaluation, considering a treatment visit | Value TB study, cost of a treatment visit - value of 4.10 - adjusted to 2023 @ 30.5% | 5.35 |
| Cost of 6INH if complete, adults | Monthly visits assumed, costs of each visit in 2018 from Value-TB dataset was estimated to be $3.64 using geometric mean; in 2023 at 30.5% inflation = $4.75 per visit. Costs of medications from projections of TB commodity costs by TB Program. Medication costs = $6.72 | 35.22 |
| Cost of 6INH if complete, children | Monthly visits assumed, costs of each visit in 2018 from Value-TB dataset was estimated to be $3.64 using geometric mean; in 2023 at 30.5% inflation = $4.75 per visit. Costs of medications from projections of TB commodity costs by TB Program. Medication costs = $45.36 | 73.86 |
| Cost of 3HR if complete, adults | Monthly visits assumed, costs of each visit in 2018 from Value-TB dataset was estimated to be $3.64 using geometric mean; in 2023 at 30.5% inflation = $4.75 per visit. Costs from IHT tool ($18 in 2021) and inflated at 14.3% = 20.57 | 34.82 |
| Cost of 3HR if complete, children | Monthly visits assumed, costs of each visit in 2018 from Value-TB dataset was estimated to be $3.64 using geometric mean; in 2023 at 30.5% inflation = $4.75 per visit. Costs of medications from projections of TB commodity costs by TB Program. Medication costs of $13.44 | 27.69 |
| Cost of 3HP if complete, adults | Monthly visits assumed, costs of each visit in 2018 from Value-TB dataset was estimated to be $3.64 using geometric mean; in 2023 at 30.5% inflation = $4.75 per visit. DOT not provided by health system, but family. Cost of medication is $14.40 from TB commodity costs by TB Program | 28.65 |
| Cost of 3HP if complete, children | Monthly visits assumed, costs of each visit in 2018 from Value-TB dataset was estimated to be $3.64 using geometric mean; in 2023 at 30.5% inflation = $4.75 per visit. DOT not provided by health system, but family. Cost of medication is $14.40 from TB commodity costs by TB Program | 28.65 |

| Cost Parameter | Description | Mean (USD 2023) |
|---|---|---|
| Cost of 1HP if complete, adults | Monthly visits assumed, costs of each visit in 2018 from Value-TB dataset was estimated to be $3.64 using geometric mean; in 2023 at 30.5% inflation = $4.75 per visit. Medication costs from IHT tool = $20.72 and inflated at 14.3% = 23.68 | 28.43 |
| Cost of 1HP if complete, children | Monthly visits assumed, costs of each visit in 2018 from Value-TB dataset was estimated to be $3.64 using geometric mean; in 2023 at 30.5% inflation = $4.75 per visit. Medication costs from IHT tool = $26.60 and inflated at 14.3% = 30.40 | 35.15 |
| Cost of 6Lfx if complete, adults | Monthly visits assumed, costs of each visit in 2018 from Value-TB dataset was estimated to be $3.64 using geometric mean; in 2023 at 30.5% inflation = $4.75 per visit. Medication costs from TB commodity costs by TB program at $14.04 | 42.54 |
| Cost of 6Lfx if complete, children | Monthly visits assumed, costs of each visit in 2018 from Value-TB dataset was estimated to be $3.64 using geometric mean; in 2023 at 30.5% inflation = $4.75 per visit. Medication costs from TB commodity costs by TB program at $8.02 | 36.52 |
| Cost of 3HR if incomplete, adults; assumes 50% the price of complete | Assumed half the cost of complete treatment | 17.41 |
| Cost of 3HR if incomplete, children; assumes 50% the price of complete | Assumed half the cost of complete treatment | 13.85 |
| Cost of 6INH if incomplete, adults; assumes 50% the price of complete | Assumed half the cost of complete treatment | 17.61 |
| Cost of 6INH if incomplete, children; assumes 50% the price of complete | Assumed half the cost of complete treatment | 36.93 |
| Cost of 3HP if incomplete, adults; assumes 50% the price of complete | Assumed half the cost of complete treatment | 14.33 |
| Cost of 3HP if incomplete, children; assumes 50% the price of complete | Assumed half the cost of complete treatment | 14.33 |
| Cost of 1HP if incomplete, adults; assumes 50% the price of complete | Assumed half the cost of complete treatment | 14.22 |
| Cost of 1HP if incomplete, children; assumes 50% the price of complete | Assumed half the cost of complete treatment | 17.58 |
| Cost of 6Lfx if incomplete, adults; assumes 50% the price of complete | Assumed half the cost of complete treatment | 21.27 |
| Cost of 6Lfx if incomplete, children; assumes 50% the price of complete | Assumed half the cost of complete treatment | 18.26 |
| Cost of drug-susceptible TB treatment, adults | Value TB Study - First Line TB Treatment for new and relapse PTB = 146.52 - inflated at 30.5%; 6 months duration | 191.21 |
| Cost of 4-month DS-TB treatment, children | Estimated from 6 month regimen, removing all medication costs (estimated to be 79.13 by TB program in 2023); then only including 4 visits = $71.50 in visits. Medication costs for 4 months = 62.49 | 133.99 |
| Cost of 6-month DS-TB treatment, adults | Value TB Study - First Line TB Treatment for new and relapse PTB = 142.82 - inflated at 30.5%; 6 months duration | 186.38 |
| Cost of BPaLM RR-TB treatment, adults (6 months bedaquiline, pretomanid, linezolid, moxifloxacin) | Value TB Study - Second Line TB Treatment, Adults, Long Regimen. Total cost of non-drug related items was equivalent to 4197.32 USD or 233.18 USD per month assuming 6 month duration. Inflated at 30.5% to 304.31 per month. Drug costs from the TB commodity costs by TB program at: 624.40 | 3385.99 |
| Cost of BPaLM RR-TB treatment, children (6 months bedaquiline, pretomanid, linezolid, moxifloxacin) | Value TB Study - Second Line TB Treatment, Adults, Long Regimen. Total cost of non-drug related items was equivalent to 4197.32 USD or 233.18 USD per month assuming 18 month duration. Inflated at 30.5% to 304.31 per month. Drug costs from the TB commodity costs by TB program at: 540.06 | 3301.65 |
| Cost of 18-month all oral RR-TB treatment, adults (6 months bedaquiline, 18 months cycloserine, clofazimine, linezolid, levofloxacin) | Value TB Study - Second Line TB Treatment, Adults, Long Regimen. Total cost of non-drug related items was equivalent to 4197.32 USD or 233.18 USD per month assuming 18 month duration. Inflated at 30.5% to 304.31 per month. Drug costs from the TB commodity costs by TB program at: 1859.13 | 7336.71 |
| Cost of 18-month all oral RR-TB treatment, children (6 months bedaquiline, 18 months cycloserine, clofazimine, linezolid, levofloxacin) | Value TB Study - Second Line TB Treatment, Adults, Long Regimen. Total cost of non-drug related items was equivalent to 4197.32 USD or 233.18 USD per month assuming 18 month duration. Inflated at 30.5% to 304.31 per month. Drug costs from the TB commodity costs by TB program at: 2460.2 | 7937.78 |
| Patient costs per month of DS-TB treatment; mean episode costs were divided by average duration of treatment (6 months) | Costs per patient month estimated from patient cost survey assuming 6 months treatment to be 4310 KEL. Exchange rate in 2017 = 0.0097; GDP deflator = 36.7% | 57.15 |
| Patient costs per month of RR-TB treatment; mean episode costs were divided by average duration of treatment (18 months) | Costs per patient month estimated from patient cost survey assuming 18 months treatment to be 8060 KEL. Exchange rate in 2017 = 0.0097; GDP deflator = 36.7% | 106.87 |
| Cost of post-tuberculosis healthcare utilization; considers 12.2 outpatient visits and 2.2 additional inpatient bed days | From Romanowski et al, considers 12.2 outpatient visits (health centre with beds) and 2.2 additional inpatient bed days (secondary hospital). Costs come from WHO-CHOICE in USD and inflated from 2008 using GDP deflator (206% increase) | 102.62 |
| Patient costs per month of TPT; assumed as a proportion of patient costs of DS-TB treatment (9.1%) | Assumed to be 9.09% of the DS-TB patient cost based on Shepardson et al. | 5.19 |

| Cost Parameter | Description | Mean (USD 2023) |
|---|---|---|
| Annual cost of healthcare worker training in intervention scenarios; annual recurrent 1-day training cost in intervention scenarios for training of 10% of healthcare workforce, plus TST training | Inclusive of annual costs for training 72 nurses on TST for 1.5 days per year, training 1200 physicians and 6400 nurses on TB 1 day per year, and paying a full time nurse manager to oversee implementation per million population; plus $10,000 cost of a portion of a zoom license to carry out virtual training | 734,000 |
| Annual cost of active case finding training in years conducted for The Intervention Package; assumes 2 days training for all years of case finding in high-risk communities, applied as an annual implementation cost | Inclusive of training teams conducting case finding for 2 days - 36 lab technicians, 36 nurses, and 36 physicians | 8000 |
| Annual cost of active case finding training in years conducted, enhanced scenario; assumes 2 days training for all years of case finding in high-risk communities, applied as an annual implementation cost | Inclusive of training teams conducting case finding for 2 days - 36 lab technicians, 72 nurses, and 36 physicians | 10,000 |
| Cost to screen one person during active case finding for The Intervention Package; considers costs of materials (cooler, ice pack, fuel, van), 50% of nights stayed out of town (hotel, food), personnel costs | 87 days of screening per year, with 36 teams. Each team has a van (annuitized at 10y lifespan) equipped with CXR, coolers, ice packs, fuel (prorated at $10,000 per 240 days), per diem for hotel and food for half the screening days. Cost includes the personnel trained above (prorated on a per team basis). Overall cost is a per person screened. | 6.12 |
| Cost to screen one person during active case finding, enhanced scenario; considers costs of materials (cooler, ice pack, fuel, van), 50% of nights stayed out of town (hotel, food), personnel costs, and cost of antigen-based skin test | 130 days of screening per year, with 36 teams. Each team has a van (annuitized at 10y lifespan) equipped with CXR, coolers, ice packs, fuel (prorated at $10,000 per 240 days), per diem for hotel and food for half the screening days. Costs also include the materials for TB antigen skin testing. Cost includes the personnel trained above (prorated on a per team basis). Overall cost is a per person screened. | 8.12 |

Abbreviations: 95% UR, 95% uncertainty range; TST, tuberculin skin test; IGRA, interferon-gamma release assay; HIV, human immunodeficiency virus; CRP, C-reactive protein; CXR, chest x-ray; CAD, computer aided detection; AFB, acid-fast bacilli; ZN, Ziehl-Nielsen; DST, drug susceptibility test; LPA, line probe assay; TPT, tuberculosis preventive treatment; 6INH, six-months of isoniazid daily; 3HR, three-months isoniazid and rifampicin daily; 3HP, three-months isoniazid and rifapentine weekly; 1HP, one-month isoniazid and rifapentine daily; 6Lfx, six months levofloxacin daily; DS-TB, drug-susceptible tuberculosis; RR-TB, rifampicin-resistant tuberculosis; BPaLM, bedaquiline, pretomanid, linezolid, and moxifloxacin; BPaLL, bedaquiline, pretomanid, linezolid, and levofloxacin.

**References this section**

1. IHT. Integrated Health Tool: TB. 2023. https://tb.integratedhealthtool.org/ (accessed March 12, 2024).
2. Kairu A, Orangi S, Oyando R, et al. Cost of TB services in healthcare facilities in Kenya. Int J Tuberc Lung Dis 2021; 25: 1028–34.
3. Kenya Ministry of Health. The first Kenya Tuberculosis Patient Cost Survey, 2017. Kenya: Kenya MoH, 2018.
4. Romanowski K, Law MR, Karim ME, et al. Healthcare Utilization After Respiratory Tuberculosis: A Controlled Interrupted Time Series Analysis. Clin Infect Dis 2023; 77: 883–91.
5. Shepardson D, Marks SM, Chesson H, et al. Cost-effectiveness of a 12-dose regimen for treating latent tuberculous infection in the United States. Int J Tuberc Lung Dis 2013; 17: 1531–7.
6. WHO-CHOICE. WHO. https://www.who.int/choice/cost-effectiveness/inputs/en/ (accessed Feb 11, 2019).
7. Stop TB Partnership | Global Drug Facility - Products List. https://www.stoptb.org/facilitate-access-to-tb-drugs-diagnostics/global-drug-facility-gdf (accessed March 11, 2024).
8. World Health Organization. WHO consolidated guidelines on tuberculosis: module 3: diagnosis: tests for TB infection. Geneva, Switzerland: WHO, 2022.

#### South Africa

Costs were estimated in South Africa leveraging costs largely collected through a cost exercise part of national tuberculosis strategic planning. All costs were reviewed by the national tuberculosis programme and those involved in costing the strategic plan, who provided additional, unpublished costs from the South Africa patient cost survey. The composition of the 18-month rifampicin-resistant tuberculosis regimen was assumed to be bedaquiline (six months), terizidone, clofazimine, linezolid, and levofloxacin. We assumed 12% of all individuals receiving tuberculosis preventive treatment received rifamycin-based regimens and 65% of all individuals treated for rifampicin-resistant tuberculosis received short regimens in 2024. The value associated with short rifampicin-resistant regimens remained the same while with rifamycin-based it increased to 50% by 2030 in status quo scenarios; for both, use increased to 100% by 2030 with The Intervention Package.

We worked with the national tuberculosis programme and others with experience screening high-risk communities—in this case, those living in the 22 highest TB prevalence subdistricts. Based on a population size of 1,109,090 and a target to screen on 60% of this population, we estimated 4 teams composed of one nurse, one physician, one driver, and one social worker each would be deployed per subdistrict (22 subdistricts) to conduct this screening at a rate of 60 people screened per day. The screening would take 126 days to complete. For each team, we assumed they would have a mobile van fully equipped with chest x-ray, with an annual, annuitized cost of $55,100 USD, they would require $1575 USD in fuel costs, $290 in coolers and ice to transport sputum samples, 63 days in hotel (at a cost of $70 per night), and a per diem of $27 per person per day. Complete costs are described below.

| Cost Parameter | Description | Mean (USD 2023) |
|---|---|---|
| Cost to perform symptom screening per person | From costing of South Africa's strategic plan. 11.58 ZAR in 2022. Conversion of 0.0613 and 3.8% inflation based on deflator | 0.74 |
| Cost of household contact investigation per index patient | Based on door-to-door screening costs. 2.36 HHC per index patient. One person to screen averages to 56.78 ZAR in 2022 based on strategic plan costing. Multiply this cost by number of contacts and convert to USD and inflate to 2023 using deflator. | 8.53 |
| Cost of TST, inclusive of materials and personnel time | From ingredients list in South Africa Strategic plan, with cost of tuberculin for one use of $0.93 USD. Visits are 49.04 ZAR in 2022, converted at 0.0613 and inflated 3.8% | 4.05 |
| Cost of IGRA, inclusive of materials | From ingredients list in South Africa Strategic plan, and converted to 2023 from 2022 (communication from Gesine) | 78.5 |
| Cost of antigen-based skin test, inclusive of materials and personnel time | Assumed $1.60 USD per one use, all other costs same as tuberculin | 4.72 |
| Cost of Xpert per person | Cost of 218.84 ZAR per test in strategic plan in 2022. Conversion of 0.0613 and 3.8% inflation based on deflator, minus $2 due to price reduction | 17.93 |
| Cost of HIV test per person | Cost of 8.14 ZAR per test in strategic plan in 2022. Conversion of 0.0613 and 3.8% inflation based on deflator | 0.52 |
| Cost of CRP per person | Cost of 102.74 ZAR per test in strategic plan in 2022. Conversion of 0.0613 and 3.8% inflation based on deflator | 6.54 |
| Cost of CXR per person | Cost of 294.14 ZAR per test in strategic plan in 2022. Conversion of 0.0613 and 3.8% inflation based on deflator | 18.72 |
| Annual cost of CAD license (annuitized); considered in analysis as a new investment and part of implementation in 5% of all healthcare facilities | InferRead CAD software (one license), with annual costs and annuitized capital costs at 3% over 10 years for software and installation/training, 5 years for laptop, and 3 years for initial maintenance agreement with the next years costing $250 per annum (GDF price catalogue) | 1058 |
| Cost of AFB smear, ZN | Cost of 36.95 ZAR per test in strategic plan in 2022. Conversion of 0.0613 and 3.8% inflation based on deflator | 2.35 |
| Cost of sputum collection per person | South Africa's strategic plan: 57.14 in 2022. Conversion of 0.0613 and 3.8% inflation based on deflator | 3.64 |
| Cost of liquid culture per sample | Cost of 75.45 based on NHLS pricing reported in Cape Town in 2018 (https://resource.capetown.gov.za/documentcentre/Documents/Agreements%20and%20contracts/SCMB%2053-10-18.pdf). Conversion of 0.076 and deflator of 28.1% | 7.35 |
| Cost of solid culture per sample | Assumed proportional increase similar to what is seen in Kenya (31.9% increase) | 9.69 |
| Cost of first-line DST per panel | Cost of 79.28 per drug based on NHLS pricing reported in Cape Town in 2018 (https://resource.capetown.gov.za/documentcentre/Documents/Agreements%20and%20contracts/SCMB%2053-10-18.pdf). Conversion of 0.076 and deflator of 28.1%. Assume 4 drugs | 30.87 |
| Cost of second-line DST per panel | Cost of 79.28 per drug based on NHLS pricing reported in Cape Town in 2018 (https://resource.capetown.gov.za/documentcentre/Documents/Agreements%20and%20contracts/SCMB%2053-10-18.pdf). Conversion of 0.076 and deflator of 28.1%. Assume 3 drugs | 23.16 |
| Cost of first-line LPA per sample | Cost of 229.35 in strategic plan. No distinguishing between first and second line, so assume identical. Conversion of 0.0613 and 3.8% inflation based on deflator | 14.59 |
| Cost of second-line LPA per sample | Cost of 229.35 in strategic plan. No distinguishing between first and second line, so assume identical. Conversion of 0.0613 and 3.8% inflation based on deflator | 14.59 |
| Cost of pre-TPT evaluation, considering a treatment visit | From costing of South Africa's strategic plan. 40.48 ZAR in 2022. Conversion of 0.0613 and 3.8% inflation based on deflator | 2.58 |
| Cost of 6INH if complete, adults | From Strategic plan, cost of one follow-up visit is 17.37 ZAR in 2022 ($1.11 USD in 2023). Follow-up assumed monthly. Cost of six months of isoniazid in strategic plan is 104.43 ZAR in 2022 (6.65 in 2023 USD) | 13.31 |
| Cost of 6INH if complete, children | From Strategic plan, cost of one follow-up visit is 17.37 ZAR in 2022 ($1.11 USD in 2023). Follow-up assumed monthly. Cost of six months of isoniazid in strategic plan is 253.8 ZAR in 2022 (16.15 in 2023 USD) | 22.81 |
| Cost of 3HR if complete, adults | From Strategic plan, cost of one follow-up visit is 17.37 ZAR in 2022 ($1.11 USD in 2023). Follow-up assumed monthly. Cost of six months of isoniazid in strategic | 20.17 |

| Cost Parameter | Description | Mean (USD 2023) |
|---|---|---|
| | plan is 264.6 ZAR in 2022 (16.84 in 2023 USD) | |
| Cost of 3HR if complete, children | From Strategic plan, cost of one follow-up visit is 17.37 ZAR in 2022 ($1.11 USD in 2023). Follow-up assumed monthly. Cost of six months of isoniazid in strategic plan is 666 ZAR in 2022 (42.38 in 2023 USD) | 45.71 |
| Cost of 3HP if complete, adults | From Strategic plan, cost of one follow-up visit is 17.37 ZAR in 2022 ($1.11 USD in 2023). Only 1 follow-up assumed as all doses given at start of treatment and SAT. Strategic plan costs are 106.25 ZAR per month in 2022. Equivalent to 20.28 USD in 2023 | 21.39 |
| Cost of 3HP if complete, children | From Strategic plan, cost of one follow-up visit is 17.37 ZAR in 2022 ($1.11 USD in 2023). Only 1 follow-up assumed as all doses given at start of treatment and SAT. Strategic plan costs are 106.25 ZAR per month in 2022. Equivalent to 20.28 USD in 2023 | 21.39 |
| Cost of 1HP if complete, adults | From Strategic plan, cost of one follow-up visit is 17.37 ZAR in 2022 ($1.11 USD in 2023). Follow-up assumed monthly. Costs are from IHT TB as rifapentine alone not listed in strategic plan. Cost is 20.72 USD in 2021, equivalent to 22.61 USD in 2023 (9.1% deflator) | 23.72 |
| Cost of 1HP if complete, children | From Strategic plan, cost of one follow-up visit is 17.37 ZAR in 2022 ($1.11 USD in 2023). Follow-up assumed monthly. Cost is 20.72 USD in 2021, equivalent to 22.61 USD in 2023 (9.1% deflator) | 23.72 |
| Cost of 6Lfx if complete, adults | From Strategic plan, cost of one follow-up visit is 17.37 ZAR in 2022 ($1.11 USD in 2023). Follow-up assumed monthly. Levofloxacin not explicitly in strategic plan, used IHT TB tool. $18 USD in 2021, with 9.1% deflator it is 19.64 USD in 2023 | 26.3 |
| Cost of 6Lfx if complete, children | From Strategic plan, cost of one follow-up visit is 17.37 ZAR in 2022 ($1.11 USD in 2023). Follow-up assumed monthly. Levofloxacin not explicitly in strategic plan, used IHT TB tool. $16.20 USD in 2021, with 9.1% deflator it is 17.67 USD in 2023 | 24.33 |
| Cost of 3HR if incomplete, adults; assumes 50% the price of complete | Assumed half the cost of complete treatment | 10.09 |
| Cost of 3HR if incomplete, children; assumes 50% the price of complete | Assumed half the cost of complete treatment | 22.86 |
| Cost of 6INH if incomplete, adults; assumes 50% the price of complete | Assumed half the cost of complete treatment | 6.66 |
| Cost of 6INH if incomplete, children; assumes 50% the price of complete | Assumed half the cost of complete treatment | 11.41 |
| Cost of 3HP if incomplete, adults; assumes 50% the price of complete | Assumed half the cost of complete treatment | 10.70 |
| Cost of 3HP if incomplete, children; assumes 50% the price of complete | Assumed half the cost of complete treatment | 10.70 |
| Cost of 1HP if incomplete, adults; assumes 50% the price of complete | Assumed half the cost of complete treatment | 11.86 |
| Cost of 1HP if incomplete, children; assumes 50% the price of complete | Assumed half the cost of complete treatment | 11.86 |
| Cost of 6Lfx if incomplete, adults; assumes 50% the price of complete | Assumed half the cost of complete treatment | 13.15 |
| Cost of 6Lfx if incomplete, children; assumes 50% the price of complete | Assumed half the cost of complete treatment | 12.17 |
| Cost of drug-susceptible TB treatment, adults | Strategic plan costing is 2098.92 ZAR in 2022. Equivalent to 133.55 USD in 2023 with 3.8% deflator and conversion of 0.0613 | 133.55 |
| Cost of 4-month DS-TB treatment, children | Microcosting using strategic plan assuming only 4 months of treatment and 2/3 the staff time efforts. Equivalent to 1054.54 ZAR in 2022. Equivalent to 67.10 USD in 2023 with 3.8% deflator and conversion of 0.0613 | 67.1 |
| Cost of 6-month DS-TB treatment, adults | Strategic plan costing is 1545.73 ZAR in 2022. Equivalent to 98.35 USD in 2023 with 3.8% deflator and conversion of 0.0613 | 98.35 |
| Cost of BPaLL RR-TB treatment, adults<br>(6 months bedaquiline, pretomanid, linezolid, levofloxacin) | Estimated from Sweeney et al, with replacement of moxifloxacin with levofloxacin, the preference in South Africa. Cost of 842.21 in 2019 USD; $335.80 were non-drug expenses. Deflator at 22.4% | 1030.87 |
| Cost of BPaLL RR-TB treatment, children<br>(6 months bedaquiline, pretomanid, linezolid, levofloxacin) | Estimated from Sweeney et al, with replacement of moxifloxacin with levofloxacin, the preference in South Africa. Cost of 824.46 in 2019 USD; $335.80 were non-drug expenses. Deflator at 22.4% | 1009.14 |
| Cost of 18-month all oral RR-TB treatment, adults<br>(6 months bedaquiline, 18 months terizidone, clofazimine, linezolid, levofloxacin) | Drug costs not provided in sufficient detail in strategic plan. IHT TB tool used: Cost of drugs in 2021 = 1908.54 (deflator 9.1%). Non-drug costs from Sweeney et al, of 374.20 in 2019 USD (Deflator 22.4%) = 458.02 | 2539.65 |
| Cost of 18-month all oral RR-TB treatment, children<br>(6 months bedaquiline, 18 months terizidone, clofazimine, linezolid, levofloxacin) | Drug costs not provided in sufficient detail in strategic plan. IHT TB tool used: Cost of drugs in 2021 = 2601.54 (deflator 9.1%). Non-drug costs from Sweeney et al, of 374.20 in 2019 USD (Deflator 22.4%) = 458.02 | 3296.3 |
| Patient costs per month of DS-TB treatment; mean episode costs were divided by average duration of treatment (6 months) | Cost of 1494 ZAR per month, assuming 6 months in 2021 patient cost survey. Converted at a rate of 0.0677 and deflator of 9.1% | 110.35 |

| Cost Parameter | Description | Mean (USD 2023) |
|---|---|---|
| Patient costs per month of RR-TB treatment; mean episode costs were divided by average duration of treatment (18 months) | Calculated as less than DS-TB, so assumed the same. | 110.35 |
| Cost of post-tuberculosis healthcare utilization; considers 12.2 outpatient visits and 2.2 additional inpatient bed days | From Romanowski et al, considers 12.2 outpatient visits (health centre with beds) and 2.2 additional inpatient bed days (secondary hospital). Costs come from WHO-CHOICE in USD and inflated from 2008 using GDP deflator (125% increase) | 547.24 |
| Patient costs per month of TPT; assumed as a proportion of patient costs of DS-TB treatment (9.1%) | Assumed to be 9.09% of the DS-TB patient cost based on Shepardson et al. | 10.03 |
| Annual cost of healthcare worker training in intervention scenarios; annual recurrent 1-day training cost in intervention scenarios for training of 10% of healthcare workforce, plus TST training | Inclusive of annual costs for training 141 nurses on TST for 1.5 days per year, training 4700 physicians and 29,600 nurses on TB 1 day per year, and paying a full time nurse manager to oversee implementation per million population; plus $10,000 cost of a portion of a zoom license to carry out virtual training | 5,800,000 |
| Annual cost of active case finding training in years conducted for The Intervention Package; assumes 2 days training for all years of case finding in high-risk communities, applied as an annual implementation cost | Inclusive of training teams conducting case finding for 2 days - 88 physicians, 88 nurses, 88 social workers, 88 drivers | 87,000 |
| Annual cost of active case finding training in years conducted, enhanced scenario; assumes 2 days training for all years of case finding in high-risk communities, applied as an annual implementation cost | Inclusive of training teams conducting case finding for 2 days - 88 physicians, 176 nurses, 88 social workers, 88 drivers | 104,000 |
| Cost to screen one person during active case finding for The Intervention Package; considers costs of materials (cooler, ice pack, fuel, van), 50% of nights stayed out of town (hotel, food), personnel costs | 126 days of screening per year, with 110 teams. Each team has a van (annuitized at 10y lifespan) equipped with CXR, coolers, ice packs, fuel (prorated at $10,000 per 240 days), per diem for hotel and food for half the screening days. Cost includes the personnel trained above (prorated on a per team basis). Overall cost is a per person screened. | 19.4 |
| Cost to screen one person during active case finding, enhanced scenario; considers costs of materials (cooler, ice pack, fuel, van), 50% of nights stayed out of town (hotel, food), personnel costs, and cost of antigen-based skin test | 189 days of screening per year, with 110 teams. Each team has a van (annuitized at 10y lifespan) equipped with CXR, coolers, ice packs, fuel (prorated at $10,000 per 240 days), per diem for hotel and food for half the screening days. Costs also include the materials for TB antigen skin testing. Cost includes the personnel trained above (prorated on a per team basis). Overall cost is a per person screened. | 23.04 |

Abbreviations: 95% UR, 95% uncertainty range; TST, tuberculin skin test; IGRA, interferon-gamma release assay; HIV, human immunodeficiency virus; CRP, C-reactive protein; CXR, chest x-ray; CAD, computer aided detection; AFB, acid-fast bacilli; ZN, Ziehl-Nielsen; DST, drug susceptibility test; LPA, line probe assay; TPT, tuberculosis preventive treatment; 6INH, six-months of isoniazid daily; 3HR, three-months isoniazid and rifampicin daily; 3HP, three-months isoniazid and rifapentine weekly; 1HP, one-month isoniazid and rifapentine daily; 6Lfx, six months levofloxacin daily; DS-TB, drug-susceptible tuberculosis; RR-TB, rifampicin-resistant tuberculosis; BPaLM, bedaquiline, pretomanid, linezolid, and moxifloxacin; BPaLL, bedaquiline, pretomanid, linezolid, and levofloxacin.

**References this section**

1. IHT. Integrated Health Tool: TB. 2023. https://tb.integratedhealthtool.org/ (accessed March 12, 2024).
2. Romanowski K, Law MR, Karim ME, et al. Healthcare Utilization After Respiratory Tuberculosis: A Controlled Interrupted Time Series Analysis. Clin Infect Dis 2023; 77: 883–91.
3. Shepardson D, Marks SM, Chesson H, et al. Cost-effectiveness of a 12-dose regimen for treating latent tuberculous infection in the United States. Int J Tuberc Lung Dis 2013; 17: 1531–7.
4. WHO-CHOICE. WHO. https://www.who.int/choice/cost-effectiveness/inputs/en/ (accessed Feb 11, 2019).
5. Stop TB Partnership | Global Drug Facility - Products List. https://www.stoptb.org/facilitate-access-to-tb-drugs-diagnostics/global-drug-facility-gdf (accessed March 11, 2024).
6. World Health Organization. WHO consolidated guidelines on tuberculosis: module 3: diagnosis: tests for TB infection. Geneva, Switzerland: WHO, 2022.
7. Sweeney S, Berry C, Kazounis E, et al. Cost-effectiveness of short, oral treatment regimens for rifampicin resistant tuberculosis. PLOS Global Public Health 2022; 2: e0001337.
8. National Health Laboratory Service Cost Catalogue, 2018. South Africa. https://resource.capetown.gov.za/documentcentre/Documents/Agreements%20and%20contracts/SCMB%2053-10-18.pdf (Accessed March 11,2024)

#### Reflexive Testing Algorithms and Number to Test to Detect One Person with Tuberculosis Outside Priority Populations

Each country employed different levels of reflexive testing algorithms (i.e., subsequent tests performed) for individuals diagnosed with drug-susceptible or rifampicin-resistant tuberculosis. We included these costs in The Intervention Package of all countries. They are described in the table below. Further, to estimate costs associated with screening individuals for tuberculosis who did not belong to any of our priority populations we had to make certain assumptions. First, we assumed those outside of our priority populations would only be diagnosed with tuberculosis if they had symptoms. Second, we took a recent review of tuberculosis prevalence surveys (Frascella B et al. *Clin Infect Dis*. 2021;73(3):e830-41) to estimate the approximate prevalence of tuberculosis among those with symptoms. We used the data from these surveys to fit a simple linear regression model of prevalence to national tuberculosis incidence ($R^2 = 0.72$), which suggested the prevalence of tuberculosis among those with symptoms increased by 0.000031 per 1 unit increase in tuberculosis incidence per 100,000. We calculated the number of people each year who initiated tuberculosis treatment but did not belong to a priority population and used the prevalence among this population, plus the diagnostic performance of country-level algorithms in this population, to estimate how many individuals who would have needed to have been screened to identify one person with tuberculosis.

| Country | DS-TB Reflexive Testing | RR-TB Reflexive Testing |
|---|---|---|
| Brazil | Only in high-risk populations: liquid culture | Phenotypic DST first-line and second-line |
| Georgia | AFB sputum smear | AFB sputum smear, first-line LPA, second-line LPA |
| Kenya | None | Liquid culture, Phenotypic DST first-line and second-line, first-line LPA, second-line LPA |
| South Africa | First-line LPA | AFB sputum smear, Liquid culture, Phenotypic DST first-line and second-line, first-line LPA, second-line LPA |

Abbreviations: DS-TB, drug-susceptible tuberculosis; RR-TB, rifampicin-resistant tuberculosis; DST, drug susceptibility testing; AFB, acid fast bacilli; LPA, line probe assay

#### Additional costing methods and explanations

For people receiving TPT, we used costs prorated based on the proportion expected to complete the regimen, assuming those who did not complete TPT received 50% of doses. Completion rates for each regimen are found in the Model Parameters Table and country-specific costs for TPT regimens are found in the costing sections of this Appendix.

We used a study in Canada estimating the excess healthcare utilization seen among tuberculosis survivors (i.e., utilization above what would be expected had they not developed tuberculosis based on healthcare utilization in the years prior to diagnosis). In this study, there were an additional 12.2 outpatient visits and 2.2 additional inpatient days. Cost estimates for each country and how we derived them are located in the costing sections of this Appendix.

To support implementation in all countries, we assumed 5% of all health facilities would have computer aided detection (CAD) for tuberculosis on chest x-ray installed, one nurse manager per million population would be hired to support scale-up, 10% of physicians and nurses would receive one-day of virtual, annual training on tuberculosis, one nurse per 4000 annual household contacts would receive annual 1.5-day training on tuberculin skin test (TST) administration and reading, and for the three years while case-finding activities were being conducted in the country-specific high-risk populations/communities, there would be training and implementation costs. See country-specific cost descriptions for additional details.

Patient cost surveys were available for all countries except Georgia, where we used a recent meta-regression to estimate patient costs associated with tuberculosis treatment. For estimates of patient costs for drug-susceptible tuberculosis, we divided the entire episode patient cost by 6 to estimate the per-month cost, while for rifampicin-resistant tuberculosis, we divided the entire episode patient cost by 18 to estimate the per-month cost as this is the most common duration of treatment. To estimate patient costs associated with tuberculosis preventive treatment, we used a study in the USA where patient costs associated with three months of once-weekly isoniazid and rifapentine, nine months of daily isoniazid, and drug-susceptible tuberculosis were compared. The average per month patient cost associated with the TPT regimens was $28.74 and for drug-susceptible tuberculosis, it was $317.78. This is equivalent to the average per month patient cost of TPT being 9% that of tuberculosis disease treatment.

#### Definition of the Enhanced Package

| Population | Percent of Population Reached by Algorithm | Age Group (years) | Tuberculosis Disease Algorithm | Tuberculosis Infection Algorithm (Assumes Tuberculosis Disease is Already Ruled Out)* |
|---|---|---|---|---|
| People living with HIV not on ART | 100% | 0-4 | Systematic tuberculosis symptom screening; Xpert Ultra if symptoms present. | No testing for tuberculosis infection; 3HR prescribed for those <2 years and 3HP for those 2-4 years. |
| | | 5-9 | Systematic assessment with both tuberculosis symptom screening and digital chest x-ray with CAD; Xpert Ultra if symptoms present or abnormal chest x-ray. | No testing for tuberculosis infection; 3HP prescribed. |
| | | 10-14 | Systematic tuberculosis symptom screening; CRP performed among those with symptoms. Xpert Ultra if symptoms present and CRP positive. | No testing for tuberculosis infection; 3HP prescribed. |
| | | ≥15 | Systematic tuberculosis symptom screening; CRP performed among those with symptoms. Xpert Ultra if symptoms present and CRP positive. | No testing for tuberculosis infection; 1HP prescribed. |
| People living with HIV on ART | 100% | 0-4 | Annual screening for tuberculosis using symptom screening; Xpert Ultra if symptoms present. | No testing for tuberculosis infection; 3HR prescribed for those <2 years and 3HP for those 2-4 years. |
| | | 5-9 | Annual screening for tuberculosis using symptom screening; Xpert Ultra if symptoms present. | No testing for tuberculosis infection; 3HP prescribed. |
| | | 10-14 | Annual screening for tuberculosis using both symptom screening and digital chest x-ray with CAD; Xpert Ultra if symptoms present or abnormal chest x-ray | No testing for tuberculosis infection; 3HP prescribed. |
| | | ≥15 | Annual screening for tuberculosis using both symptom screening and digital chest x-ray with CAD; Xpert Ultra if symptoms present or abnormal chest x-ray | No testing for tuberculosis infection; 1HP prescribed. |
| Household Contacts | 100% | 0-4 | Systematic symptom screening and chest x-ray; Xpert Ultra if symptoms present or abnormal chest x-ray | Testing with antigen-based skin test; if positive, 3HR prescribed for those <2 years and 3HP for those 2-4 years. |
| | | 5-14 | Systematic symptom screening and chest x-ray with CAD; Xpert Ultra if symptoms present or abnormal chest x-ray | Testing with antigen-based skin test; if positive, 3HP prescribed |
| | | ≥15 | Systematic symptom screening and chest x-ray with CAD; Xpert Ultra if symptoms present or abnormal chest x-ray | Testing with antigen-based skin test; if positive, 1HP prescribed |
| High-Risk Population | 0% | 0-14 | No specific intervention. | No specific intervention. |
| | 90% | ≥15 | Systematic symptom screening and chest x-ray with CAD. Xpert Ultra if symptoms present or abnormal chest x-ray. The intervention is implemented in this population for 3 consecutive years (2024-2026) then halted. | All people reached for tuberculosis screening will also receive an antigen-based skin test; if positive and tuberculosis has been ruled out, then 1HP prescribed. The intervention is implemented in this population for 3 consecutive years (2024-2026) then halted. |

*TPT is only provided one time in the model.
Note: all costs are provided in the costing section of this Appendix.
Abbreviations: HIV, human immunodeficiency virus; CRP, C-reactive protein; ART, antiretroviral treatment; CAD, computer aided detection; TPT, tuberculosis preventive treatment; 3HR, three-months isoniazid and rifampicin daily; 3HP, three-months isoniazid and rifapentine weekly; 1HP, one-month isoniazid and rifapentine daily.

### Additional Results

#### Incremental Outcomes (Absolute Values), by 2050.

| Country and Comparison | Number Developing TB | Number Dying from TB | Years of Life Lost due to TB | TB-Associated DALYs | Number Screened for TB | Number Initiating TPT | TB-Related Health System Cost | TB-Associated Patient and Family Costs | TB-Related Societal Cost |
|---|---|---|---|---|---|---|---|---|---|
| Brazil | | | | | | | | | |
| The Intervention Package without TPT Compared to Status Quo | -254,971 (-287,564 to -234,154) | -31,702 (-35,699 to -29,152) | -1,165,677 (-1,325,078 to -1,064,709) | -1,209,001 (-1,375,859 to -1,103,319) | 41,430,986 (38,737,106 to 45,569,807) | -30,442 (-38,272 to -25,982) | 1.3 billion (0.06 billion to 2.26 billion) | -50 million (-113 million to -15 million) | -9.14 billion (-11.2 billion to -7.65 billion) |
| The Intervention Package with TPT Compared to The Intervention Package without TPT | -229,540 (-252,929 to -213,971) | -20,567 (-22,602 to -19,182) | -773,086 (-856,161 to -717,643) | -814,593 (-902,858 to -755,844) | -4,679,987 (-5,562,827 to -4,121,233) | 14,406,367 (13,551,628 to 15,721,855) | 0.26 billion (0.05 billion to 0.52 billion) | 430 million (-90 million to 1412 million) | -6.21 billion (-7.33 billion to -5.08 billion) |
| Georgia | | | | | | | | | |
| The Intervention Package without TPT Compared to Status Quo | -6,257 (-11,640 to -3,560) | -772 (-1,269 to -496) | -29,180 (-48,822 to -18,580) | -33,210 (-55,279 to -21,131) | 2,650,774 (2,209,823 to 3,322,408) | -1,326 (-1,760 to -1,012) | 37 million (4 million to 70 million) | -7 million (-20 million to -1 million) | -0.16 billion (-0.3 billion to -0.08 billion) |
| The Intervention Package with TPT Compared to The Intervention Package without TPT | -3,960 (-5,833 to -2,817) | -198 (-270 to -153) | -7,409 (-10,390 to -5,574) | -8,790 (-12,453 to -6,546) | -664,978 (-994,073 to -470,466) | 767,492 (639,060 to 953,900) | 22 million (9 million to 40 million) | 31 million (-2 million to 94 million) | 0 billion (-0.04 billion to 0.07 billion) |
| Kenya | | | | | | | | | |
| The Intervention Package without TPT Compared to Status Quo | -949,389 (-1,305,679 to -843,836) | -205,138 (-267,080 to -183,061) | -8,088,043 (-10,569,104 to -7,195,363) | -8,530,784 (-11,113,181 to -7,600,633) | 111,302,193 (95,913,969 to 139,289,951) | -5,368,074 (-6,678,016 to -4,593,241) | 1.3 billion (-0.69 billion to 4.86 billion) | -68 million (-160 million to -20 million) | -15.75 billion (-21.18 billion to -11.84 billion) |
| The Intervention Package with TPT Compared to The Intervention Package without TPT | -416,229 (-488,785 to -372,738) | -30,233 (-34,971 to -27,657) | -1,260,062 (-1,476,157 to -1,151,223) | -1,389,712 (-1,623,051 to -1,269,646) | -72,928,397 (-98,018,033 to -61,582,099) | 26,371,419 (23,854,528 to 29,019,505) | -0.58 billion (-1.91 billion to 0.17 billion) | 150 million (-81 million to 513 million) | -3.08 billion (-4.56 billion to -2.17 billion) |
| South Africa | | | | | | | | | |
| The Intervention Package without TPT Compared to Status Quo | -991,591 (-1,584,431 to -550,257) | -248,602 (-400,609 to -103,835) | -9,007,443 (-14,131,410 to -3,810,368) | -9,530,038 (-14,849,978 to -4,102,683) | 169,165,222 (95,210,590 to 240,147,807) | -623,839 (-1,550,744 to -62,623) | 2.37 billion (-2.34 billion to 6.52 billion) | -158 million (-398 million to -30 million) | -58.83 billion (-93.74 billion to -21.82 billion) |
| The Intervention Package with TPT Compared to The Intervention Package without TPT | -436,806 (-539,529 to -262,913) | -27,416 (-35,379 to -17,280) | -1,204,573 (-1,515,401 to -711,698) | -1,363,026 (-1,690,753 to -814,873) | -98,640,091 (-157,751,972 to -38,906,890) | 37,403,007 (27,760,013 to 45,023,330) | -0.6 billion (-2.15 billion to 0.33 billion) | 228 million (-210 million to 859 million) | -8.54 billion (-11.58 billion to -4.45 billion) |

Notes: Value represent mean (95% UR). All costs are in 2023 USD. No discounting.
Abbreviations: TPT, tuberculosis preventive treatment; TB, tuberculosis; DALY, disability-adjusted life years

#### Incremental Outcomes (Relative Values), by 2050.

| Country and Comparison | Number Developing TB | Number Dying from TB | Years of Life Lost due to TB | TB-Associated DALYs | Number Screened for TB | Number Initiating TPT | TB-Related Health System Cost | TB-Associated Patient and Family Costs | TB-Related Societal Cost |
|---|---|---|---|---|---|---|---|---|---|
| Brazil | | | | | | | | | |
| The Intervention Package without TPT Compared to Status Quo | -9.6% (-10% to -9.3%) | -14.1% (-14.6% to -13.8%) | -13.4% (-14% to -13.1%) | -10.8% (-11.3% to -10.4%) | 2217.6% (2132.4% to 2302.3%) | -3.6% (-4.3% to -3.2%) | 67.5% (1.9% to 148.1%) | -4% (-14.1% to -1.1%) | -11.2% (-12.8% to -9.9%) |
| The Intervention Package with TPT Compared to The Intervention Package without TPT | -9.5% (-9.8% to -9.3%) | -10.7% (-10.8% to -10.5%) | -10.3% (-10.5% to -10.2%) | -8.1% (-8.4% to -8%) | -10.8% (-11.7% to -10.2%) | 1781.4% (1714.5% to 1848.5%) | 7.8% (1.1% to 17.1%) | 44.6% (-3.2% to 202.7%) | -8.6% (-9.5% to -7.3%) |
| Georgia | | | | | | | | | |
| The Intervention Package without TPT Compared to Status Quo | -14.4% (-19.6% to -11%) | -25.8% (-31.7% to -21.6%) | -26.4% (-32.4% to -22.2%) | -17.8% (-22.9% to -14.3%) | 18105.2% (13915.3% to 25251.7%) | -23.7% (-31.5% to -19.4%) | 85.8% (5.6% to 186.5%) | -11.5% (-26% to -4%) | -19.1% (-26.4% to -12.7%) |
| The Intervention Package with TPT Compared to The Intervention Package without TPT | -10.9% (-12.3% to -9.8%) | -9% (-9.8% to -8.4%) | -9.2% (-10.2% to -8.5%) | -5.8% (-6.7% to -5.2%) | -24.7% (-29.8% to -21.1%) | 18061.9% (14632.9% to 24569.9%) | 26.5% (9% to 51.4%) | 83.3% (-3.2% to 346.4%) | 0.6% (-5% to 10.5%) |
| Kenya | | | | | | | | | |
| The Intervention Package without TPT Compared to Status Quo | -30.3% (-33.1% to -29%) | -45.1% (-46.8% to -43%) | -45.4% (-46.6% to -43.5%) | -41.7% (-43.2% to -39.8%) | 878.4% (579.1% to 1156.1%) | -46.1% (-47.2% to -44.5%) | 49.6% (-14% to 172.6%) | -17.7% (-33.6% to -10.7%) | -38.4% (-42.5% to -30.1%) |
| The Intervention Package with TPT Compared to The Intervention Package without TPT | -19.2% (-20.1% to -17.6%) | -12.2% (-12.9% to -11.1%) | -13% (-13.9% to -11.8%) | -11.7% (-12.4% to -10.7%) | -58.5% (-63.6% to -55.2%) | 425.2% (299.9% to 509.6%) | -11.1% (-23% to 6.3%) | 77.3% (-12.5% to 356%) | -12.1% (-15.2% to -9.4%) |
| South Africa | | | | | | | | | |
| The Intervention Package without TPT Compared to Status Quo | -22.7% (-27.2% to -19.4%) | -35.5% (-38.1% to -30%) | -36.8% (-39.7% to -29.8%) | -33.5% (-36.6% to -26.5%) | 3038% (1320.5% to 6025.8%) | -12% (-22.1% to -1.8%) | 154.9% (-31.5% to 534.4%) | -11.7% (-22% to -4.5%) | -34.3% (-38.5% to -25%) |
| The Intervention Package with TPT Compared to The Intervention Package without TPT | -13.1% (-14.4% to -11.2%) | -6.4% (-7.9% to -4.8%) | -8.2% (-9.5% to -6.5%) | -7.5% (-8.5% to -6.3%) | -54.4% (-64.7% to -38.4%) | 929.8% (551.9% to 1342.9%) | -9% (-24.8% to 11.2%) | 29.8% (-8.2% to 133.8%) | -7.9% (-9.5% to -6.3%) |

Notes: Value represent mean (95% UR). All costs are in 2023 USD. No discounting.

Abbreviations: TPT, tuberculosis preventive treatment; TB, tuberculosis; DALY, disability-adjusted life years

#### Relative Change in Tuberculosis Compared to the Status Quo

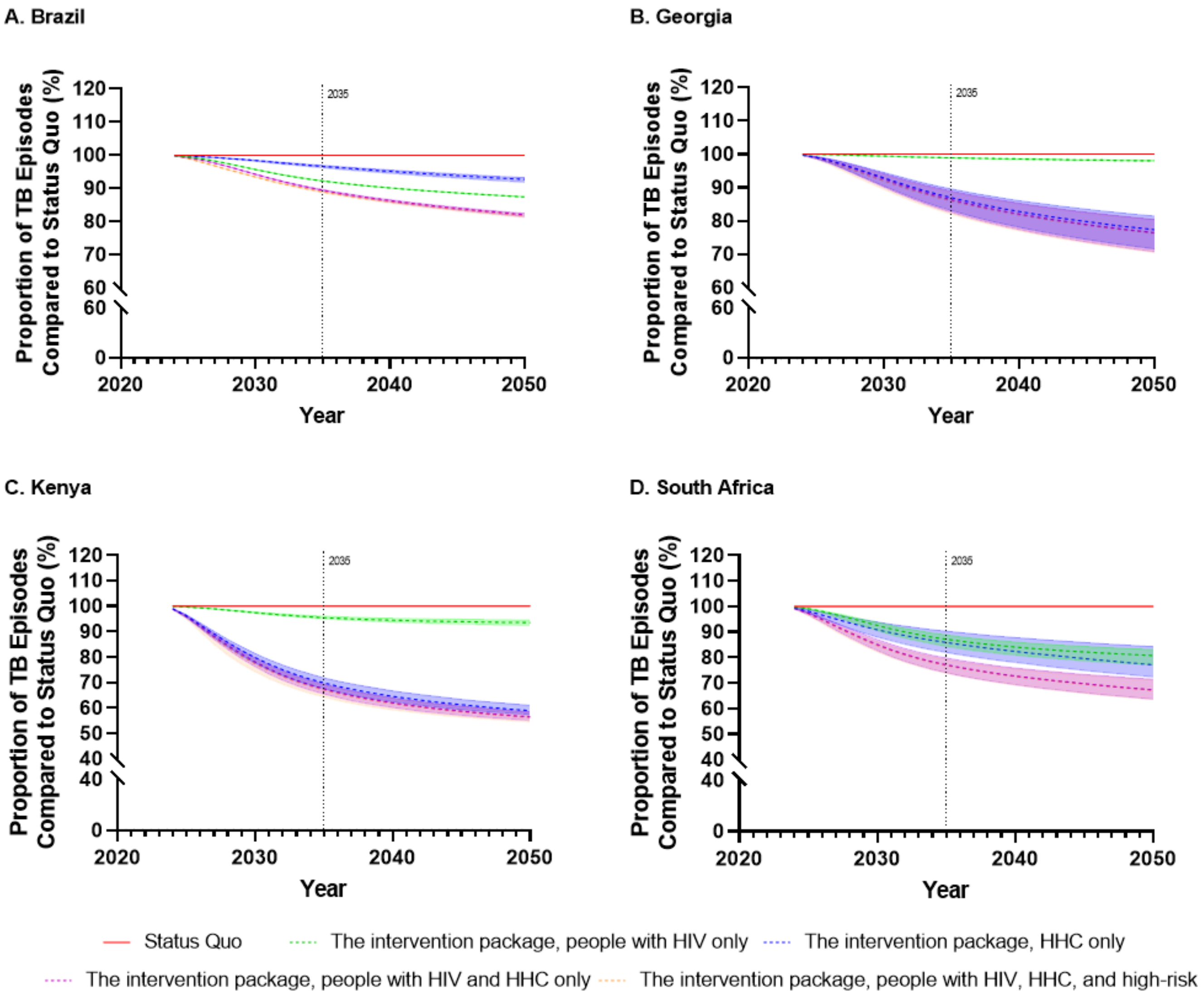

**Caption**: Trajectories from 2024 through to 2050 of the cumulative change in tuberculosis episodes across the entire population of each country. Strategies shown are the status quo and the intervention package scaled up with TPT in different priority populations. Different scales have been used for each graph. Shaded regions represent the 95% uncertainty range.

#### Outcomes (Relative) Among People living with HIV by 2050

| Country and Comparison | Number Developing TB | Number Dying from TB | Years of Life Lost due to TB | TB-Associated DALYs |
|---|---|---|---|---|
| Brazil | | | | |
| The Intervention Package without TPT Compared to Status Quo | -21.4% (-21.6% to -21.2%) | -22.5% (-22.8% to -22.3%) | -22.7% (-23% to -22.5%) | -22% (-22.4% to -21.9%) |
| The Intervention Package with TPT Compared to The Intervention Package without TPT | -24.9% (-25.1% to -24.7%) | -19% (-19.2% to -18.9%) | -19% (-19.2% to -18.9%) | -18.7% (-18.9% to -18.5%) |
| Georgia | | | | |
| The Intervention Package without TPT Compared to Status Quo | -7.3% (-9.9% to -5.6%) | -15.5% (-17.9% to -14.1%) | -15.7% (-18.2% to -14.3%) | -15% (-17.4% to -13.6%) |
| The Intervention Package with TPT Compared to The Intervention Package without TPT | -20.2% (-20.9% to -19.7%) | -12% (-12.5% to -11.7%) | -12.2% (-12.6% to -11.8%) | -12.1% (-12.6% to -11.8%) |
| Kenya | | | | |
| The Intervention Package without TPT Compared to Status Quo | -22.6% (-24.6% to -21.4%) | -29.3% (-31% to -28.2%) | -29.8% (-31.4% to -28.6%) | -28.5% (-30.2% to -27.3%) |
| The Intervention Package with TPT Compared to The Intervention Package without TPT | -17% (-17.8% to -15.8%) | -10.8% (-11.5% to -10.1%) | -11.6% (-12.2% to -10.8%) | -11.2% (-11.8% to -10.4%) |
| South Africa | | | | |
| The Intervention Package without TPT Compared to Status Quo | -17.6% (-21.2% to -14.8%) | -23.9% (-26.6% to -21.7%) | -24% (-26.6% to -21.7%) | -21.5% (-25.2% to -18.2%) |
| The Intervention Package with TPT Compared to The Intervention Package without TPT | -7.8% (-11.4% to -4.8%) | -3% (-6% to -0.8%) | -3.5% (-6.8% to -1.2%) | -3.5% (-6.4% to -1.4%) |

Notes: Value represent mean (95% UR). All costs are in 2023 USD. No discounting.
Abbreviations: TPT, tuberculosis preventive treatment; TB, tuberculosis; DALY, disability-adjusted life years

#### Annual Change in TB Disease Incidence Among People Living with HIV with The Intervention Package with TPT

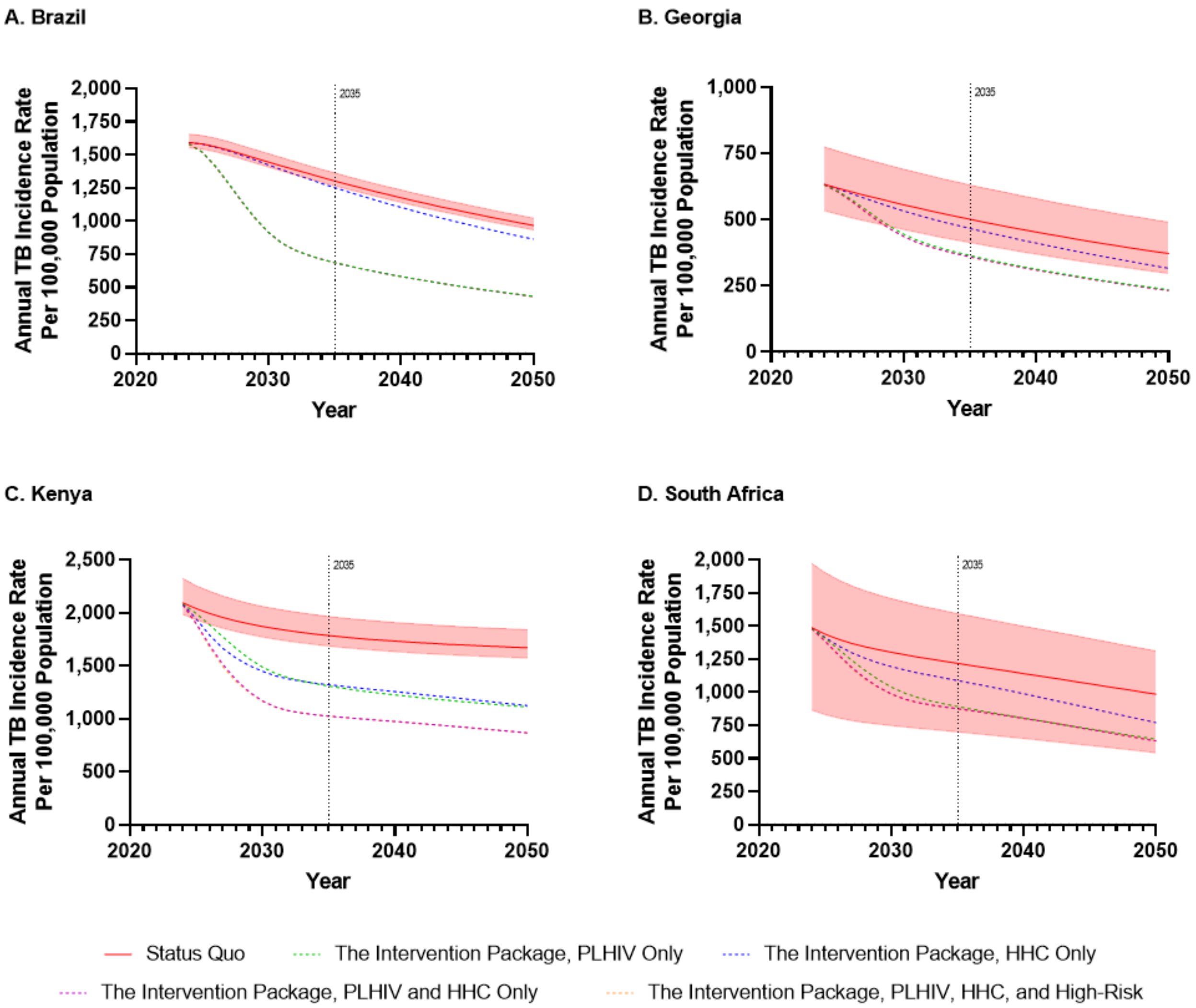

**Caption**: Tuberculosis incidence trajectories from 2024 through to 2050 for PLHIV in each country. Strategies shown are the status quo and The Intervention Package scaled up with TPT in different priority populations. Different incidence scales have been used for each graph. Shaded regions represent the 95% uncertainty range of the status quo. Uncertainty ranges are not presented for others for clarity.

Abbreviations: TB, tuberculosis; PLHIV, people living with HIV; HHC, household contacts.

#### Outcomes (Relative) Among High-Risk Communities by 2050

| Country and Comparison | Number Developing TB | Number Dying from TB | Years of Life Lost due to TB | TB-Associated DALYs |
|---|---|---|---|---|
| Brazil | | | | |
| The Intervention Package without TPT Compared to Status Quo | -8.7% (-9.2% to -8.3%) | -9.1% (-9.6% to -8.7%) | -10% (-10.6% to -9.7%) | -9.8% (-10.3% to -9.4%) |
| The Intervention Package with TPT Compared to The Intervention Package without TPT | -8.8% (-9% to -8.6%) | -10.1% (-10.3% to -9.9%) | -6.8% (-7.1% to -6.5%) | -6.6% (-6.9% to -6.4%) |
| Georgia | | | | |
| The Intervention Package without TPT Compared to Status Quo | -19.2% (-24.2% to -15.7%) | -21.6% (-26.2% to -18.2%) | -22.1% (-26.4% to -19%) | -15.3% (-19.6% to -12.4%) |
| The Intervention Package with TPT Compared to The Intervention Package without TPT | -5.6% (-6.5% to -5%) | -6% (-6.8% to -5.4%) | -5% (-5.8% to -4.4%) | -3.2% (-4% to -2.7%) |
| Kenya | | | | |
| The Intervention Package without TPT Compared to Status Quo | -30.1% (-38.1% to -26.6%) | -30.2% (-38% to -26.6%) | -30.1% (-36.3% to -27.3%) | -28.3% (-35.1% to -25.4%) |
| The Intervention Package with TPT Compared to The Intervention Package without TPT | -6.6% (-7.2% to -5.7%) | -6.5% (-7.1% to -5.7%) | -6.1% (-7% to -5.2%) | -5.6% (-6.2% to -4.8%) |
| South Africa | | | | |
| The Intervention Package without TPT Compared to Status Quo | -20.4% (-23.5% to -17.4%) | -20% (-23% to -17%) | -22.6% (-26.6% to -19.8%) | -21.6% (-25.2% to -18.9%) |
| The Intervention Package with TPT Compared to The Intervention Package without TPT | -4.9% (-5.8% to -3.9%) | -4.4% (-5.2% to -3.5%) | -4.6% (-6.1% to -3.2%) | -4.4% (-5.7% to -3%) |

Notes: Value represent mean (95% UR). All costs are in 2023 USD. No discounting.
Abbreviations: TPT, tuberculosis preventive treatment; TB, tuberculosis; DALY, disability-adjusted life years

#### Annual Change in TB Disease Incidence Among People in High-Risk Communities with The Intervention Package with TPT

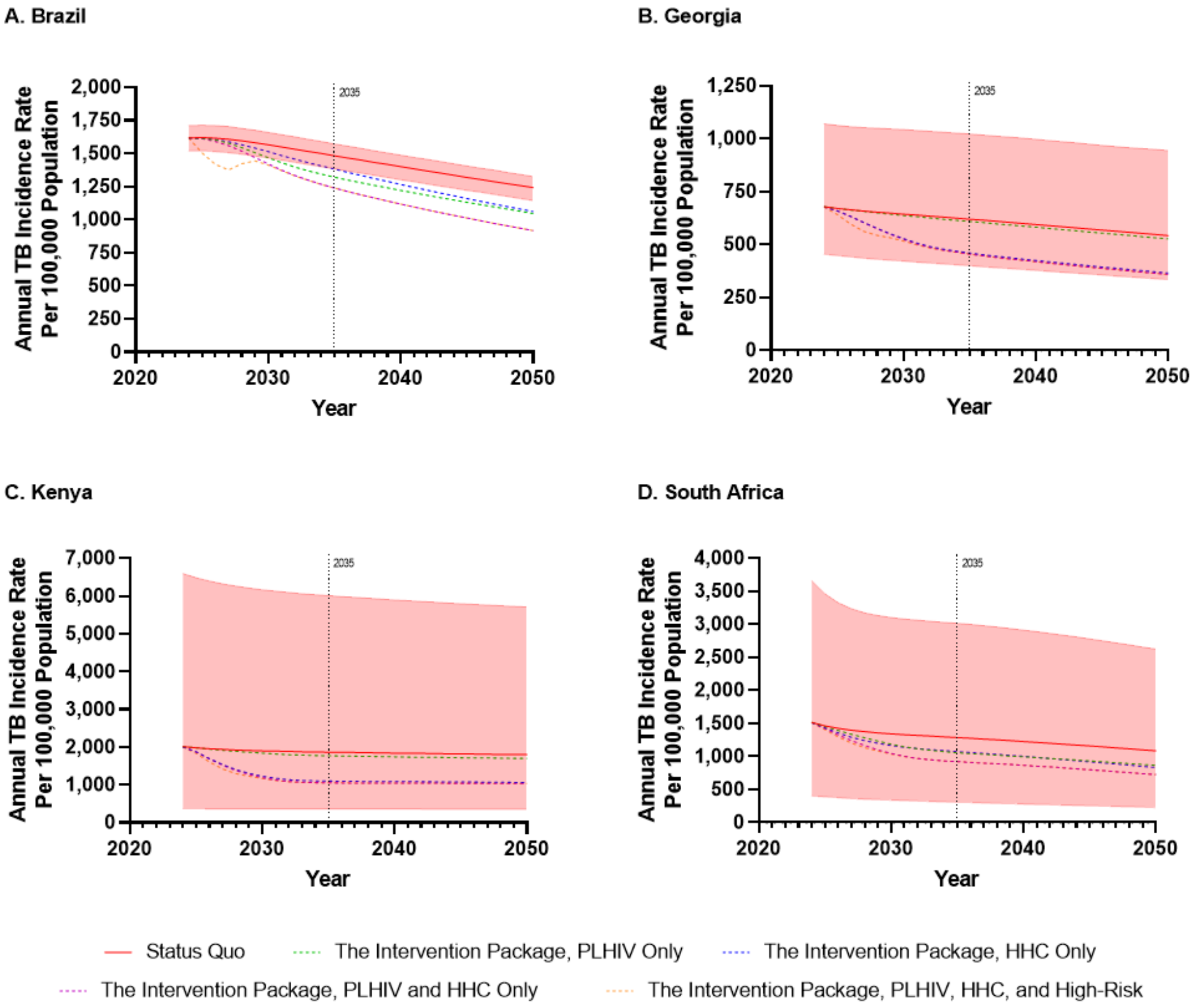

**Caption**: Tuberculosis incidence trajectories from 2024 through to 2050 for the high-risk communities modelled in each country. Strategies shown are the status quo and The Intervention Package scaled up with TPT in different priority populations. Different incidence scales have been used for each graph. Shaded regions represent the 95% uncertainty range of the status quo. Uncertainty ranges are not presented for others for clarity.

Abbreviations: TB, tuberculosis; PLHIV, people living with HIV; HHC, household contacts.

#### Outcomes Among Household Contacts and the Rest of the Population by 2050

| Country and Comparison | Number Developing TB | Number Dying from TB | Years of Life Lost due to TB | TB-Associated DALYs |
|---|---|---|---|---|
| Brazil | | | | |
| The Intervention Package without TPT Compared to Status Quo | -6.1% (-6.8% to -5.7%) | -8.2% (-9.2% to -7.6%) | -8.3% (-9.2% to -7.8%) | -6.1% (-6.9% to -5.6%) |
| The Intervention Package with TPT Compared to The Intervention Package without TPT | -5.8% (-6.2% to -5.5%) | -5.3% (-5.7% to -5%) | -6.3% (-6.7% to -6%) | -4.6% (-4.9% to -4.3%) |
| Georgia | | | | |
| The Intervention Package without TPT Compared to Status Quo | -14.3% (-19.6% to -10.9%) | -28.4% (-35% to -23.7%) | -28.4% (-34.7% to -23.8%) | -18.2% (-23.6% to -14.5%) |
| The Intervention Package with TPT Compared to The Intervention Package without TPT | -10.8% (-12.4% to -9.6%) | -8.5% (-9.5% to -7.8%) | -9% (-10.2% to -8.1%) | -5.3% (-6.3% to -4.7%) |
| Kenya | | | | |
| The Intervention Package without TPT Compared to Status Quo | -31.4% (-32.5% to -30.3%) | -56.2% (-60.7% to -54.3%) | -54.1% (-57.9% to -52.4%) | -48.3% (-51.8% to -46.6%) |
| The Intervention Package with TPT Compared to The Intervention Package without TPT | -21.4% (-23.2% to -20%) | -14.8% (-16.1% to -13.8%) | -15.5% (-16.9% to -14.6%) | -13.1% (-14.1% to -12.3%) |
| South Africa | | | | |
| The Intervention Package without TPT Compared to Status Quo | -32.7% (-39.9% to -30.5%) | -65.5% (-73% to -58.6%) | -54.6% (-60.2% to -45.4%) | -50.7% (-56.1% to -41%) |
| The Intervention Package with TPT Compared to The Intervention Package without TPT | -27.9% (-33.2% to -21.4%) | -24.4% (-31.3% to -17.7%) | -18.4% (-21.5% to -12.9%) | -16.6% (-19.4% to -11.5%) |

Notes: Value represent mean (95% UR). All costs are in 2023 USD. No discounting.

Abbreviations: TPT, tuberculosis preventive treatment; TB, tuberculosis; DALY, disability-adjusted life years

#### Annual Change in TB Disease Incidence Among Household Contacts and the Rest of the Population with The Intervention Package with TPT

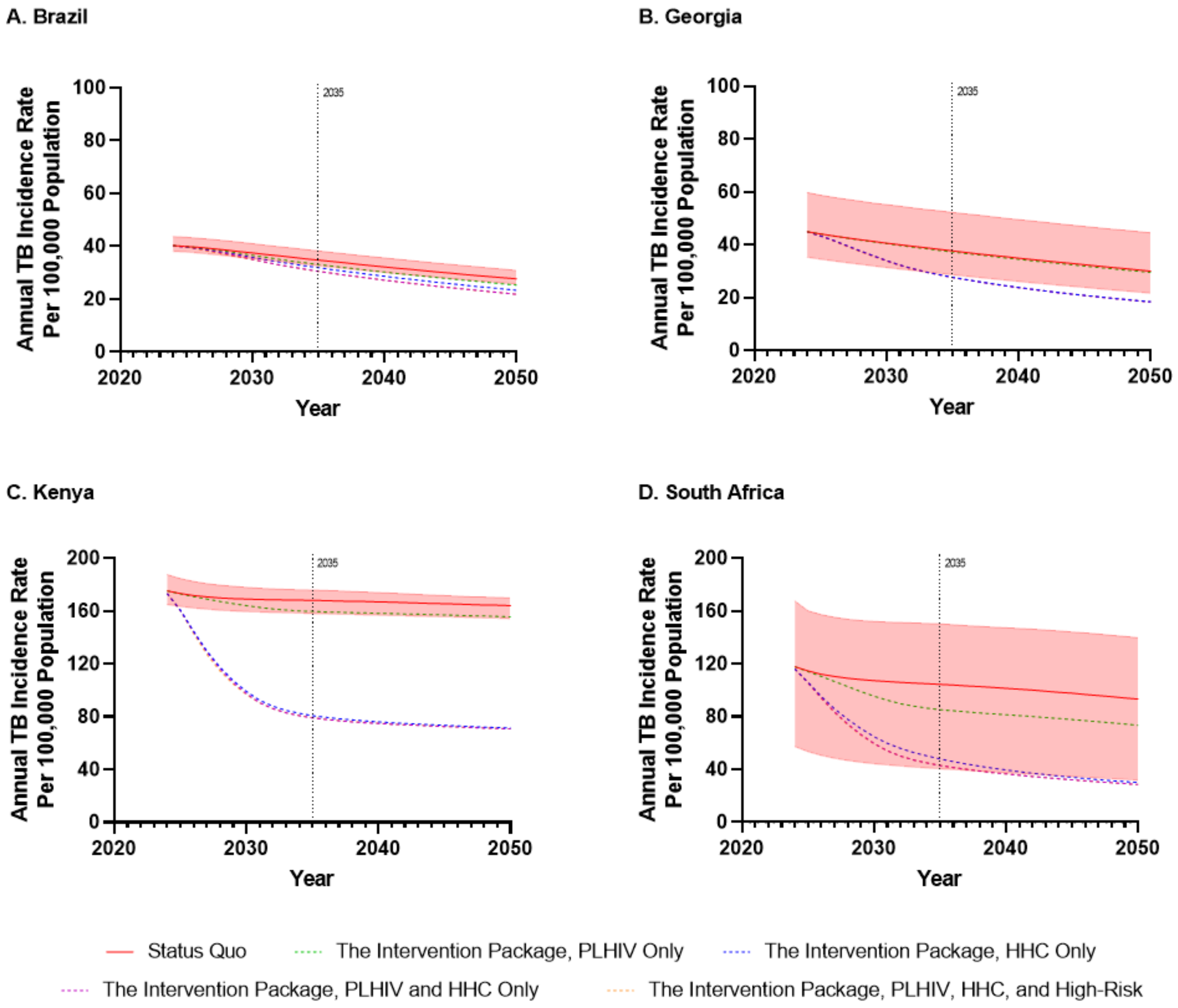

**Caption**: Tuberculosis incidence trajectories from 2024 through to 2050 for household contacts and the rest of the population in each country. Strategies shown are the status quo and The Intervention Package scaled up with TPT in different priority populations. Different incidence scales have been used for each graph. Shaded regions represent the 95% uncertainty range of the status quo. Uncertainty ranges are not presented for others for clarity.

Abbreviations: TB, tuberculosis; PLHIV, people living with HIV; HHC, household contacts.

#### Relative Impact of Differential Targeting of The Intervention Package on Tuberculosis Incidence in Each Priority Group by 2050

| Country and Comparison | The Intervention Package with TPT Only Among PLHIV | The Intervention Package with TPT Only Among HHC | The Intervention Package with TPT Only Among PLHIV and HHC | The Intervention Package with TPT Only Among All Priority Groups |
|---|---|---|---|---|
| Brazil | | | | |
| Change in TB Disease Incidence Among PLHIV Compared to Status Quo | -40.8%<br>(-41.1% to -40.6%) | -4.7%<br>(-5% to -4.5%) | -40.9%<br>(-41.2% to -40.6%) | -40.9%<br>(-41.3% to -40.7%) |
| Change in TB Disease Incidence Among HHC and the Rest of the Population Compared to Status Quo | -4.4%<br>(-4.6% to -4.3%) | -8.1%<br>(-9.1% to -7.4%) | -11.3%<br>(-12.4% to -10.7%) | -11.5%<br>(-12.6% to -10.8%) |
| Change in TB Disease Incidence Among High-Risk Communities Compared to Status Quo | -9.6%<br>(-9.6% to -9.6%) | -7.2%<br>(-8% to -6.6%) | -15.1%<br>(-15.8% to -14.6%) | -16.7%<br>(-17.4% to -16.2%) |
| Georgia | | | | |
| Change in TB Disease Incidence Among PLHIV Compared to Status Quo | -25%<br>(-26.5% to -24%) | -7.5%<br>(-10.3% to -5.9%) | -25.9%<br>(-28.5% to -24.4%) | -26%<br>(-28.7% to -24.4%) |
| Change in TB Disease Incidence Among HHC and the Rest of the Population Compared to Status Quo | -0.9%<br>(-1.1% to -0.8%) | -23.3%<br>(-29.2% to -19.1%) | -23.4%<br>(-29.4% to -19.3%) | -23.6%<br>(-29.5% to -19.4%) |
| Change in TB Disease Incidence Among High-Risk Communities Compared to Status Quo | -1.7%<br>(-1.9% to -1.5%) | -22.1%<br>(-27.3% to -18.3%) | -22.8%<br>(-28% to -19%) | -23.8%<br>(-29% to -19.9%) |
| Kenya | | | | |
| Change in TB Disease Incidence Among PLHIV Compared to Status Quo | -22%<br>(-22.6% to -21.2%) | -22.5%<br>(-24.3% to -21.2%) | -35.5%<br>(-37.1% to -33.8%) | -35.7%<br>(-37.7% to -34%) |
| Change in TB Disease Incidence Among HHC and the Rest of the Population Compared to Status Quo | -4%<br>(-5.4% to -3%) | -45.3%<br>(-47.3% to -43.4%) | -45.9%<br>(-47.9% to -44.2%) | -46%<br>(-48.1% to -44.3%) |
| Change in TB Disease Incidence Among High-Risk Communities Compared to Status Quo | -4.1%<br>(-4.9% to -3.6%) | -32.2%<br>(-39.6% to -28.8%) | -34.1%<br>(-41.4% to -30.8%) | -34.7%<br>(-42.5% to -31.2%) |
| South Africa | | | | |
| Change in TB Disease Incidence Among PLHIV Compared to Status Quo | -22.4%<br>(-26.3% to -20.5%) | -10.2%<br>(-13.1% to -7%) | -23.9%<br>(-26.8% to -22.9%) | -24%<br>(-26.9% to -22.9%) |
| Change in TB Disease Incidence Among HHC and the Rest of the Population Compared to Status Quo | -15.3%<br>(-18.4% to -12.6%) | -48.4%<br>(-54.2% to -43.5%) | -51.5%<br>(-56.1% to -48.1%) | -51.5%<br>(-56.2% to -48.2%) |
| Change in TB Disease Incidence Among High-Risk Communities Compared to Status Quo | -14.6%<br>(-16.5% to -13.4%) | -15.1%<br>(-18.6% to -11.6%) | -24%<br>(-26% to -21.3%) | -24.4%<br>(-26.6% to -21.5%) |

Notes: Value represent mean (95% UR). All costs are in 2023 USD. No discounting.
Abbreviations: TPT, tuberculosis preventive treatment; TB, tuberculosis; PLHIV, people living with HIV; HHC, household contacts.

#### Incremental Outcomes (Absolute Values), by 2050, Discount rate of 3%

| Country and Comparison | Number Developing TB | Number Dying from TB | Years of Life Lost due to TB | TB-Associated DALYs | Number Screened for TB | Number Initiating TPT | TB-Related Health System Cost | TB-Associated Patient and Family Costs | TB-Related Societal Cost |
|---|---|---|---|---|---|---|---|---|---|
| Brazil | | | | | | | | | |
| The Intervention Package without TPT Compared to Status Quo | -167,553 (-188,545 to -154,026) | -20,954 (-23,555 to -19,287) | -768,942 (-872,488 to -703,079) | -796,688 (-904,906 to -727,860) | 28,381,365 (26,566,223 to 31,169,100) | -20,674 (-25,868 to -17,699) | 0.89 billion (0.02 billion to 1.55 billion) | -30 million (-72 million to -8 million) | -5.99 billion (-7.37 billion to -5 billion) |
| The Intervention Package with TPT Compared to The Intervention Package without TPT | -143,158 (-157,290 to -133,758) | -12,953 (-14,199 to -12,105) | -484,741 (-535,390 to -450,877) | -508,961 (-562,604 to -473,099) | -2,855,468 (-3,391,662 to -2,516,021) | 10,244,645 (9,645,601 to 11,167,244) | 0.2 billion (0.06 billion to 0.39 billion) | 339 million (-35 million to 1068 million) | -3.78 billion (-4.51 billion to -2.99 billion) |
| Georgia | | | | | | | | | |
| The Intervention Package without TPT Compared to Status Quo | -4,123 (-7,628 to -2,363) | -520 (-849 to -336) | -19,548 (-32,519 to -12,517) | -22,170 (-36,686 to -14,189) | 1,821,333 (1,521,050 to 2,277,236) | -912 (-1,199 to -703) | 25 million (3 million to 48 million) | -4 million (-12 million to -1 million) | -0.11 billion (-0.2 billion to -0.06 billion) |
| The Intervention Package with TPT Compared to The Intervention Package without TPT | -2,412 (-3,551 to -1,717) | -121 (-164 to -93) | -4,491 (-6,289 to -3,383) | -5,266 (-7,447 to -3,928) | -414,704 (-621,738 to -292,489) | 556,938 (461,081 to 696,651) | 17 million (8 million to 31 million) | 24 million (-1 million to 73 million) | 0.01 billion (-0.02 billion to 0.06 billion) |
| Kenya | | | | | | | | | |
| The Intervention Package without TPT Compared to Status Quo | -639,544 (-887,248 to -565,891) | -138,898 (-181,872 to -123,652) | -5,461,905 (-7,174,627 to -4,848,080) | -5,753,266 (-7,536,777 to -5,113,347) | 75,896,674 (65,415,955 to 94,882,923) | -3,643,334 (-4,561,208 to -3,103,666) | 0.87 billion (-0.45 billion to 3.22 billion) | -43 million (-99 million to -12 million) | -10.64 billion (-14.38 billion to -8.04 billion) |
| The Intervention Package with TPT Compared to The Intervention Package without TPT | -259,673 (-305,283 to -232,780) | -19,120 (-22,153 to -17,517) | -790,850 (-927,463 to -723,719) | -866,217 (-1,013,426 to -792,692) | -47,015,753 (-63,642,559 to -39,686,106) | 20,294,109 (18,292,594 to 22,485,836) | -0.23 billion (-1.05 billion to 0.28 billion) | 141 million (-38 million to 451 million) | -1.75 billion (-2.72 billion to -1.13 billion) |
| South Africa | | | | | | | | | |
| The Intervention Package without TPT Compared to Status Quo | -658,564 (-1,055,341 to -367,445) | -168,757 (-271,439 to -70,900) | -6,108,058 (-9,568,988 to -2,591,113) | -6,453,113 (-10,045,403 to -2,784,900) | 118,252,864 (66,852,256 to 166,879,734) | -435,782 (-1,074,257 to -49,310) | 1.61 billion (-1.74 billion to 4.47 billion) | -94 million (-241 million to -14 million) | -39.88 billion (-63.47 billion to -14.85 billion) |
| The Intervention Package with TPT Compared to The Intervention Package without TPT | -285,684 (-348,305 to -175,262) | -17,941 (-23,496 to -11,283) | -776,884 (-984,769 to -462,037) | -873,183 (-1,087,104 to -524,803) | -66,075,894 (-105,711,861 to -25,984,538) | 28,864,203 (21,351,763 to 34,815,124) | -0.29 billion (-1.27 billion to 0.31 billion) | 234 million (-95 million to 801 million) | -5.32 billion (-7.27 billion to -2.72 billion) |

Notes: Value represent mean (95% UR). All costs are in 2023 USD. No discounting.

Abbreviations: TPT, tuberculosis preventive treatment; TB, tuberculosis; DALY, disability-adjusted life years

#### Incremental Outcomes (Absolute Values), by 2050, Discount Rate of 4% for Brazil, Georgia, and South Africa and Rate of 5% for Kenya

| Country and Comparison | Number Developing TB | Number Dying from TB | Years of Life Lost due to TB | TB-Associated DALYs | Number Screened for TB | Number Initiating TPT | TB-Related Health System Cost | TB-Associated Patient and Family Costs | TB-Related Societal Cost |
|---|---|---|---|---|---|---|---|---|---|
| Brazil (4% discount) | | | | | | | | | |
| The Intervention Package without TPT Compared to Status Quo | -147,365 (-165,698 to -135,527) | -18,464 (-20,744 to -17,001) | -677,093 (-767,784 to -619,319) | -701,296 (-796,031 to -640,952) | 25,313,678 (23,703,692 to 27,786,101) | -18,380 (-22,963 to -15,750) | 0.8 billion (0 billion to 1.39 billion) | -25 million (-62 million to -7 million) | -5.27 billion (-6.49 billion to -4.39 billion) |
| The Intervention Package with TPT Compared to The Intervention Package without TPT | -123,642 (-135,714 to -115,615) | -11,225 (-12,294 to -10,497) | -419,407 (-462,810 to -390,370) | -439,836 (-485,780 to -409,071) | -2,446,196 (-2,904,838 to -2,155,820) | 9,249,247 (8,711,300 to 10,077,960) | 0.19 billion (0.06 billion to 0.35 billion) | 316 million (-27 million to 984 million) | -3.24 billion (-3.88 billion to -2.53 billion) |
| Georgia (4% discount) | | | | | | | | | |
| The Intervention Package without TPT Compared to Status Quo | -3,629 (-6,701 to -2,084) | -461 (-752 to -299) | -17,300 (-28,728 to -11,096) | -19,599 (-32,373 to -12,566) | 1,626,226 (1,358,812 to 2,031,849) | -814 (-1,068 to -629) | 23 million (2 million to 43 million) | -4 million (-11 million to -1 million) | -0.1 billion (-0.18 billion to -0.05 billion) |
| The Intervention Package with TPT Compared to The Intervention Package without TPT | -2,066 (-3,041 to -1,471) | -103 (-140 to -80) | -3,839 (-5,375 to -2,894) | -4,483 (-6,337 to -3,346) | -357,968 (-537,241 to -252,192) | 505,917 (418,060 to 634,156) | 16 million (8 million to 28 million) | 23 million (0 million to 67 million) | 0.01 billion (-0.01 billion to 0.06 billion) |
| Kenya (5% discount) | | | | | | | | | |
| The Intervention Package without TPT Compared to Status Quo | -505,607 (-705,601 to -446,027) | -110,285 (-144,974 to -98,013) | -4,328,774 (-5,706,555 to -3,836,185) | -4,555,930 (-5,991,066 to -4,042,244) | 60,638,034 (52,266,334 to 75,746,270) | -2,897,870 (-3,643,349 to -2,461,307) | 0.69 billion (-0.35 billion to 2.53 billion) | -32 million (-74 million to -9 million) | -8.43 billion (-11.43 billion to -6.38 billion) |
| The Intervention Package with TPT Compared to The Intervention Package without TPT | -194,902 (-229,301 to -174,841) | -14,489 (-16,808 to -13,287) | -596,077 (-699,599 to -546,078) | -649,781 (-760,901 to -595,229) | -36,053,299 (-49,039,253 to -30,423,965) | 17,482,051 (15,722,669 to 19,448,320) | -0.1 billion (-0.71 billion to 0.3 billion) | 135 million (-22 million to 422 million) | -1.21 billion (-1.94 billion to -0.7 billion) |
| South Africa (4% discount) | | | | | | | | | |
| The Intervention Package without TPT Compared to Status Quo | -581,280 (-932,546 to -324,877) | -150,062 (-241,219 to -63,161) | -5,429,541 (-8,502,166 to -2,305,013) | -5,733,651 (-8,923,972 to -2,476,005) | 106,179,317 (60,106,818 to 149,559,695) | -391,024 (-961,406 to -46,899) | 1.43 billion (-1.6 billion to 4.01 billion) | -80 million (-205 million to -10 million) | -35.45 billion (-56.38 billion to -13.21 billion) |
| The Intervention Package with TPT Compared to The Intervention Package without TPT | -250,843 (-304,776 to -154,814) | -15,761 (-20,804 to -9,898) | -678,969 (-861,260 to -404,197) | -761,388 (-950,912 to -458,158) | -58,429,468 (-93,500,370 to -22,950,812) | 26,763,997 (19,792,751 to 32,312,151) | -0.22 billion (-1.07 billion to 0.3 billion) | 234 million (-81 million to 786 million) | -4.59 billion (-6.27 billion to -2.35 billion) |

Notes: Value represent mean (95% UR). All costs are in 2023 USD. No discounting.

Abbreviations: TPT, tuberculosis preventive treatment; TB, tuberculosis; DALY, disability-adjusted life years

#### Cost Effectiveness and Return on Investment by 2050, Discount Rate of 3%

| Outcome | Brazil | Georgia | Kenya | South Africa |
|---|---|---|---|---|
| **Intervention Package without TPT vs. Status Quo** | | | | |
| Return on Investment | 7.7 | 5.3 | 13.2 | 25.8 |
| Incremental Health System Cost per DALY averted | $1120 | $1146 | $151 | $249 |
| Incremental Health System Cost per TB case averted | $5327 | $6163 | $1359 | $2438 |
| Incremental Health System Cost per TB death averted | $42,593 | $48,868 | $6257 | $9514 |
| Incremental Societal Cost per DALY averted | Cost Saving | Cost Saving | Cost Saving | Cost Saving |
| Incremental Societal Cost per TB case averted | Cost Saving | Cost Saving | Cost Saving | Cost Saving |
| Incremental Societal Cost per TB death averted | Cost Saving | Cost Saving | Cost Saving | Cost Saving |
| **Intervention Package with TPT vs. Intervention Package without TPT** | | | | |
| Return on Investment | 19.9 | 0.3 | Both Health System and Societal Savings | Both Health System and Societal Savings |
| Incremental Health System Cost per DALY averted | $394 | $3247 | Cost Saving | Cost Saving |
| Incremental Health System Cost per TB case averted | $1401 | $7089 | Cost Saving | Cost Saving |
| Incremental Health System Cost per TB death averted | $15,481 | $141,303 | Cost Saving | Cost Saving |
| Incremental Societal Cost per DALY averted | Cost Saving | $2224 | Cost Saving | Cost Saving |
| Incremental Societal Cost per TB case averted | Cost Saving | $4856 | Cost Saving | Cost Saving |
| Incremental Societal Cost per TB death averted | Cost Saving | $96,801 | Cost Saving | Cost Saving |
| **Intervention Package with TPT vs. Status Quo** | | | | |
| Return on Investment | 9.9 | 3.3 | 20.5 | 35.3 |
| Incremental Health System Cost per DALY averted | $837 | $1549 | $96 | $180 |
| Incremental Health System Cost per TB case averted | $3518 | $6505 | $707 | $1395 |
| Incremental Health System Cost per TB death averted | $32,236 | $66,317 | $4025 | $7057 |
| Incremental Societal Cost per DALY averted | Cost Saving | Cost Saving | Cost Saving | Cost Saving |
| Incremental Societal Cost per TB case averted | Cost Saving | Cost Saving | Cost Saving | Cost Saving |
| Incremental Societal Cost per TB death averted | Cost Saving | Cost Saving | Cost Saving | Cost Saving |

Notes: All costs are in 2023 USD. Return on investment is defined as the societal return per $1 USD invested by the health system. It cannot be calculated if there are projected health system cost savings. A return on investment of 8 means that $8 USD are returned to society for every $1 USD invested by the health system. Willingness to pay per DALY averted by country—Brazil, $13,644; Georgia, $1603; Kenya, $1002; South Africa, $4834.
Abbreviations: TPT, tuberculosis preventive treatment; DALY, disability adjusted life year; TB, tuberculosis

#### Cost Effectiveness and Return on Investment by 2050, Discount Rate of 4% for Brazil, Georgia, and South Africa and Rate of 5% for Kenya

| **Outcome** | **Brazil** (4% discount) | **Georgia** (4% discount) | **Kenya** (5% discount) | **South Africa** (4% discount) |
|---|---|---|---|---|
| **Intervention Package without TPT vs. Status Quo** | | | | |
| Return on Investment | 7.6 | 5.2 | 13.2 | 25.9 |
| Incremental Health System Cost per DALY averted | $1136 | $1157 | $151 | $249 |
| Incremental Health System Cost per TB case averted | $5408 | $6249 | $1365 | $2453 |
| Incremental Health System Cost per TB death averted | $43,164 | $49,194 | $6256 | $9503 |
| Incremental Societal Cost per DALY averted | Cost Saving | Cost Saving | Cost Saving | Cost Saving |
| Incremental Societal Cost per TB case averted | Cost Saving | Cost Saving | Cost Saving | Cost Saving |
| Incremental Societal Cost per TB death averted | Cost Saving | Cost Saving | Cost Saving | Cost Saving |
| **Intervention Package with TPT vs. Intervention Package without TPT** | | | | |
| Return on Investment | 18.4 | 0.2 | Both Health System and Societal Savings | Both Health System and Societal Savings |
| Incremental Health System Cost per DALY averted | $423 | $3544 | Cost Saving | Cost Saving |
| Incremental Health System Cost per TB case averted | $1503 | $7690 | Cost Saving | Cost Saving |
| Incremental Health System Cost per TB death averted | $16,559 | $154,256 | Cost Saving | Cost Saving |
| Incremental Societal Cost per DALY averted | Cost Saving | $2923.00 | Cost Saving | Cost Saving |
| Incremental Societal Cost per TB case averted | Cost Saving | $6342 | Cost Saving | Cost Saving |
| Incremental Societal Cost per TB death averted | Cost Saving | $127,217 | Cost Saving | Cost Saving |
| **Intervention Package with TPT vs. Status Quo** | | | | |
| Return on Investment | 9.7 | 3.1 | 17.2 | 34.2 |
| Incremental Health System Cost per DALY averted | $861 | $1601 | $114 | $186 |
| Incremental Health System Cost per TB case averted | $3627 | $6772 | $848 | $1451 |
| Incremental Health System Cost per TB death averted | $33,105 | $68,380 | $4759 | $7282 |
| Incremental Societal Cost per DALY averted | Cost Saving | Cost Saving | Cost Saving | Cost Saving |
| Incremental Societal Cost per TB case averted | Cost Saving | Cost Saving | Cost Saving | Cost Saving |
| Incremental Societal Cost per TB death averted | Cost Saving | Cost Saving | Cost Saving | Cost Saving |

Notes: All costs are in 2023 USD. Return on investment is defined as the societal return per $1 USD invested by the health system. It cannot be calculated if there are projected health system cost savings. A return on investment of 8 means that $8 USD are returned to society for every $1 USD invested by the health system. Willingness to pay per DALY averted by country—Brazil, $13,644; Georgia, $1603; Kenya, $1002; South Africa, $4834.
Abbreviations: TPT, tuberculosis preventive treatment; DALY, disability adjusted life year; TB, tuberculosis

#### Outcomes under the status quo, 2024-2035

| Outcome | Brazil 2024-2035 | Georgia 2024-2035 | Kenya 2024-2035 | South Africa 2024-2035 |
|---|---|---|---|---|
| **Total People Developing TB** | 1,303,429<br>(1,239,728 to 1,398,834) | 20,910<br>(16,225 to 28,399) | 1,443,660<br>(1,319,152 to 1,850,448) | 2,193,077<br>(1,476,317 to 2,913,501) |
| **Total People Dying from TB** | 109,132<br>(103,219 to 118,110) | 1,443<br>(1,149 to 1,901) | 218,811<br>(202,307 to 273,453) | 354,462<br>(172,038 to 562,498) |
| **Total Years of Life Lost due to TB** | 4,214,436<br>(3,971,529 to 4,586,186) | 53,327<br>(41,701 to 72,103) | 8,487,465<br>(7,772,222 to 10,795,548) | 12,284,555<br>(6,296,919 to 18,614,855) |
| **Total DALYs** | 5,444,693<br>(5,163,564 to 5,873,708) | 90,235<br>(73,519 to 116,257) | 9,716,551<br>(8,920,241 to 12,181,852) | 14,187,116<br>(7,701,096 to 20,627,256) |
| **Total Health System Cost** | 1.08 billion<br>(0.58 billion to 1.92 billion) | 24 million<br>(14 million to 42 million) | 1.49 billion<br>(0.71 billion to 2.98 billion) | 1.45 billion<br>(0.39 billion to 4.26 billion) |
| **Total Costs to Patients and Families** | 849 million<br>(162 million to 2259 million) | 32 million<br>(7 million to 77 million) | 200 million<br>(41 million to 504 million) | 743 million<br>(170 million to 1925 million) |
| **Total Societal Cost** | 39.51 billion<br>(36.81 billion to 43.67 billion) | 0.41 billion<br>(0.31 billion to 0.55 billion) | 19.5 billion<br>(17.36 billion to 24.77 billion) | 85.44 billion<br>(44.33 billion to 128.89 billion) |

#### Incremental Outcomes (Absolute Values), by 2035, including the enhanced package.

| Country and Comparison | Number Developing TB | Number Dying from TB | Years of Life Lost due to TB | TB-Associated DALYs | Number Screened for TB | Number Initiating TPT | TB-Related Health System Cost | TB-Associated Patient and Family Costs | TB-Related Societal Cost |
|---|---|---|---|---|---|---|---|---|---|
| Brazil | | | | | | | | | |
| The Intervention Package without TPT Compared to Status Quo | -87,833 (-97,875 to -81,149) | -11,311 (-12,622 to -10,449) | -411,260 (-462,922 to -377,760) | -424,060 (-477,626 to -389,312) | 17,642,777 (16,575,373 to 19,279,283) | -12,467 (-15,337 to -10,795) | 0.54 billion (-0.06 billion to 0.96 billion) | -9 million (-31 million to 6 million) | -3.14 billion (-3.95 billion to -2.57 billion) |
| The Intervention Package with TPT Compared to The Intervention Package without TPT | -55,907 (-60,212 to -53,039) | -5,417 (-5,845 to -5,126) | -196,552 (-213,264 to -185,228) | -201,919 (-219,196 to -190,112) | -929,555 (-1,097,965 to -822,778) | 7,224,993 (6,813,609 to 7,859,783) | 0.19 billion (0.09 billion to 0.31 billion) | 303 million (16 million to 889 million) | -1.26 billion (-1.64 billion to -0.67 billion) |
| Enhanced Package with TPT Compared to The Intervention Package with TPT | -56,667 (-63,722 to -52,002) | -5,944 (-6,702 to -5,458) | -220,646 (-250,730 to -201,550) | -228,595 (-259,829 to -208,517) | 10,950,758 (10,429,502 to 11,733,438) | 2,095,254 (1,963,706 to 2,307,429) | 0.41 billion (0.12 billion to 0.8 billion) | -68 million (-174 million to -15 million) | -1.62 billion (-2.09 billion to -1.16 billion) |
| Georgia | | | | | | | | | |
| The Intervention Package without TPT Compared to Status Quo | -2,201 (-3,967 to -1,290) | -304 (-484 to -202) | -11,200 (-18,165 to -7,344) | -12,510 (-20,167 to -8,211) | 1,144,657 (962,417 to 1,417,597) | -580 (-737 to -463) | 16 million (1 million to 29 million) | -2 million (-6 million to 0 million) | -0.06 billion (-0.11 billion to -0.03 billion) |
| The Intervention Package with TPT Compared to The Intervention Package without TPT | -768 (-1,129 to -548) | -39 (-52 to -30) | -1,372 (-1,903 to -1,045) | -1,454 (-2,037 to -1,099) | -159,331 (-244,440 to -109,606) | 421,497 (343,362 to 536,719) | 16 million (9 million to 27 million) | 23 million (2 million to 64 million) | 0.03 billion (0.01 billion to 0.07 billion) |
| Enhanced Package with TPT Compared to The Intervention Package with TPT | -1,320 (-2,007 to -911) | -138 (-190 to -104) | -4,883 (-6,969 to -3,607) | -5,500 (-7,887 to -4,031) | 423,665 (399,724 to 448,774) | 72,446 (62,346 to 84,411) | 3 million (-8 million to 14 million) | -11 million (-30 million to -2 million) | -0.04 billion (-0.07 billion to -0.02 billion) |
| Kenya | | | | | | | | | |
| The Intervention Package without TPT Compared to Status Quo | -375,625 (-538,308 to -325,861) | -82,796 (-110,540 to -73,253) | -3,223,724 (-4,313,534 to -2,841,618) | -3,376,320 (-4,512,637 to -2,978,101) | 45,919,210 (39,795,579 to 57,220,209) | -2,188,804 (-2,804,724 to -1,833,312) | 0.47 billion (-0.27 billion to 1.72 billion) | -20 million (-49 million to -5 million) | -6.32 billion (-8.65 billion to -4.84 billion) |
| The Intervention Package with TPT Compared to The Intervention Package without TPT | -101,009 (-120,231 to -91,278) | -8,185 (-9,557 to -7,595) | -320,916 (-378,337 to -297,173) | -336,716 (-397,506 to -311,187) | -22,534,638 (-31,555,848 to -18,941,588) | 17,395,376 (15,589,461 to 19,494,702) | 0.22 billion (-0.14 billion to 0.53 billion) | 167 million (4 million to 495 million) | -0.29 billion (-0.71 billion to 0.17 billion) |
| Enhanced Package with TPT Compared to The Intervention Package with TPT | -103,614 (-147,832 to -90,151) | -19,175 (-24,601 to -17,620) | -728,889 (-948,341 to -665,628) | -765,951 (-995,403 to -699,147) | 4,422,987 (4,019,188 to 4,592,403) | 1,265,702 (669,977 to 1,561,970) | 0.41 billion (-0.1 billion to 1.39 billion) | -36 million (-92 million to -8 million) | -1.16 billion (-1.86 billion to -0.24 billion) |
| South Africa | | | | | | | | | |
| The Intervention Package without TPT Compared to Status Quo | -362,653 (-587,823 to -206,768) | -101,289 (-161,974 to -43,479) | -3,648,285 (-5,689,346 to -1,565,074) | -3,832,457 (-5,930,093 to -1,670,705) | 78,611,940 (45,064,662 to 108,772,832) | -291,805 (-698,725 to -48,856) | 0.95 billion (-1.32 billion to 2.81 billion) | -31 million (-91 million to 15 million) | -23.81 billion (-37.89 billion to -8.98 billion) |

| Country and Comparison | Number Developing TB | Number Dying from TB | Years of Life Lost due to TB | TB-Associated DALYs | Number Screened for TB | Number Initiating TPT | TB-Related Health System Cost | TB-Associated Patient and Family Costs | TB-Related Societal Cost |
|---|---|---|---|---|---|---|---|---|---|
| The Intervention Package with TPT Compared to The Intervention Package without TPT | -147,915 (-169,411 to -98,948) | -9,550 (-13,723 to -5,887) | -377,800 (-495,718 to -227,447) | -409,640 (-529,314 to -250,542) | -38,379,542 (-61,573,402 to -14,521,136) | 24,433,085 (18,023,811 to 29,385,912) | 0.09 billion (-0.37 billion to 0.42 billion) | 296 million (-16 million to 913 million) | -2.17 billion (-3.13 billion to -0.89 billion) |
| Enhanced Package with TPT Compared to The Intervention Package with TPT | -89,464 (-141,827 to -60,038) | -23,921 (-37,506 to -12,453) | -821,703 (-1,235,543 to -437,646) | -869,786 (-1,290,428 to -469,464) | 11,774,445 (9,274,119 to 14,164,440) | 497,268 (307,145 to 734,966) | 1.81 billion (0.45 billion to 4.7 billion) | -72 million (-202 million to -8 million) | -3.83 billion (-7.16 billion to 0.79 billion) |

Notes: Value represent mean (95% UR). All costs are in 2023 USD. No discounting.

Abbreviations: TPT, tuberculosis preventive treatment; TB, tuberculosis; DALY, disability-adjusted life years

#### Incremental Outcomes (Relative Values), by 2035, including the enhanced package.

| Country and Comparison | Number Developing TB | Number Dying from TB | Years of Life Lost due to TB | TB-Associated DALYs | Number Screened for TB | Number Initiating TPT | TB-Related Health System Cost | TB-Associated Patient and Family Costs | TB-Related Societal Cost |
|---|---|---|---|---|---|---|---|---|---|
| Brazil | | | | | | | | | |
| The Intervention Package without TPT Compared to Status Quo | -6.7% (-7% to -6.5%) | -10.3% (-10.7% to -10.1%) | -9.8% (-10.1% to -9.5%) | -7.8% (-8.1% to -7.5%) | 1960.4% (1890.4% to 2028.2%) | -3.1% (-3.6% to -2.8%) | 57.7% (-3.4% to 128.4%) | -2% (-9.7% to 0.4%) | -7.9% (-9.4% to -6.8%) |
| The Intervention Package with TPT Compared to The Intervention Package without TPT | -4.6% (-4.6% to -4.6%) | -5.5% (-5.6% to -5.5%) | -5.2% (-5.2% to -5.2%) | -4% (-4.1% to -4%) | -5% (-5.4% to -4.7%) | 1856.8% (1796.5% to 1914.8%) | 11.9% (5.1% to 21.9%) | 57.8% (1.4% to 240.1%) | -3.5% (-4.3% to -1.9%) |
| Enhanced Package with TPT Compared to The Intervention Package with TPT | -4.9% (-5.1% to -4.7%) | -6.4% (-6.7% to -6.2%) | -6.1% (-6.4% to -5.9%) | -4.7% (-5% to -4.6%) | 62.2% (61.3% to 62.8%) | 27.5% (27.3% to 27.9%) | 23% (7.5% to 44.5%) | -6.4% (-12.7% to -1.9%) | -4.6% (-5.6% to -3.4%) |
| Georgia | | | | | | | | | |
| The Intervention Package without TPT Compared to Status Quo | -10.3% (-14% to -7.9%) | -20.8% (-25.3% to -17.7%) | -20.8% (-25.2% to -17.6%) | -13.7% (-17.4% to -11.2%) | 15699.4% (11913.4% to 22206.3%) | -19.5% (-25.1% to -16.3%) | 73.1% (1.6% to 157.1%) | -5.7% (-17.8% to 0.4%) | -14.5% (-20% to -9.2%) |
| The Intervention Package with TPT Compared to The Intervention Package without TPT | -4.1% (-4.6% to -3.7%) | -3.4% (-3.7% to -3.2%) | -3.2% (-3.5% to -3%) | -1.9% (-2.1% to -1.7%) | -13.7% (-17.2% to -11.3%) | 17695.4% (14003.2% to 24534.3%) | 41.3% (20.2% to 74.4%) | 109.7% (5.3% to 425.4%) | 8.5% (1.8% to 20.7%) |
| Enhanced Package with TPT Compared to The Intervention Package with TPT | -7.3% (-8.6% to -6.3%) | -12.4% (-14% to -11.3%) | -11.9% (-13.4% to -10.8%) | -7.2% (-8.4% to -6.3%) | 42.8% (38.1% to 46.5%) | 17.2% (15.5% to 18.1%) | 5.5% (-12.8% to 24.1%) | -21.4% (-37.8% to -5.9%) | -10.7% (-15.5% to -6.6%) |
| Kenya | | | | | | | | | |
| The Intervention Package without TPT Compared to Status Quo | -25.9% (-29.3% to -24.3%) | -37.8% (-40.2% to -35.7%) | -37.9% (-39.7% to -35.9%) | -34.7% (-36.8% to -32.7%) | 782.8% (502.4% to 1040.3%) | -40.5% (-41.1% to -39.1%) | 39.6% (-11.4% to 135.3%) | -11.6% (-26.9% to -4.7%) | -32.3% (-36.2% to -26.2%) |
| The Intervention Package with TPT Compared to The Intervention Package without TPT | -9.5% (-9.8% to -8.7%) | -6% (-6.3% to -5.6%) | -6.1% (-6.4% to -5.6%) | -5.3% (-5.6% to -5%) | -43.2% (-49.4% to -40.1%) | 548.2% (376.1% to 661.6%) | 13.8% (-4.1% to 38.2%) | 147.7% (1.9% to 572.4%) | -2.1% (-4.8% to 1.4%) |
| Enhanced Package with TPT Compared to The Intervention Package with TPT | -10.7% (-12.3% to -10.1%) | -15% (-15.8% to -14.6%) | -14.7% (-15.2% to -14.4%) | -12.7% (-13.4% to -12.4%) | 15.1% (12.5% to 16.4%) | 6.2% (3% to 7.9%) | 17.9% (-4.3% to 48%) | -10.3% (-14.2% to -5.6%) | -9% (-13.1% to -1.8%) |
| South Africa | | | | | | | | | |
| The Intervention Package without TPT Compared to Status Quo | -16.3% (-20.2% to -13.7%) | -28.2% (-30.6% to -24.1%) | -29.2% (-31.7% to -23.5%) | -26.4% (-28.9% to -20.8%) | 2827.5% (1219.1% to 5852.4%) | -11.2% (-18.9% to -3%) | 131.2% (-36.3% to 455.4%) | -4.9% (-14.6% to 2.6%) | -27.2% (-30.9% to -19.4%) |

| Country and Comparison | Number Developing TB | Number Dying from TB | Years of Life Lost due to TB | TB-Associated DALYs | Number Screened for TB | Number Initiating TPT | TB-Related Health System Cost | TB-Associated Patient and Family Costs | TB-Related Societal Cost |
|---|---|---|---|---|---|---|---|---|---|
| The Intervention Package with TPT Compared to The Intervention Package without TPT | -8.1% (-9.2% to -6.9%) | -3.9% (-5% to -2.9%) | -4.5% (-5.3% to -3.6%) | -4% (-4.8% to -3.3%) | -45.2% (-55.7% to -29.6%) | 1190.3% (689.4% to 1805.9%) | 6.3% (-10% to 28.4%) | 61.4% (-1.8% to 226.6%) | -3.5% (-4.6% to -2.2%) |
| Enhanced Package with TPT Compared to The Intervention Package with TPT | -5.3% (-7% to -4.7%) | -9.9% (-10.5% to -9.5%) | -9.9% (-10.5% to -9.4%) | -8.7% (-9.4% to -8.1%) | 27.6% (22.8% to 37.5%) | 1.9% (1% to 3.2%) | 69.6% (22.5% to 149.9%) | -7.6% (-16.2% to -1.1%) | -6.1% (-9% to 2.1%) |

Notes: Value represent mean (95% UR). All costs are in 2023 USD. No discounting.

Abbreviations: TPT, tuberculosis preventive treatment; TB, tuberculosis; DALY, disability-adjusted life years

#### Cost Effectiveness and Return on Investment by 2035

| Outcome | Brazil | Georgia | Kenya | South Africa |
|---|---|---|---|---|
| **Intervention Package without TPT vs. Status Quo** | | | | |
| Return on Investment | 6.8 | 4.9 | 14.4 | 26.1 |
| Incremental Health System Cost per DALY averted | $1274 | $1243 | $139 | $247 |
| Incremental Health System Cost per TB case averted | $6151 | $7066 | $1251 | $2614 |
| Incremental Health System Cost per TB death averted | $47,762 | $51,156 | $5674 | $9359 |
| Incremental Societal Cost per DALY averted | Cost Saving | Cost Saving | Cost Saving | Cost Saving |
| Incremental Societal Cost per TB case averted | Cost Saving | Cost Saving | Cost Saving | Cost Saving |
| Incremental Societal Cost per TB death averted | Cost Saving | Cost Saving | Cost Saving | Cost Saving |
| **Intervention Package with TPT vs. Intervention Package without TPT** | | | | |
| Return on Investment | 7.8 | -0.8 | 2.3 | 24.3 |
| Incremental Health System Cost per DALY averted | $918 | $10,965 | $654 | $228 |
| Incremental Health System Cost per TB case averted | $3314 | $20,760 | $2179 | $630 |
| Incremental Health System Cost per TB death averted | $34,207 | $408,805 | $26,889 | $9760 |
| Incremental Societal Cost per DALY averted | Cost Saving | $20,260 | Cost Saving | Cost Saving |
| Incremental Societal Cost per TB case averted | Cost Saving | $38,356 | Cost Saving | Cost Saving |
| Incremental Societal Cost per TB death averted | Cost Saving | $755,316 | Cost Saving | Cost Saving |
| **Intervention Package with TPT vs. Status Quo** | | | | |
| Return on Investment | 7.1 | 2 | 10.6 | 26 |
| Incremental Health System Cost per DALY averted | $1159 | $2255 | $186 | $245 |
| Incremental Health System Cost per TB case averted | $5048 | $10,608 | $1447 | $2039 |
| Incremental Health System Cost per TB death averted | $43,372 | $91,821 | $7583 | $9394 |
| Incremental Societal Cost per DALY averted | Cost Saving | Cost Saving | Cost Saving | Cost Saving |
| Incremental Societal Cost per TB case averted | Cost Saving | Cost Saving | Cost Saving | Cost Saving |
| Incremental Societal Cost per TB death averted | Cost Saving | Cost Saving | Cost Saving | Cost Saving |

Notes: All costs are in 2023 USD. Return on investment is defined as the societal return per $1 USD invested by the health system. It cannot be calculated if there are projected health system cost savings. A return on investment of 8 means that $8 USD are returned to society for every $1 USD invested by the health system. A negative return on investment means a health system investment is not fully recovered through benefits to society; a value of -0.8 suggests that there is an additional cost of $0.80 USD to society per $1 USD invested by the health system. In this analysis, this is solely driven by increasing costs to patients and families associated with tuberculosis preventive treatment that is not recovered through averted tuberculosis disease. Willingness to pay per DALY averted by country—Brazil, $13,644; Georgia, $1603; Kenya, $1002; South Africa, $4834.

Abbreviations: TPT, tuberculosis preventive treatment; DALY, disability adjusted life year; TB, tuberculosis

Impact of The Intervention Package without and with tuberculosis preventive treatment on cases and deaths by 2035

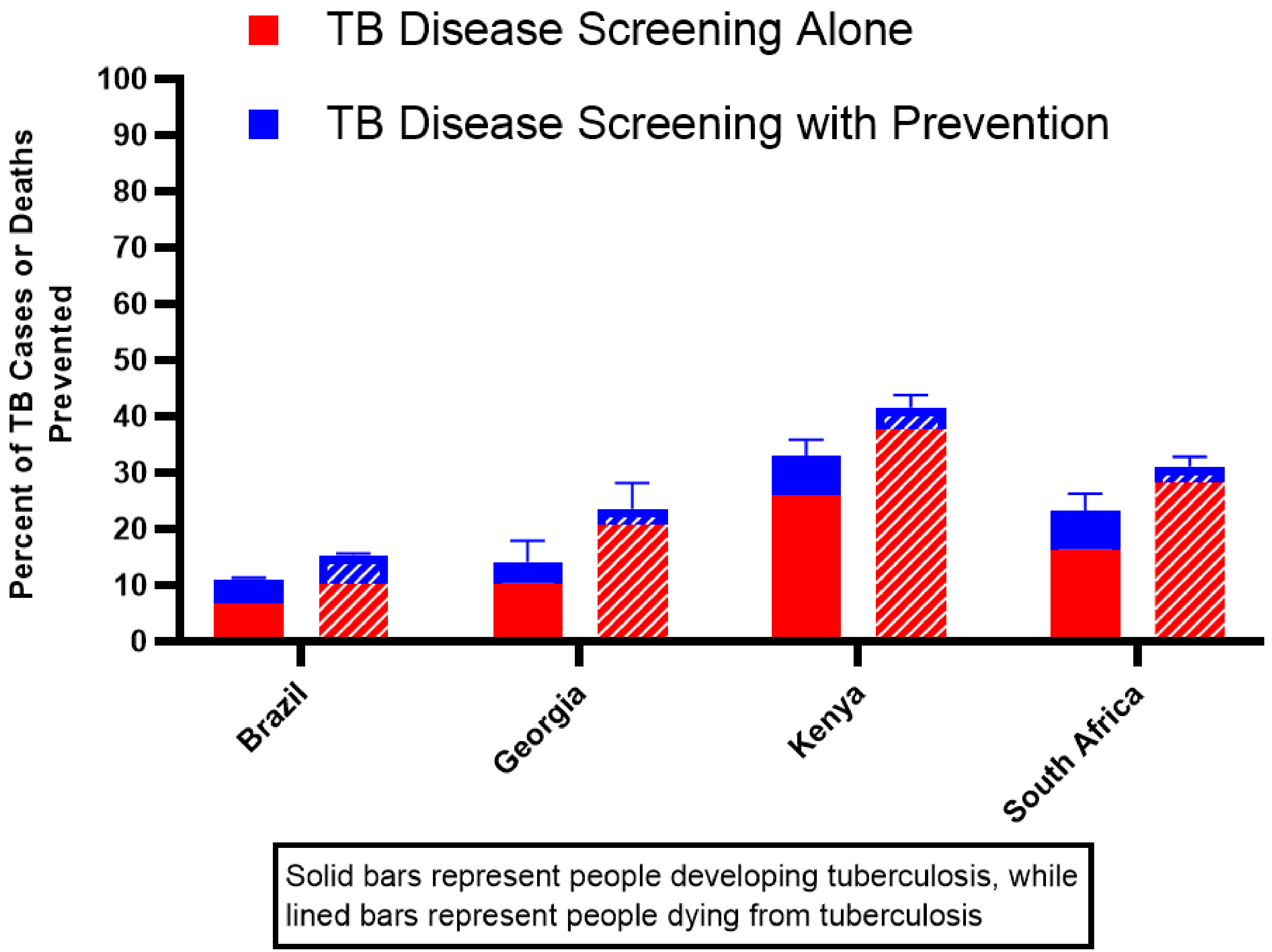

**Caption**: The incremental impact of including TPT in The Intervention Package on cumulative tuberculosis incidence and mortality from 2024 to 2035.

#### Incremental Outcomes (Absolute Values), by 2050, including the enhanced package.

| Country and Comparison | Number Developing TB | Number Dying from TB | Years of Life Lost due to TB | TB-Associated DALYs | Number Screened for TB | Number Initiating TPT | TB-Related Health System Cost | TB-Associated Patient and Family Costs | TB-Related Societal Cost |
|---|---|---|---|---|---|---|---|---|---|
| Brazil | | | | | | | | | |
| The Intervention Package without TPT Compared to Status Quo | -254,971<br><br>(-287,564 to -234,154) | -31,702<br><br>(-35,699 to -29,152) | -1,165,677<br><br>(-1,325,078 to -1,064,709) | -1,209,001<br><br>(-1,375,859 to -1,103,319) | 41,430,986<br><br>(38,737,106 to 45,569,807) | -30,442<br><br>(-38,272 to -25,982) | 1.3 billion<br><br>(0.06 billion to 2.26 billion) | -50 million<br><br>(-113 million to -15 million) | -9.14 billion<br><br>(-11.2 billion to -7.65 billion) |
| The Intervention Package with TPT Compared to The Intervention Package without TPT | -229,540<br><br>(-252,929 to -213,971) | -20,567<br><br>(-22,602 to -19,182) | -773,086<br><br>(-856,161 to -717,643) | -814,593<br><br>(-902,858 to -755,844) | -4,679,987<br><br>(-5,562,827 to -4,121,233) | 14,406,367<br><br>(13,551,628 to 15,721,855) | 0.26 billion<br><br>(0.05 billion to 0.52 billion) | 430 million<br><br>(-90 million to 1412 million) | -6.21 billion<br><br>(-7.33 billion to -5.08 billion) |
| Enhanced Package with TPT Compared to The Intervention Package with TPT | -158,742<br><br>(-179,435 to -145,196) | -14,895<br><br>(-16,873 to -13,631) | -565,744<br><br>(-645,791 to -514,786) | -598,295<br><br>(-683,766 to -543,891) | 22,322,635<br><br>(21,359,404 to 23,751,024) | 2,875,395<br><br>(2,744,706 to 3,083,999) | 0.88 billion<br><br>(0.26 billion to 1.73 billion) | -244 million<br><br>(-592 million to -62 million) | -4.41 billion<br><br>(-5.57 billion to -3.32 billion) |
| Georgia | | | | | | | | | |
| The Intervention Package without TPT Compared to Status Quo | -6,257<br><br>(-11,640 to -3,560) | -772<br><br>(-1,269 to -496) | -29,180<br><br>(-48,822 to -18,580) | -33,210<br><br>(-55,279 to -21,131) | 2,650,774<br><br>(2,209,823 to 3,322,408) | -1,326<br><br>(-1,760 to -1,012) | 37 million<br><br>(4 million to 70 million) | -7 million<br><br>(-20 million to -1 million) | -0.16 billion<br><br>(-0.3 billion to -0.08 billion) |
| The Intervention Package with TPT Compared to The Intervention Package without TPT | -3,960<br><br>(-5,833 to -2,817) | -198<br><br>(-270 to -153) | -7,409<br><br>(-10,390 to -5,574) | -8,790<br><br>(-12,453 to -6,546) | -664,978<br><br>(-994,073 to -470,466) | 767,492<br><br>(639,060 to 953,900) | 22 million<br><br>(9 million to 40 million) | 31 million<br><br>(-2 million to 94 million) | 0 billion<br><br>(-0.04 billion to 0.07 billion) |
| Enhanced Package with TPT Compared to The Intervention Package with TPT | -3,285<br><br>(-4,753 to -2,385) | -297<br><br>(-403 to -229) | -10,700<br><br>(-14,828 to -8,095) | -12,579<br><br>(-17,579 to -9,472) | 771,545<br><br>(738,462 to 804,544) | 33,509<br><br>(24,740 to 37,001) | 5 million<br><br>(-13 million to 23 million) | -23 million<br><br>(-57 million to -5 million) | -0.09 billion<br><br>(-0.14 billion to -0.05 billion) |
| Kenya | | | | | | | | | |
| The Intervention Package without TPT Compared to Status Quo | -949,389<br><br>(-1,305,679 to -843,836) | -205,138<br><br>(-267,080 to -183,061) | -8,088,043<br><br>(-10,569,104 to -7,195,363) | -8,530,784<br><br>(-11,113,181 to -7,600,633) | 111,302,193<br><br>(95,913,969 to 139,289,951) | -5,368,074<br><br>(-6,678,016 to -4,593,241) | 1.3 billion<br><br>(-0.69 billion to 4.86 billion) | -68 million<br><br>(-160 million to -20 million) | -15.75 billion<br><br>(-21.18 billion to -11.84 billion) |
| The Intervention Package with TPT Compared to The Intervention Package without TPT | -416,229<br><br>(-488,785 to -372,738) | -30,233<br><br>(-34,971 to -27,657) | -1,260,062<br><br>(-1,476,157 to -1,151,223) | -1,389,712<br><br>(-1,623,051 to -1,269,646) | -72,928,397<br><br>(-98,018,033 to -61,582,099) | 26,371,419<br><br>(23,854,528 to 29,019,505) | -0.58 billion<br><br>(-1.91 billion to 0.17 billion) | 150 million<br><br>(-81 million to 513 million) | -3.08 billion<br><br>(-4.56 billion to -2.17 billion) |
| Enhanced Package with TPT Compared to The Intervention Package with TPT | -190,435<br><br>(-255,804 to -172,655) | -35,180<br><br>(-43,835 to -32,718) | -1,350,076<br><br>(-1,704,623 to -1,246,795) | -1,437,283<br><br>(-1,807,386 to -1,328,210) | 9,319,538<br><br>(8,521,039 to 10,642,534) | -710,321<br><br>(-1,112,244 to -495,451) | 0.87 billion<br><br>(-0.09 billion to 3.06 billion) | -72 million<br><br>(-179 million to -17 million) | -2.04 billion<br><br>(-3.26 billion to 0.13 billion) |
| South Africa | | | | | | | | | |

| Country and Comparison | Number Developing TB | Number Dying from TB | Years of Life Lost due to TB | TB-Associated DALYs | Number Screened for TB | Number Initiating TPT | TB-Related Health System Cost | TB-Associated Patient and Family Costs | TB-Related Societal Cost |
|---|---|---|---|---|---|---|---|---|---|
| The Intervention Package without TPT Compared to Status Quo | -991,591 (-1,584,431 to -550,257) | -248,602 (-400,609 to -103,835) | -9,007,443 (-14,131,410 to -3,810,368) | -9,530,038 (-14,849,978 to -4,102,683) | 169,165,222 (95,210,590 to 240,147,807) | -623,839 (-1,550,744 to -62,623) | 2.37 billion (-2.34 billion to 6.52 billion) | -158 million (-398 million to -30 million) | -58.83 billion (-93.74 billion to -21.82 billion) |
| The Intervention Package with TPT Compared to The Intervention Package without TPT | -436,806 (-539,529 to -262,913) | -27,416 (-35,379 to -17,280) | -1,204,573 (-1,515,401 to -711,698) | -1,363,026 (-1,690,753 to -814,873) | -98,640,091 (-157,751,972 to -38,906,890) | 37,403,007 (27,760,013 to 45,023,330) | -0.6 billion (-2.15 billion to 0.33 billion) | 228 million (-210 million to 859 million) | -8.54 billion (-11.58 billion to -4.45 billion) |
| Enhanced Package with TPT Compared to The Intervention Package with TPT | -138,302 (-223,343 to -96,675) | -40,185 (-63,057 to -21,442) | -1,379,814 (-2,095,105 to -760,291) | -1,469,541 (-2,200,674 to -823,075) | 25,856,738 (19,854,491 to 29,865,544) | -619,976 (-717,317 to -514,716) | 3.65 billion (0.91 billion to 9.43 billion) | -112 million (-313 million to -8 million) | -5.81 billion (-11.68 billion to 2.53 billion) |

Notes: Value represent mean (95% UR). All costs are in 2023 USD. No discounting.
Abbreviations: TPT, tuberculosis preventive treatment; TB, tuberculosis; DALY, disability-adjusted life years

#### Incremental Outcomes (Relative Values), by 2050, including the enhanced package.

| Country and Comparison | Number Developing TB | Number Dying from TB | Years of Life Lost due to TB | TB-Associated DALYs | Number Screened for TB | Number Initiating TPT | TB-Related Health System Cost | TB-Associated Patient and Family Costs | TB-Related Societal Cost |
|---|---|---|---|---|---|---|---|---|---|
| Brazil | | | | | | | | | |
| The Intervention Package without TPT Compared to Status Quo | -9.6% (-10% to -9.3%) | -14.1% (-14.6% to -13.8%) | -13.4% (-14% to -13.1%) | -10.8% (-11.3% to -10.4%) | 2217.6% (2132.4% to 2302.3%) | -3.6% (-4.3% to -3.2%) | 67.5% (1.9% to 148.1%) | -4% (-14.1% to -1.1%) | -11.2% (-12.8% to -9.9%) |
| The Intervention Package with TPT Compared to The Intervention Package without TPT | -9.5% (-9.8% to -9.3%) | -10.7% (-10.8% to -10.5%) | -10.3% (-10.5% to -10.2%) | -8.1% (-8.4% to -8%) | -10.8% (-11.7% to -10.2%) | 1781.4% (1714.5% to 1848.5%) | 7.8% (1.1% to 17.1%) | 44.6% (-3.2% to 202.7%) | -8.6% (-9.5% to -7.3%) |
| Enhanced Package with TPT Compared to The Intervention Package with TPT | -7.3% (-7.7% to -7%) | -8.6% (-9.1% to -8.4%) | -8.4% (-8.8% to -8.1%) | -6.5% (-6.9% to -6.2%) | 57.9% (56.6% to 58.7%) | 18.9% (18.6% to 19.2%) | 23.5% (7.7% to 45%) | -12.6% (-24.8% to -4.9%) | -6.7% (-7.9% to -5.2%) |
| Georgia | | | | | | | | | |
| The Intervention Package without TPT Compared to Status Quo | -14.4% (-19.6% to -11%) | -25.8% (-31.7% to -21.6%) | -26.4% (-32.4% to -22.2%) | -17.8% (-22.9% to -14.3%) | 18105.2% (13915.3% to 25251.7%) | -23.7% (-31.5% to -19.4%) | 85.8% (5.6% to 186.5%) | -11.5% (-26% to -4%) | -19.1% (-26.4% to -12.7%) |
| The Intervention Package with TPT Compared to The Intervention Package without TPT | -10.9% (-12.3% to -9.8%) | -9% (-9.8% to -8.4%) | -9.2% (-10.2% to -8.5%) | -5.8% (-6.7% to -5.2%) | -24.7% (-29.8% to -21.1%) | 18061.9% (14632.9% to 24569.9%) | 26.5% (9% to 51.4%) | 83.3% (-3.2% to 346.4%) | 0.6% (-5% to 10.5%) |
| Enhanced Package with TPT Compared to The Intervention Package with TPT | -10.1% (-11.3% to -9.2%) | -14.9% (-16.4% to -13.8%) | -14.7% (-16.2% to -13.6%) | -8.8% (-10% to -7.9%) | 38.7% (34.4% to 42.1%) | 4.4% (2.5% to 5.7%) | 5% (-10.8% to 21.1%) | -25.5% (-43.5% to -9.3%) | -12.9% (-17.7% to -8.7%) |
| Kenya | | | | | | | | | |
| The Intervention Package without TPT Compared to Status Quo | -30.3% (-33.1% to -29%) | -45.1% (-46.8% to -43%) | -45.4% (-46.6% to -43.5%) | -41.7% (-43.2% to -39.8%) | 878.4% (579.1% to 1156.1%) | -46.1% (-47.2% to -44.5%) | 49.6% (-14% to 172.6%) | -17.7% (-33.6% to -10.7%) | -38.4% (-42.5% to -30.1%) |
| The Intervention Package with TPT Compared to The Intervention Package without TPT | -19.2% (-20.1% to -17.6%) | -12.2% (-12.9% to -11.1%) | -13% (-13.9% to -11.8%) | -11.7% (-12.4% to -10.7%) | -58.5% (-63.6% to -55.2%) | 425.2% (299.9% to 509.6%) | -11.1% (-23% to 6.3%) | 77.3% (-12.5% to 356%) | -12.1% (-15.2% to -9.4%) |
| Enhanced Package with TPT Compared to The Intervention Package with TPT | -10.8% (-11.8% to -10.5%) | -16.1% (-16.4% to -15.7%) | -16% (-16.4% to -15.7%) | -13.7% (-13.9% to -13.4%) | 18.2% (17.4% to 19.5%) | -2.2% (-3.1% to -1.6%) | 20.1% (-2.3% to 49.8%) | -14.4% (-22.9% to -6.5%) | -9.3% (-14% to 0.4%) |
| South Africa | | | | | | | | | |
| The Intervention Package without TPT Compared to Status Quo | -22.7% (-27.2% to -19.4%) | -35.5% (-38.1% to -30%) | -36.8% (-39.7% to -29.8%) | -33.5% (-36.6% to -26.5%) | 3038% (1320.5% to 6025.8%) | -12% (-22.1% to -1.8%) | 154.9% (-31.5% to 534.4%) | -11.7% (-22% to -4.5%) | -34.3% (-38.5% to -25%) |

| Country and Comparison | Number Developing TB | Number Dying from TB | Years of Life Lost due to TB | TB-Associated DALYs | Number Screened for TB | Number Initiating TPT | TB-Related Health System Cost | TB-Associated Patient and Family Costs | TB-Related Societal Cost |
|---|---|---|---|---|---|---|---|---|---|
| The Intervention Package with TPT Compared to The Intervention Package without TPT | -13.1%<br>(-14.4% to -11.2%) | -6.4%<br>(-7.9% to -4.8%) | -8.2%<br>(-9.5% to -6.5%) | -7.5%<br>(-8.5% to -6.3%) | -54.4%<br>(-64.7% to -38.4%) | 929.8%<br>(551.9% to 1342.9%) | -9%<br>(-24.8% to 11.2%) | 29.8%<br>(-8.2% to 133.8%) | -7.9%<br>(-9.5% to -6.3%) |
| Enhanced Package with TPT Compared to The Intervention Package with TPT | -4.8%<br>(-7% to -4.1%) | -9.8%<br>(-10.7% to -9.3%) | -10%<br>(-10.9% to -9.4%) | -8.7%<br>(-9.6% to -8.3%) | 33.9%<br>(29% to 41.3%) | -1.5%<br>(-1.9% to -1.1%) | 74.8%<br>(25.9% to 157.2%) | -8.4%<br>(-21.3% to -0.6%) | -5.4%<br>(-8.9% to 3.6%) |

Notes: Value represent mean (95% UR). All costs are in 2023 USD. No discounting.

Abbreviations: TPT, tuberculosis preventive treatment; TB, tuberculosis; DALY, disability-adjusted life years

#### Incremental Outcomes (Absolute Values), by 2050, Slow Implementation (2024-2050) and Fast implementation (2024-2027)

| Country and Comparison | Number Developing TB | Number Dying from TB | Years of Life Lost due to TB | TB-Associated DALYs | Number Screened for TB | Number Initiating TPT | TB-Related Health System Cost | TB-Associated Patient and Family Costs | TB-Related Societal Cost |
|---|---|---|---|---|---|---|---|---|---|
| Brazil | | | | | | | | | |
| The Intervention Package with TPT Compared to The Status Quo (*PRIMARY*) | -484,511 (-540,492 to -448,152) | -52,270 (-58,301 to -48,368) | -1,938,763 (-2,181,239 to -1,783,135) | -2,023,594 (-2,279,453 to -1,859,500) | 36,750,999 (34,614,346 to 40,006,979) | 14,375,925 (13,525,835 to 15,683,633) | 1.56 billion (0.31 billion to 2.48 billion) | 380 million (-135 million to 1310 million) | -15.35 billion (-18.2 billion to -13.22 billion) |
| Slow Implementation of Intervention Package with TPT vs. Status Quo | -269,567 (-300,625 to -249,310) | -30,621 (-34,196 to -28,298) | -1,125,497 (-1,267,305 to -1,034,321) | -1,168,269 (-1,317,013 to -1,072,707) | 22,295,855 (20,879,352 to 24,473,380) | 9,272,919 (8,716,868 to 10,135,912) | 1.24 billion (0.04 billion to 2.03 billion) | 269 million (-32 million to 857 million) | -8.52 billion (-10.53 billion to -7.09 billion) |
| Fast Implementation of Intervention Package with TPT vs. Status Quo | -562,743 (-626,325 to -521,422) | -59,973 (-66,752 to -55,586) | -2,226,707 (-2,499,680 to -2,051,400) | -2,329,111 (-2,617,754 to -2,143,875) | 39,623,710 (37,371,668 to 43,050,841) | 15,461,831 (14,559,643 to 16,846,652) | 1.6 billion (0.33 billion to 2.53 billion) | 414 million (-180 million to 1462 million) | -17.85 billion (-20.98 billion to -15.46 billion) |
| Georgia | | | | | | | | | |
| The Intervention Package with TPT Compared to The Status Quo (*PRIMARY*) | -10,217 (-17,472 to -6,324) | -970 (-1,549 to -651) | -36,590 (-59,548 to -24,125) | -42,001 (-67,988 to -27,669) | 1,985,796 (1,739,282 to 2,327,543) | 766,166 (638,010 to 952,195) | 59 million (27 million to 92 million) | 24 million (-12 million to 88 million) | -0.16 billion (-0.31 billion to -0.06 billion) |
| Slow Implementation of Intervention Package with TPT vs. Status Quo | -5,454 (-10,091 to -3,125) | -575 (-967 to -364) | -21,722 (-37,257 to -13,586) | -24,621 (-42,072 to -15,361) | 1,369,253 (1,159,397 to 1,674,950) | 538,630 (432,927 to 698,551) | 44 million (15 million to 70 million) | 19 million (-7 million to 66 million) | -0.08 billion (-0.18 billion to -0.02 billion) |
| Fast Implementation of Intervention Package with TPT vs. Status Quo | -11,743 (-19,742 to -7,386) | -1,089 (-1,717 to -740) | -40,984 (-65,911 to -27,340) | -47,270 (-75,576 to -31,522) | 2,095,132 (1,846,345 to 2,437,296) | 811,300 (679,788 to 1,000,795) | 61 million (29 million to 95 million) | 25 million (-15 million to 92 million) | -0.19 billion (-0.35 billion to -0.08 billion) |
| Kenya | | | | | | | | | |
| The Intervention Package with TPT Compared to The Status Quo (*PRIMARY*) | -1,365,618 (-1,805,943 to -1,216,387) | -235,371 (-302,411 to -211,038) | -9,348,105 (-12,052,477 to -8,350,249) | -9,920,496 (-12,739,399 to -8,872,438) | 38,373,796 (34,118,754 to 42,720,081) | 21,003,345 (17,508,831 to 23,943,807) | 0.71 billion (-1.41 billion to 3.21 billion) | 82 million (-181 million to 453 million) | -18.83 billion (-24.56 billion to -15.48 billion) |
| Slow Implementation of Intervention Package with TPT vs. Status Quo | -939,844 (-1,280,476 to -824,816) | -169,821 (-222,514 to -149,961) | -6,780,151 (-8,892,968 to -5,967,322) | -7,170,525 (-9,376,828 to -6,319,098) | 31,460,595 (27,308,214 to 35,421,581) | 18,114,314 (14,845,662 to 20,890,580) | 0.64 billion (-0.98 billion to 2.56 billion) | 92 million (-89 million to 380 million) | -13.5 billion (-18.07 billion to -10.86 billion) |
| Fast Implementation of Intervention Package with TPT vs. Status Quo | -1,500,076 (-1,971,860 to -1,340,388) | -255,676 (-327,090 to -230,037) | -10,116,572 (-13,002,192 to -9,067,506) | -10,749,672 (-13,758,724 to -9,647,063) | 39,479,875 (35,173,640 to 43,946,524) | 21,473,440 (17,931,180 to 24,480,823) | 0.71 billion (-1.5 billion to 3.27 billion) | 84 million (-215 million to 494 million) | -20.44 billion (-26.5 billion to -16.99 billion) |
| South Africa | | | | | | | | | |
| The Intervention Package with TPT Compared to The Status Quo (*PRIMARY*) | -1,428,397 (-2,113,280 to -813,169) | -276,018 (-434,787 to -121,242) | -10,212,016 (-15,656,418 to -4,523,351) | -10,893,064 (-16,506,651 to -4,917,768) | 70,525,130 (55,054,057 to 82,169,398) | 36,779,168 (27,223,200 to 44,932,012) | 1.77 billion (-3.09 billion to 4.87 billion) | 70 million (-527 million to 628 million) | -67.37 billion (-103.95 billion to -26.75 billion) |

| Country and Comparison | Number Developing TB | Number Dying from TB | Years of Life Lost due to TB | TB-Associated DALYs | Number Screened for TB | Number Initiating TPT | TB-Related Health System Cost | TB-Associated Patient and Family Costs | TB-Related Societal Cost |
|---|---|---|---|---|---|---|---|---|---|
| Slow Implementation of Intervention Package with TPT vs. Status Quo | -881,629<br><br>(-1,356,428 to -493,049) | -183,076<br><br>(-288,713 to -77,768) | -6,908,681<br><br>(-10,710,490 to -2,907,203) | -7,328,274<br><br>(-11,284,691 to -3,142,406) | 55,656,881<br><br>(40,861,282 to 67,513,627) | 30,236,848<br><br>(21,174,383 to 38,039,053) | 1.37 billion<br><br>(-3.12 billion to 4.12 billion) | 129 million<br><br>(-196 million to 582 million) | -45.32 billion<br><br>(-70.93 billion to -16.78 billion) |
| Fast Implementation of Intervention Package with TPT vs. Status Quo | -1,657,779<br><br>(-2,411,468 to -944,232) | -309,893<br><br>(-489,554 to -136,457) | -11,376,229<br><br>(-17,451,506 to -5,084,460) | -12,167,351<br><br>(-18,437,281 to -5,544,951) | 72,166,121<br><br>(56,520,120 to 83,641,549) | 37,851,961<br><br>(28,345,940 to 46,075,091) | 1.75 billion<br><br>(-3.2 billion to 4.95 billion) | 38 million<br><br>(-667 million to 673 million) | -75.31 billion<br><br>(-116.15 billion to -30.58 billion) |

Notes: Value represent mean (95% UR). All costs are in 2023 USD. No discounting.

Abbreviations: TPT, tuberculosis preventive treatment; TB, tuberculosis; DALY, disability-adjusted life years

#### Cost Effectiveness and Return on Investment by 2050, Slow Implementation (2024-2050) and Fast implementation (2024-2027)

| Outcome | Brazil | Georgia | Kenya | South Africa |
|---|---|---|---|---|
| **Slow Implementation of Intervention Package with TPT vs. Status Quo** | | | | |
| Return on Investment | 7.8 | 2.9 | 22.2 | 34.1 |
| Incremental Health System Cost per DALY averted | $1065 | $1768 | 89 | 187 |
| Incremental Health System Cost per TB case averted | $4617 | $7983 | 677 | 1553 |
| Incremental Health System Cost per TB death averted | $40,642 | $75,722 | 3746 | 7479 |
| Incremental Societal Cost per DALY averted | Cost Saving | Cost Saving | Cost Saving | Cost Saving |
| Incremental Societal Cost per TB case averted | Cost Saving | Cost Saving | Cost Saving | Cost Saving |
| Incremental Societal Cost per TB death averted | Cost Saving | Cost Saving | Cost Saving | Cost Saving |
| **Fast Implementation of Intervention Package with TPT vs. Status Quo** | | | | |
| Return on Investment | 12.2 | 4 | 29.9 | 44.1 |
| Incremental Health System Cost per DALY averted | $685 | $1292 | 66 | 144 |
| Incremental Health System Cost per TB case averted | $2836 | $5202 | 472 | 1054 |
| Incremental Health System Cost per TB death averted | $26,615 | $56,098 | 2770 | 5638 |
| Incremental Societal Cost per DALY averted | Cost Saving | Cost Saving | Cost Saving | Cost Saving |
| Incremental Societal Cost per TB case averted | Cost Saving | Cost Saving | Cost Saving | Cost Saving |
| Incremental Societal Cost per TB death averted | Cost Saving | Cost Saving | Cost Saving | Cost Saving |
| **Intervention Package with TPT vs. Status Quo (PRIMARY ANALYSIS)** | | | | |
| Return on Investment | 10.8 | 3.7 | 27.4 | 39 |
| Incremental Health System Cost per DALY averted | $771 | $1402 | $72 | $163 |
| Incremental Health System Cost per TB case averted | $3219 | $5762 | $521 | $1240 |
| Incremental Health System Cost per TB death averted | $29,838 | $60,693 | $3025 | $6415 |
| Incremental Societal Cost per DALY averted | Cost Saving | Cost Saving | Cost Saving | Cost Saving |
| Incremental Societal Cost per TB case averted | Cost Saving | Cost Saving | Cost Saving | Cost Saving |
| Incremental Societal Cost per TB death averted | Cost Saving | Cost Saving | Cost Saving | Cost Saving |

Notes: All costs are in 2023 USD. Return on investment is defined as the societal return per $1 USD invested by the health system. It cannot be calculated if there are projected health system cost savings. A return on investment of 8 means that $8 USD are returned to society for every $1 USD invested by the health system. Willingness to pay per DALY averted by country—Brazil, $13,644; Georgia, $1603; Kenya, $1002; South Africa, $4834.
Abbreviations: TPT, tuberculosis preventive treatment; DALY, disability adjusted life year; TB, tuberculosis

#### Incremental Outcomes (Absolute Values), by 2050, No Tuberculosis Infection Testing for Household Contacts

| Country and Comparison | Number Developing TB | Number Dying from TB | Years of Life Lost due to TB | TB-Associated DALYs | Number Screened for TB | Number Initiating TPT | TB-Related Health System Cost | TB-Associated Patient and Family Costs | TB-Related Societal Cost |
|---|---|---|---|---|---|---|---|---|---|
| Brazil | | | | | | | | | |
| The Intervention Package with TPT and no TB Infection Testing Compared to The Status Quo | -543,807 (-611,392 to -499,770) | -55,822 (-62,598 to -51,442) | -2,088,335 (-2,362,938 to -1,912,169) | -2,180,764 (-2,471,542 to -1,993,824) | 33,985,251 (32,176,805 to 36,727,160) | 34,760,350 (32,957,091 to 37,498,219) | 2.03 billion (0.75 billion to 3.18 billion) | 1394 million (-14 million to 4210 million) | -15.2 billion (-18.83 billion to -11.7 billion) |
| The Intervention Package with TPT Compared to The Status Quo (*PRIMARY*) | -484,511 (-540,492 to -448,152) | -52,270 (-58,301 to -48,368) | -1,938,763 (-2,181,239 to -1,783,135) | -2,023,594 (-2,279,453 to -1,859,500) | 36,750,999 (34,614,346 to 40,006,979) | 14,375,925 (13,525,835 to 15,683,633) | 1.56 billion (0.31 billion to 2.48 billion) | 380 million (-135 million to 1310 million) | -15.35 billion (-18.2 billion to -13.22 billion) |
| Georgia | | | | | | | | | |
| The Intervention Package with TPT and no TB Infection Testing Compared to The Status Quo | -11,942 (-20,123 to -7,483) | -1,043 (-1,660 to -704) | -39,546 (-64,157 to -26,136) | -45,420 (-73,422 to -29,958) | 1,676,108 (1,496,576 to 1,918,123) | 1,644,741 (1,469,677 to 1,880,802) | 86 million (46 million to 134 million) | 82 million (-5 million to 251 million) | -0.09 billion (-0.28 billion to 0.1 billion) |
| The Intervention Package with TPT Compared to The Status Quo (*PRIMARY*) | -10,217 (-17,472 to -6,324) | -970 (-1,549 to -651) | -36,590 (-59,548 to -24,125) | -42,001 (-67,988 to -27,669) | 1,985,796 (1,739,282 to 2,327,543) | 766,166 (638,010 to 952,195) | 59 million (27 million to 92 million) | 24 million (-12 million to 88 million) | -0.16 billion (-0.31 billion to -0.06 billion) |
| Kenya | | | | | | | | | |
| The Intervention Package with TPT and no TB Infection Testing Compared to The Status Quo | -1,503,761 (-1,954,505 to -1,358,468) | -244,148 (-311,735 to -220,057) | -9,733,991 (-12,475,894 to -8,741,606) | -10,350,591 (-13,215,989 to -9,311,844) | 28,447,934 (23,812,082 to 32,150,567) | 28,431,372 (25,402,292 to 31,187,990) | 0.29 billion (-1.77 billion to 2.33 billion) | 106 million (-217 million to 532 million) | -20.03 billion (-25.89 billion to -16.96 billion) |
| The Intervention Package with TPT Compared to The Status Quo (*PRIMARY*) | -1,365,618 (-1,805,943 to -1,216,387) | -235,371 (-302,411 to -211,038) | -9,348,105 (-12,052,477 to -8,350,249) | -9,920,496 (-12,739,399 to -8,872,438) | 38,373,796 (34,118,754 to 42,720,081) | 21,003,345 (17,508,831 to 23,943,807) | 0.71 billion (-1.41 billion to 3.21 billion) | 82 million (-181 million to 453 million) | -18.83 billion (-24.56 billion to -15.48 billion) |
| South Africa | | | | | | | | | |
| The Intervention Package with TPT and no TB Infection Testing Compared to The Status Quo | -1,566,440 (-2,290,546 to -874,784) | -287,332 (-450,527 to -126,687) | -10,683,225 (-16,229,854 to -4,746,150) | -11,410,677 (-17,157,488 to -5,157,625) | 51,214,856 (41,475,169 to 59,144,529) | 52,673,554 (44,311,089 to 58,668,793) | 1.37 billion (-3.58 billion to 4.26 billion) | 189 million (-602 million to 1076 million) | -70.84 billion (-108.46 billion to -28.62 billion) |
| The Intervention Package with TPT Compared to The Status Quo (*PRIMARY*) | -1,428,397 (-2,113,280 to -813,169) | -276,018 (-434,787 to -121,242) | -10,212,016 (-15,656,418 to -4,523,351) | -10,893,064 (-16,506,651 to -4,917,768) | 70,525,130 (55,054,057 to 82,169,398) | 36,779,168 (27,223,200 to 44,932,012) | 1.77 billion (-3.09 billion to 4.87 billion) | 70 million (-527 million to 628 million) | -67.37 billion (-103.95 billion to -26.75 billion) |

Notes: Value represent mean (95% UR). All costs are in 2023 USD. No discounting.

Abbreviations: TPT, tuberculosis preventive treatment; TB, tuberculosis; DALY, disability-adjusted life years

#### Cost Effectiveness and Return on Investment by 2050, No Tuberculosis Infection Testing for Household Contacts

| Outcome | Brazil | Georgia | Kenya | South Africa |
|---|---|---|---|---|
| **The Intervention Package with TPT and no TB Infection Testing vs. Status Quo** | | | | |
| Return on Investment | 8.5 | 2.1 | 69.6 | 52.7 |
| Incremental Health System Cost per DALY averted | $931 | $1885 | $28 | $120 |
| Incremental Health System Cost per TB case averted | $3735 | $7168 | $194 | $874 |
| Incremental Health System Cost per TB death averted | $36,385 | $82,066 | $1195 | $4767 |
| Incremental Societal Cost per DALY averted | Cost Saving | Cost Saving | Cost Saving | Cost Saving |
| Incremental Societal Cost per TB case averted | Cost Saving | Cost Saving | Cost Saving | Cost Saving |
| Incremental Societal Cost per TB death averted | Cost Saving | Cost Saving | Cost Saving | Cost Saving |
| **Intervention Package with TPT vs. Status Quo (PRIMARY ANALYSIS)** | | | | |
| Return on Investment | 10.8 | 3.7 | 27.4 | 39 |
| Incremental Health System Cost per DALY averted | $771 | $1402 | $72 | $163 |
| Incremental Health System Cost per TB case averted | $3219 | $5762 | $521 | $1240 |
| Incremental Health System Cost per TB death averted | $29,838 | $60,693 | $3025 | $6415 |
| Incremental Societal Cost per DALY averted | Cost Saving | Cost Saving | Cost Saving | Cost Saving |
| Incremental Societal Cost per TB case averted | Cost Saving | Cost Saving | Cost Saving | Cost Saving |
| Incremental Societal Cost per TB death averted | Cost Saving | Cost Saving | Cost Saving | Cost Saving |

Notes: All costs are in 2023 USD. Return on investment is defined as the societal return per $1 USD invested by the health system. It cannot be calculated if there are projected health system cost savings. A return on investment of 8 means that $8 USD are returned to society for every $1 USD invested by the health system. Willingness to pay per DALY averted by country—Brazil, $13,644; Georgia, $1603; Kenya, $1002; South Africa, $4834.

Abbreviations: TPT, tuberculosis preventive treatment; DALY, disability adjusted life year; TB, tuberculosis
